# Genetic study of psoriasis highlights its close link with socio-economic status and affective symptoms

**DOI:** 10.1101/2023.03.24.23287463

**Authors:** Anni Heikkilä, Eeva Sliz, Laura Huilaja, FinnGen, Estonian Biobank Research Team, Kadri Reis, Priit Palta, Abdelrahman G. Elnahas, Anu Reigo, Tõnu Esko, Triin Laisk, Maris Teder-Laving, Kaisa Tasanen, Johannes Kettunen

## Abstract

Psoriasis is an inflammatory skin disease with an estimated heritability of around 70 %. Previous genome-wide association studies (GWASs) have detected several risk loci for psoriasis. To further improve the understanding of the genetic risk factors impacting the disease, we conducted a discovery GWAS in FinnGen and a subsequent replication and meta-analysis with data from the Estonian Biobank and the UK biobank; the study sample included 925 649 individuals (22 659 cases and 902 990 controls), the largest sample for psoriasis yet. In addition, we conducted downstream analyses to find out more about psoriasis’ cross-trait genetic correlations and causal relationships. We report 8 novel risk loci, most of which harbor genes related to nuclear factor kappa-light-chain-enhancer of activated B-cells-signaling pathway and overall immunity. Genetic correlations highlight the relationship between psoriasis and smoking, higher body weight, and lower education level. Additionally, we report novel causal relationships between psoriasis and mood symptoms, as well as two-directioned causal relationship between psoriasis and lower education level. Our results provide new knowledge on psoriasis risk factors, which may be useful in the development of future treatment strategies.

## Introduction

Psoriasis is a chronic autoinflammatory disease typically manifesting as sharply demarcated erythematous scaling plaques on the skin. The exact symptoms depend on the subtype of the disease, the most common being psoriasis vulgaris, which affects around 90 % of the psoriasis patients^1^. Both genetic and environmental factors play a role in psoriasis pathogenesis. Heritability has been estimated to be around 70 % in twin studies^2^. Known risk loci include the human leukocyte antigen (HLA) region, especially HLA-C^3^. Additionally, for example many nuclear factor kappa-light-chain-enhancer of activated B-cells (NF-κB)-^4^ and interleukin (IL)-23/IL-27-signaling pathway-related genes^5^ have been associated with psoriasis. Known epidemiological risk factors affecting psoriasis risk include smoking^6^, obesity^7^, and stress^8^, among others. Psoriasis also has multiple comorbidities^9^,including cardiovascular diseases, other inflammatory diseases, and psychiatric disorders^10^.

The aim of this study was to find new risk loci for psoriasis. To do this, we analyzed genome-wide data of over 900 000 people from Finland (FinnGen), Estonia (The Estonian Biobank), and United Kingdom (UK Biobank). We further utilized the newly generated genome-wide results to evaluate genetic correlations and causal inferences between psoriasis and other traits.

## Materials and methods

### Study populations

#### FinnGen

(www.finngen.fi/en) is a Finnish collaboration project that several public and private partners are taking part in. The aim of the project is to collect 500 000 samples of genotype data combined with health record data from Finnish registries. We used FinnGen data freeze 7, in which there were 6 995 cases diagnosed with psoriasis (one or more diagnoses with International Classification of Diseases (ICD)-10 codes L40.0-L40.9) and 299 128 controls.

FinnGen participants provided written informed consent for biobank research, based on the Finnish Biobank Act. More detailed description of the consents, ethics committees and study approvals are presented in the **Supplementary Note**.

#### Estonian Biobank

(EstBB) is a population-based biobank at the Institute of Genomics, University of Tartu. The current cohort size is 200,000 individuals (aged ≥ 18), reflecting the age, sex and geographical distribution of the adult Estonian population. Estonian samples represent 83%, Russians 14%, and other ethnicities 3% of all participants. All subjects have been recruited by general practitioners (GP), physicians in hospitals and during promotional events. The EstBB database is linked with national registries (such as Cancer Registry and Causes of Death Registry), hospital databases, and the database of the national health insurance fund, which holds treatment and procedure service bills.

Diseases and health problems are recorded as ICD-10 codes and prescribed medicine according to the ATC classification. These health data are continuously updated through periodical linking to national electronic databases and registries. All participants were genotyped with genome-wide chip arrays and further imputed with a population-specific imputation panel consisting of 2,244 high-coverage (30x) whole genome sequenced individuals and 16,271,975 high-quality variants^11^. Association analysis was conducted with SAIGE^12^ mixed models with age, sex and 10 PCs used as covariates.

Here, individuals with psoriasis were identified using the ICD-10 codes including L40. EstBB GWAS analysis is supported by Personal research funding: Team grant PRG1291. Computations were performed in the High Performance Computing Center, University of Tartu.

#### UK Biobank

(UKBB)^13^ is a large biobank project launched in United Kingdom, with 40 to 69 years old adults as participants. The phenotypes were identified with registry data and questionnaires. The project is aiming to follow up participants for at least 30 years. The PanUKBB (https://pan.ukbb.broadinstitute.org/) project performed a pan-ancestry genetic analysis for 500 000 UKBB samples. We used the subset of European ancestry for this analysis, which consisted of 2 868 psoriasis cases and 417 663 controls.

### Genotyping, imputation, and quality control

In FinnGen, the samples were genotyped using Illumina and Affymetrix arrays (Illumina Inc., San Diego and Thermo Fischer Scientific, Santa Clara, CA, USA). Sample quality control (QC) was performed to exclude individuals with genotype missingness of over 5 %, ambiguous gender, excess heterozygosity (±4 SDs), and non-Finnish ancestry. Additionally, variants with low Hardy-Weinberg equilibrium (p < 1 * 10^−6^), over 2 % missingness or minor allele count under 3 %, were excluded from analysis in variant QC. Prephasing was done using Eagle 2.3.5. Imputation was performed with Beagle 4.1., with Finnish population specific SISu v3 reference panel. In post-imputation QC, variants with imputation info < 0.6 were excluded from further analyses.

### GWAS and meta-analysis

Psoriasis GWAS in FinnGen was performed using regenie^14^ software. The association model was adjusted for age, sex, the first 10 genetic principal components, and genotyping batch.

For meta-analysis, EstBB and UKBB summary statistics were lifted from hg19 to hg38 build. A Python-based software was used for inverse-variance weighted meta-analysis (available at https://github.com/FINNGEN/META_ANALYSIS/). Cochran’s Q-test was performed to estimate heterogeneity.

### Characterization of association signals

The association was considered novel, if there were no previous psoriasis associations reported in a ±0.5 MB window from the lead variant. In the novel loci, all genes in a ±0.5 MB window from the lead variant were annotated. GenBank^15^ and UniProt^16^ were used to obtain information about each gene. Additionally, broad literature research was conducted on genes of interest. Based on this attained knowledge about the biological functions and relevance of the genes, we aimed to find the gene most likely to have a role in the pathogenesis of psoriasis. Further, to perform functional gene mapping and gene-based association and enrichment tests, we run Functional Mapping and Annotation of Genome-Wide Association Studies (FUMA)^17^ analysis for the psoriasis summary statistics. Additionally, the Genotype-Tissue Expression (GTEx)^18^ data was checked to study the tissue expression of the genes of interest and to identify potential associations between psoriasis lead variants and altered gene expression.

To estimate the SNP-based heritability and genomic inflation factor, we used the linkage disequilibrium score (LDSC) pipeline tool^19^.

### Genetic correlations

We further applied LDSC regression to estimate genetic correlations between psoriasis and other phenotypes. We tested the correlations of psoriasis with 772 phenotypes available in the in-house LDSC analysis pipeline. The major histocompatibility complex was excluded from the analysis, as suggested by developers of the method^20^.

### Mendelian randomization

We used “TwoSampleMR” R library^21,22^ to conduct bi-directional two-sample Mendelian randomization between psoriasis and a selection of traits found to be genetically correlated with psoriasis. To avoid sample overlap, we used FinnGen summary data to extract genetic instruments for psoriasis. For other traits, we used mostly UKBB-based data provided by the Medical Research Council Integrative Epidemiology Unit (MRC IEU) database and implemented in the TwoSampleMR. LD clumping was performed with “clump_data” function and its default values, i.e., r^2^=0.001, a clumping window of 10 kb, and European population reference. We considered the inverse-variance weighted (IVW) method as the primary analysis. Egger intercepts were used to evaluate horizontal pleiotropy. Additionally, we tested heterogeneity with Cochrans Q test (“mr_heterogeneity”) and performed leave-one-out analysis (“mr_leaveoneout”). A causal effect was deemed significant if MR Egger estimates obtained in the sensitivity analyses were in a matching direction with the IVW-estimates and false-discovery rate (FDR)-corrected p-value was less than 0.05.

## Results

### Four novel psoriasis loci discovered in FinnGen

In the discovery GWAS conducted in FinnGen in 306 123 individuals (**Figure 1A, Supplementary Table S1**), we found 38 genome-wide significant loci, 4 of which were novel (**Figure 2**; **Table 1**): the associations at 2q32.3, 5q35.3, 7p22.1, and 16p12.1 had not been associated with psoriasis in prior studies. The lead variant at 2q32.3, rs77509633, is an intergenic variant near solute carrier family 39 member 10 (*SLC39A10*; also known as *ZIP10*). The ZIP10 zinc carrier protein coded by this gene has been found to express abnormally in patients with atopic dermatitis^23^. ZIP10 is also indicated to have a role in humoral immunity^24^. The novel association at 5q35.3, with a lead variant rs13153019, locates near fibroblast growth factor receptor 4 (*FGFR4*). It acts as a receptor for fibroblast growth factor 19 (*FGF19*), which has been found to regulate the proliferation of keratinocytes in psoriasis skin ^25^. The study also found that the inhibition of *FGFR4* attenuates the effect of *FGF19* on keratinocytes. The novel association at 7p22.1, with a lead variant rs748670681, is near ring finger protein 216 (*RNF216*), which is known to participate in the regulation of the NF-κB signaling pathway^26^. At 16p12.1, the lead variant rs144651842 is a missense variant in interleukin 4 receptor (*IL4R*), which acts as a receptor for interleukins 4 and 13, both of which have been associated with psoriasis in earlier GWASs^5,27^.

**Figure 1A-B.**
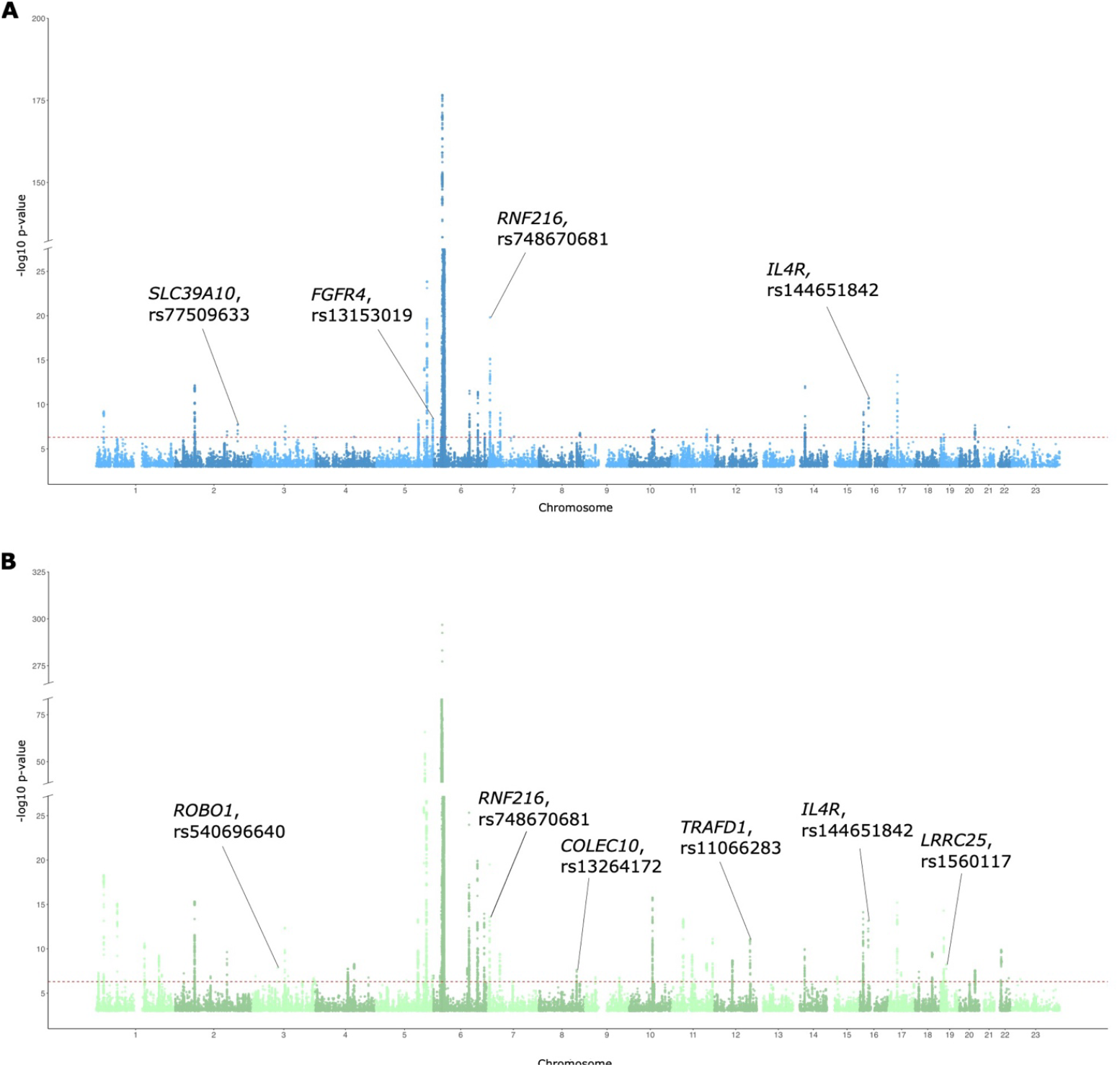
Manhattan plots of A) FinnGen-based discovery GWAS and B) FinnGen, EstBB (Estonian BioBank) and UKBB (UK Biobank) meta-analysis GWAS. Candidate genes and rs-numbers for novel lead variants are marked in each plot. Y-axis represents the -log10 p-value. The genome-wide significance threshold is marked with dashed horizontal line.

**Figure 2A-D.**
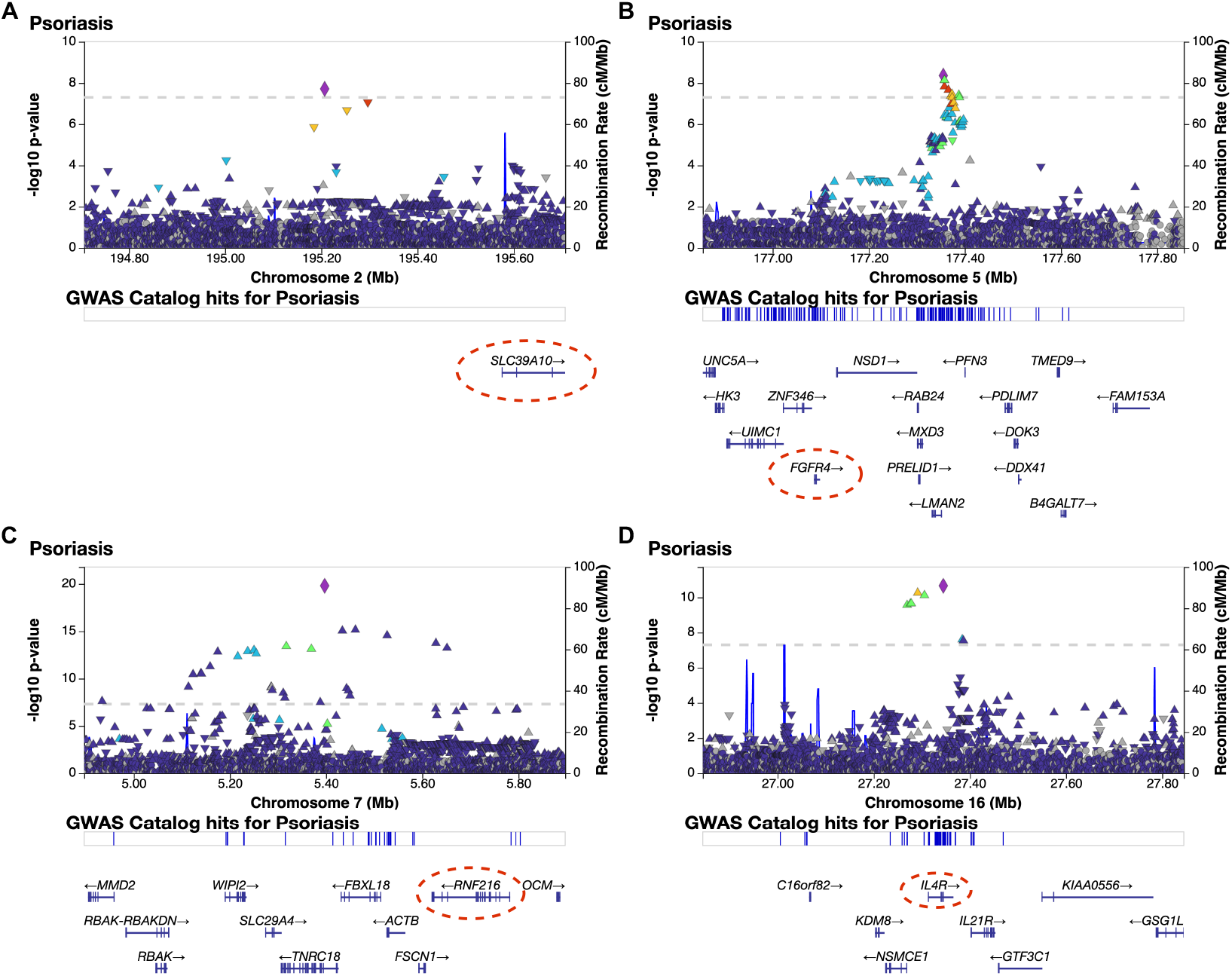
Regional association plots (LocusZooms) of the discovery GWAS (FinnGen-only) novel risk loci (±0.5 MB around lead variant) at: A) 2q32.3, B) 5q35.3, C) 7p22.1, D) 16p12.1. Candidate genes are circled in the figures with red dashed line.

**Table 1.**
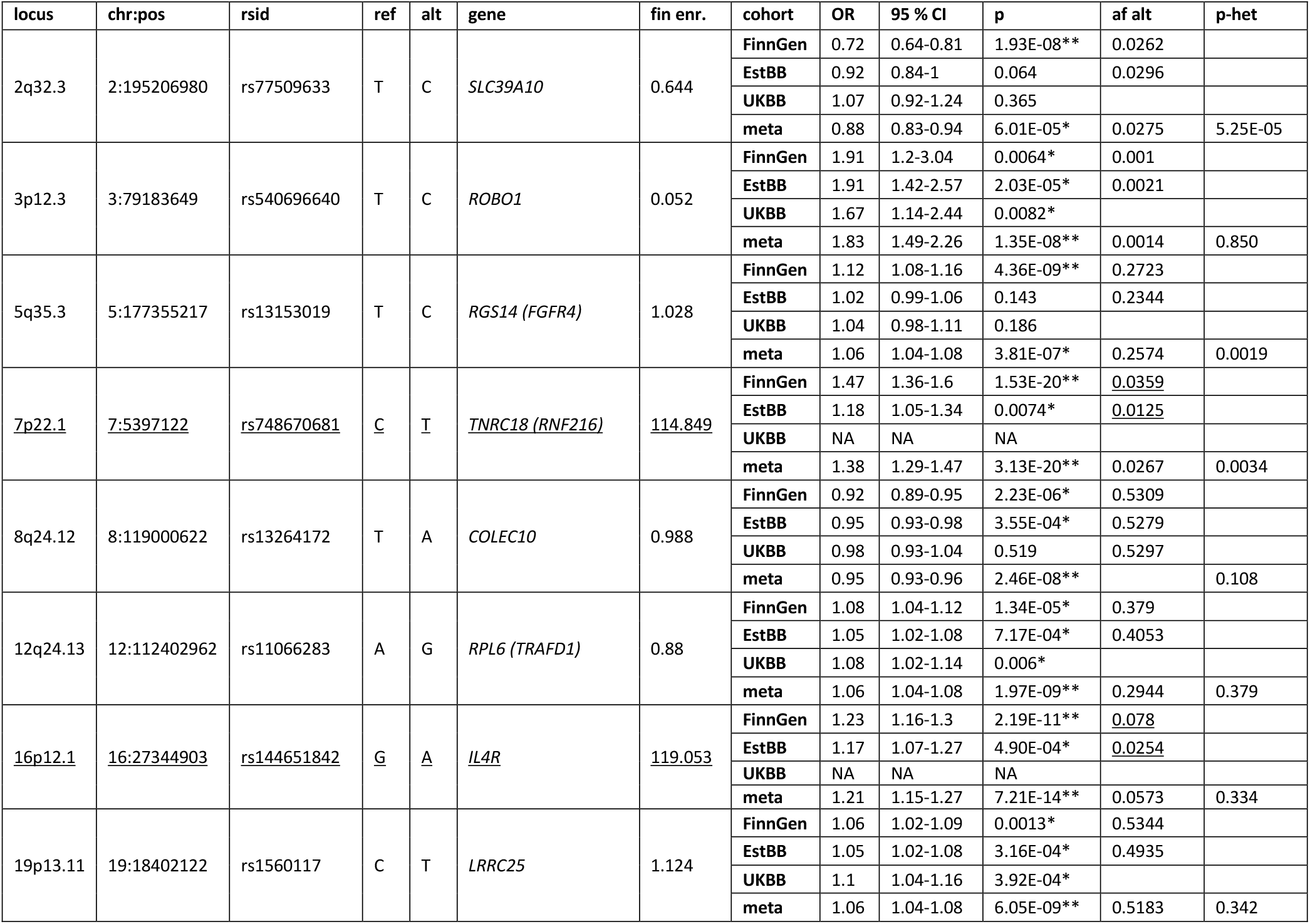
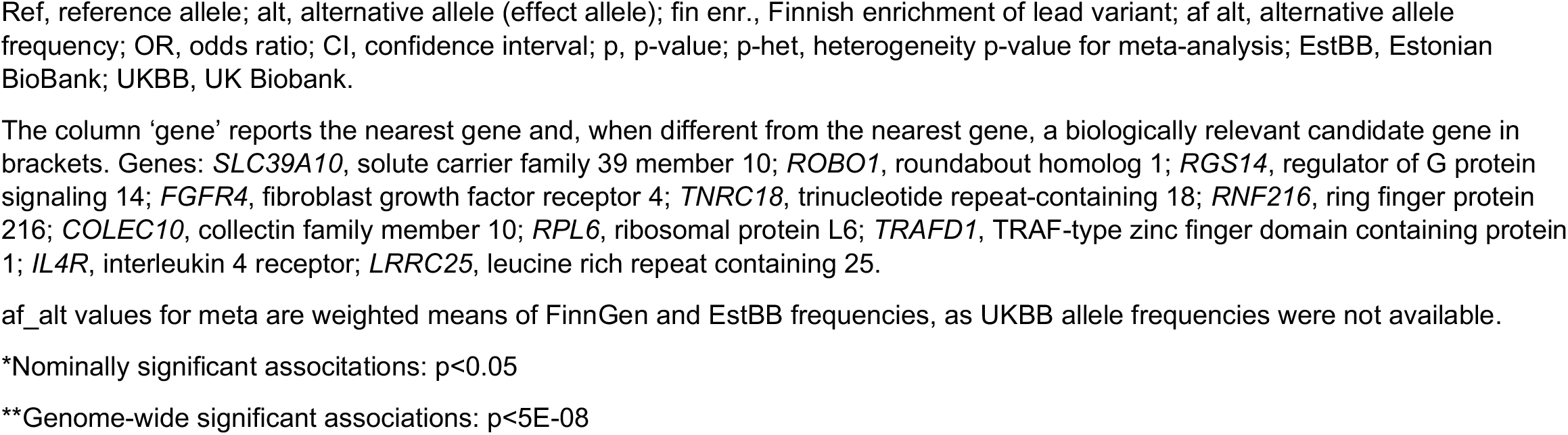
Novel psoriasis risk loci in the discovery GWAS (FinnGen) and meta-analysis. Associations significant in both discovery GWAS and meta-analysis (in loci 7p22.1 and 16p12.1) are underlined.

The novel loci detected in the discovery GWAS mostly appeared to be specific to the Finnish population (**Table 1**; **Supplementary Figure S1**). The associations near *RNF216* and *IL4R* replicated on a nominal level in EstBB (respective p-values of the lead variants in the three populations: rs748670681 FinnGen 1.53 * 10^−20^, EstBB 0.0074, variant not available in UKBB; rs144651842 FinnGen 2.19 * 10^−11^, EstBB 0.00049, variant not available in UKBB) whereas the associations near *SLC39A10* and *FGFR4* reached significance in FinnGen only (respective p-values of the lead variants in the three poplations: rs77509633 FinnGen 1.93 * 10^−8^, EstBB 0.064, UKBB 0.365; rs13153019 FinnGen 4.36 * 10^−9^, EstBB 0.143, UKBB 0.186). The association signals near *RNF216* and *IL4R* were driven by variants enriched in the Finnish population with respective Finnish enrichment values of 114.849 and 119.053 calculated as ratios of the Finnish allele frequency and the non-Finnish-non-Estonian European allele frequency (**Table 1**).

### Meta-analysis discovers additional four novel psoriasis loci

In the meta-analysis of FinnGen, EstBB and UKBB, totalling up to 925 649 individuals in the study (**Figure 1B, Supplementary Table S2**), we found altogether 57 loci associated with psoriasis; of these, four loci were not reported in previous studies nor detected in the discovery GWAS (**Figure 3**; **Table 1**). These included the associations at 3p12.3, 8q24.12, 12q24.13, and 19p13.11. In addition, the associations at 7p22.1 near *RNF216* and 16p12.1 near *ILR4* attained genome-wide significance in the meta-analysis.

**Figure 3A-F.**
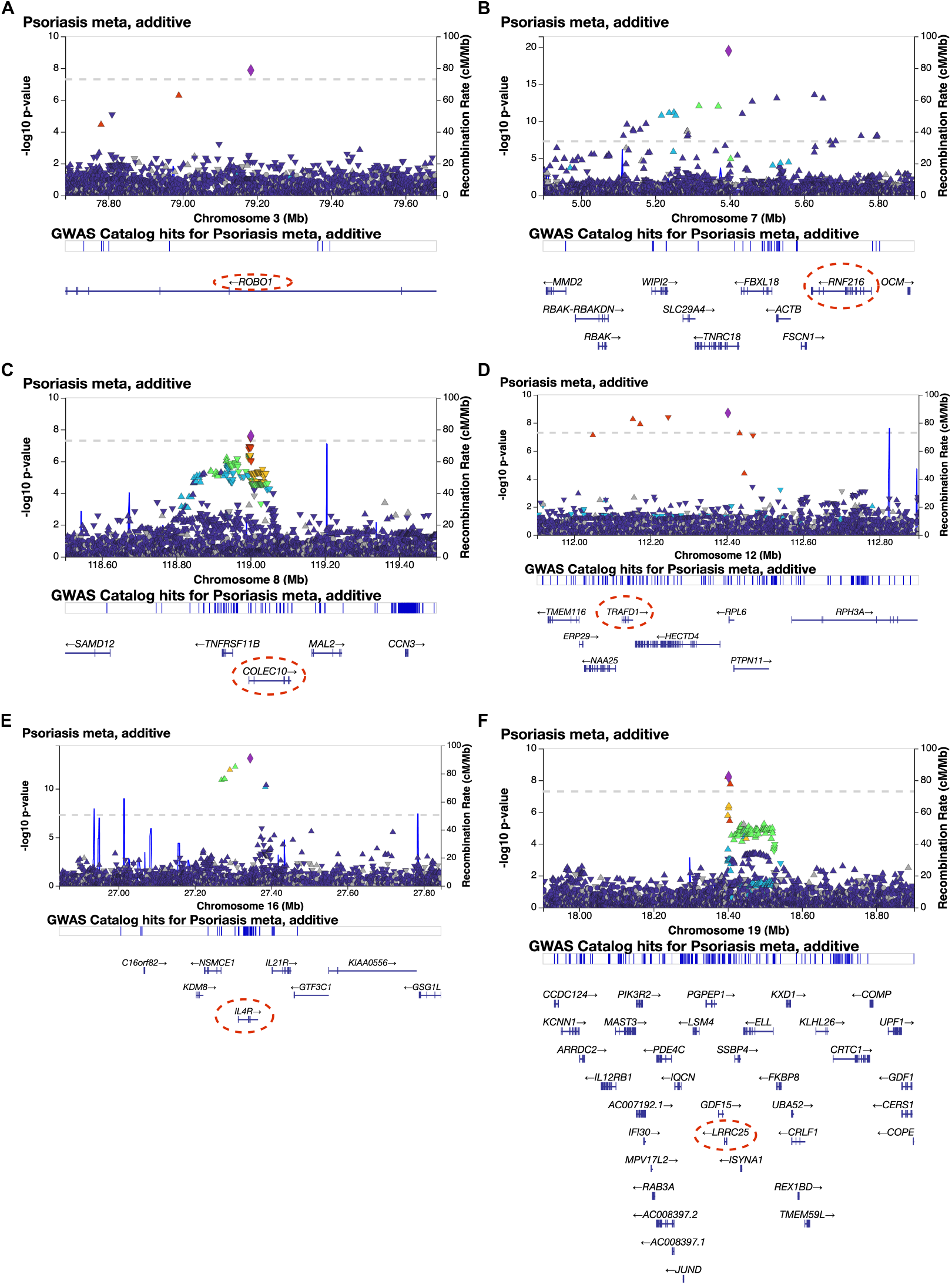
Regional association plots (LocusZooms) of meta-analysis (FinnGen, Estonian Biobank (EstBB) and UK Biobank (UKBB) novel risk loci (±0.5 MB around lead variant) at: A) 3p12.3, B) 7p22.1, C) 8q24.12, D) 12q24.13, E) 16p12.1, F) 19p13.11. Candidate genes are circled in the figures with red dashed line.

At 3p12.3, the lead variant rs540696640 is located in the intron area of roundabout homolog 1 (*ROBO1*). It is part of the Slit/Robo signaling pathway, which is known to affect various processes including cell migration, organogenesis and angiogenesis^28,29^. At 8q24.12, the lead variant rs13264172 is in the intron area of collectin family member 10 (*COLEC10*) encoding collectin liver 1 (CL-L1), which belongs to the collectin family. Collectins detect pathogens, so even though CL-L1’s function is not apparent yet, it’s probable that it has a role in innate immunity^30^. Serum levels of CL-L1 and collectin kidney 1 (CL-K1), another collectin family member often forming a complex with CL-L1, have been found to be lower in patients suffering from an autoimmune disease, systemic lupus erythematosus^31^. At 12q24.13, the gene nearest to the lead variant rs11066283 is ribosomal protein L6 (*RPL6*), but it has no apparent biological role in psoriasis pathogenesis. A nearby TRAF-type zinc finger domain containing protein 1 (*TRAFD1*), also known as *FLN29*, has a function that is strongly related to innate immunity and NF-κB pathway^32^, as it has been suggested to negatively regulate Toll-like receptor (TLR) and retinoic acid-inducible gene I (RIG-I)-like pathways^33^ and, thus, appears as a more likely candidate to drive the psoriasis association in this locus. At 19p13.11, the lead variant rs1560117 locates near leucine-rich repeat containing 25 (*LRRC25*), which has a role in the inhibition of the NF-κB signaling pathway by promoting degradation of p65/RelA, one of NF-κBs transcription factors^34^.

Of the novel loci showing genome-wide significant association with psoriasis in meta-analysis, only *RNF216* locus showed nominally significant heterogeneity (p-value for heterogeneity measured with Cochran’s Q test [p-het]=0.0034; **Table 1**). This suggest that the results were highly matching across the study populations, as also indicated by the matching odds ratios (**Supplementary Figure S1**).

### Psoriasis loci are enriched for inflammation-related genes

Using the psoriasis GWAS results from the meta-analysis, we performed a Multi-marker Analysis of GenoMic Annotation (MAGMA) enrichment analysis^35^ as implemented in FUMA^17^. Psoriasis loci were significantly enriched for 20 inflammation-related gene sets (**Supplementary Figure S2**), including ‘gene ontology (GO) cellular component (cc): major histocompatibility complex (mhc) protein complex’ (p = 1.45 * 10^−10^), ‘GO biological process (bp): cytokine-mediated signaling pathway’ (p = 1.49 * 10^−8^), ‘GO bp: regulation of defense response’ (p = 3.68 * 10^−8^), and ‘GO bp: positive regulation of immune system process’ (p = 1.40 * 10^−7^).

### LDSC

Altogether 319 genetic correlations (r_g_) tested with LDSC pipeline were found significant after FDR correction. Psoriasis was genetically correlated with many anthropometric traits (e.g., whole body fat mass [r_g_=0.20, p=6.89 * 10^−12^], body fat percentage [r_g_=0.20, p=1.39 * 10-^11^]), smoking (e.g., current tobacco smoking [r_g_=0.29, p=3.43 * 10^−13^]), level of education (e.g., years of schooling [r_g_=-0.25, p=8.32 * 10^−15^], qualifications: A levels/AS levels or equivalent [r_g_=-0.26, p=6.58 * 10^−13^], qualifications: college or university degree [r_g_=-0.25, p=2.57 * 10^−12^]), other diseases (e.g., type 2 diabetes [r_g_=0.27, p=7.23 * 10^−10^], major depressive disorder [r_g_=0.32, p=1.30 * 10^−5^]) or mood (e.g., miserableness [r_g_=0.19, p=2.08 *10^−7^], frequency of tiredness/lethargy in last 2 weeks [r_g_=0.20, p=2.86 * 10^−7^]) (**Figure 4**). Additionally, correlations include many nuclear magnetic resonance (NMR)- based metabolic measures (e.g., ratio of linoleic acid to total fatty acids [r_g_=-0.27, p=3.71 * 10^−7^], ratio of polyunsaturated fatty acids to total fatty acids [r_g_=-0.22, p=5.30 *10^−7^] and ratio of polyunsaturated fatty acids to monounsaturated fatty acids [r_g_=-0.19, p=1.06 * 10^−^ _5_]), financial difficulties (e.g., ‘illness, injury, bereavement, stress in last 2 years: financial difficulties’ [r_g_=0.26, p=1.08 * 10^−7^), financial situation satisfaction (r_g_=0.29, p=8.96 * 10^−7^), ‘illness, injury, bereavement, stress in last 2 years: Serious illness, injury or assault to yourself’ (r_g_=0.29, p=7.05 * 10^−6^) or low age at giving birth (age at first live birth [r_g_=-0.24, p=7.44 * 10^−9^], age at last live birth [r_g_=-0.22, p=3.39 * 10^−6^]) (**Supplementary Table S3**).

**Figure 4.**
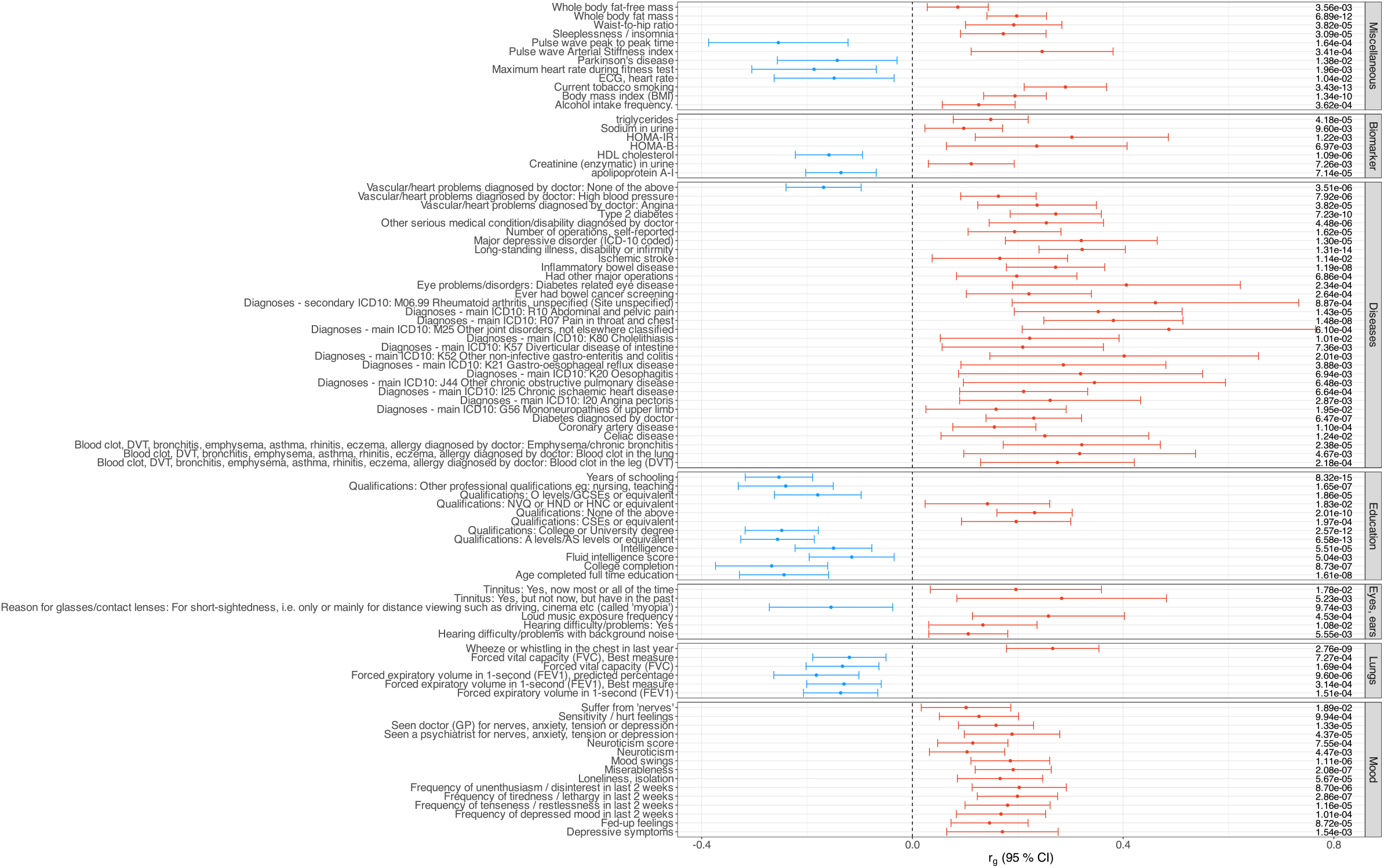
Forest plot of genetic correlations with psoriasis selected for Mendelian randomization analysis. Plot contains 99 out of total 319 significant associations (**Supplementary Table S3**). Traits (on the left side of the plot) are sorted by category (right side of the plot). Genetic correlation (rg) is indicated with a dot and the whiskers show 95 % confidence interval. P-value of each correlation is listed on the right side of the plot. Trait names are as in the LDSC pipeline output, but some category names have been shortened or modified (full original results available in supplements).

### MR

We tested 99 phenotypes for causality, based on the key traits of LDSC regression results. List of tested traits and studies used along with descriptions of certain traits are available in **Supplementary Table S4**. After FDR-correction and removing traits showing significant pleiotropy, differing direction of causal estimates using IVW and MR Egger methods, or if the result was based on only a few SNPs, we detected altogether 9 phenotypes that are causal to psoriasis and 18 that psoriasis is causal to (**Figure 5, Supplementary Table S5-6**). Results suggest that following traits are causal to psoriasis: Years of schooling (p=6.12 * 10^−10^), Body mass index (BMI) (4.51 * 10^−9^), Whole body fat mass (4.41 * 10^−7^), Qualifications: College or University degree (2.68 * 10^−5^), Frequency of tiredness/ lethargy in last 2 weeks (1.37 * 10^−4^), Inflammatory bowel disease (4.19 *10^−4^), Qualifications: None of the above (1.08 * 10^−3^), Triglycerides (3.17 * 10^−3^) and Sleeplessness/insomnia (4.35 * 10^−3^). In the opposite direction, we found psoriasis to be causal to Neuroticism (1.53 * 10^−4^), Frequency of depressed mood in last 2 weeks (1.19 * 10-4), Suffer from ‘nerves’ (2.02 * 10^−4^), Creatinine (enzymatic) in urine (2.41 * 10^−4^), Sodium in urine (1.71 * 10^−3^), Diagnoses - main ICD10: K57 Diverticular disease of intestine (1.94 * 10^−3^), Qualifications: Other professional qualifications eg: nursing, teaching (2.74 * 10^−3^), Qualifications: None of the above (3.03 * 10^−3^), Age completed full time education (3.38 * 10^−3^), Loneliness & isolation (3.25 * 10^−3^), Fed-up feelings (3.65 * 10^−3^), Qualifications: College or university degree (4.24 * 10^−3^), Diagnoses - main ICD10: R10 Abdominal and pelvic pain (5.41 * 10^−3^), Sensitivity / hurt feelings (5.30 * 10^−3^), Diagnoses - main ICD10: K80 Cholelithiasis (5.73 * 10^−3^), Neuroticism score (6.98 * 10^−3^), Apolipoprotein A-I (7.64 * 10^−3^) and Qualifications: A levels/AS levels (8.29 * 10^−3^).

**Figure 5A-B.**
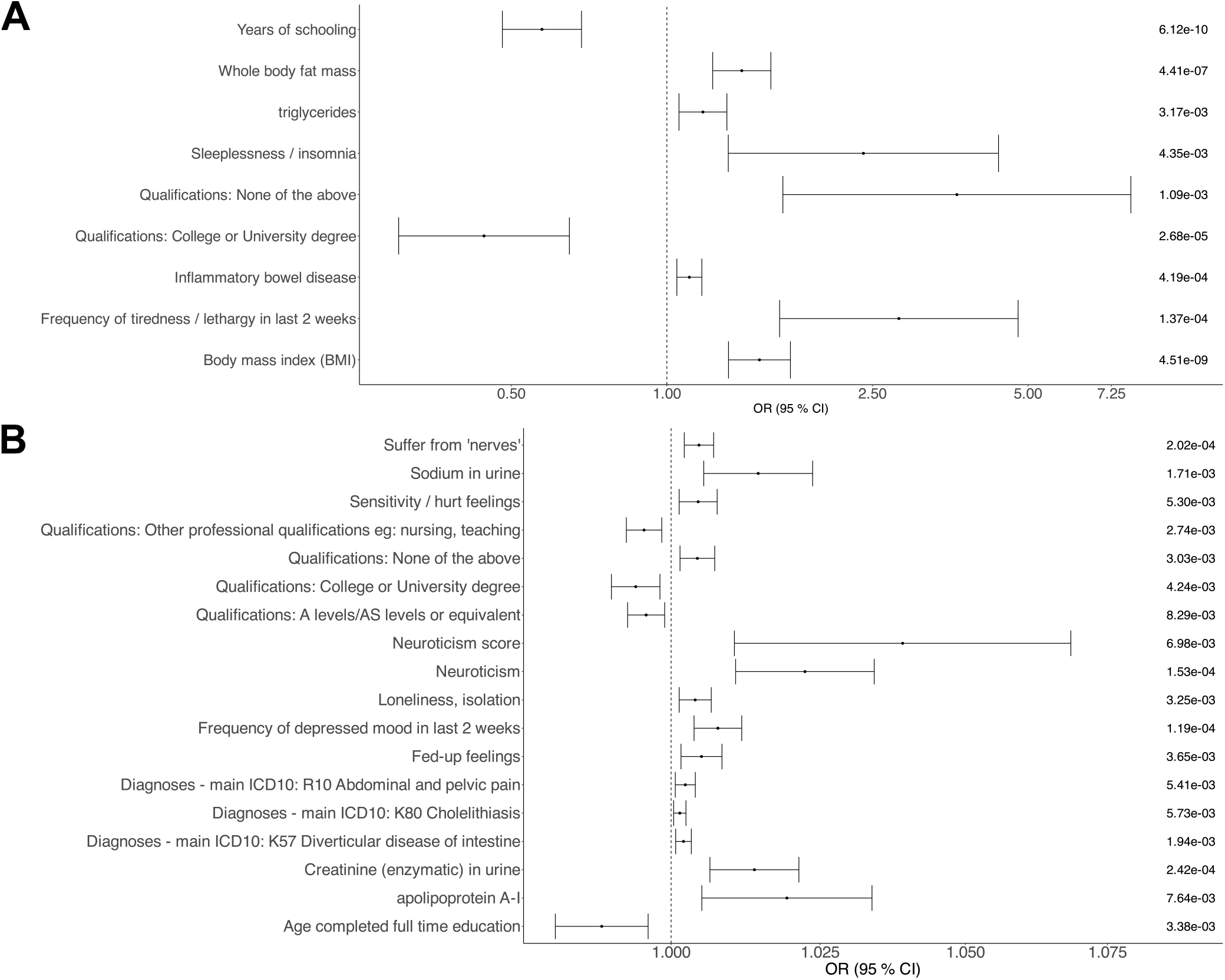
Forest plot of significant causal relationships between psoriasis and other traits. Mendelian randomization IVW estimates on significant causal relationships. Directions of causality: A) exposure to psoriasis, B) psoriasis to outcome. Odds ratios and 95 % confidence intervals on the x-axis, trait names on left side of the y-axis and p-values on the right side of the plot.

## Discussion

In this study, we discovered 8 novel psoriasis risk loci and replicated multiple previously reported loci. Based on careful manual curation, we suggest a potential biologically relevant candidate gene driving the associations at each novel locus. These genes encode proteins involved in either processes in the psoriatic skin or in the regulation of immune system. In addition, we report significant genetic correlations between psoriasis and 319 other traits. We further tested some of the correlated traits for causality and report evidence of causal relationship between psoriasis and mood-related symptoms and low education level.

One of the most prominent psoriasis pathogenesis features is hyperproliferation of keratinocytes^1^, the primary cell type found on epidermis. *FGFR4* is a fibroblast growth factor receptor, that is expressed mainly on lungs (**Supplementary Figure S3**). It acts as a receptor for fibroblast growth factor 19 (FGF19). It has been found to activate Wnt/-catenin pathway^25^. Through this pathway, high levels of FGF19 lead to elevated beta catenin binding with TCF4, which in turn leads to keratinocyte proliferation. *FGFR4*s role was also confirmed, as its inhibition removed FGF19’s promotional effect on pathway and keratinocyte proliferation. Thus, *FGFR4* has a significant role in process leading to one of the most significant characteristics of psoriatic skin.

Another known feature of psoriatic skin is excessive angiogenesis^29^, the formation of new blood vessels. This process is linked with the proliferation of keratinocytes in psoriasis, as many angiogenic factors, like vascular endothelial growth factor (VEGF)^36^ are upregulated by interleukins activated in psoriasis^29,37^. *ROBO1*, a roundabout homolog, has the highest expression level in fibroblasts, vagina, cervix, and skin (**Supplementary Figure S4**). It acts as a part of the Slit/Robo signaling pathway, which is included in a wide range of biological functions, including neuronal axon guidance, inflammatory cell chemotaxis and angiogenesis^28^. Angiogenesis activity has been found to be promoted through pathway including *ROBO1* and *VEGFR2*, VEGF receptor^38^. Thus, the found association with psoriasis could be linked *ROBO1*’s role in angiogenesis.

The rest of the candidate genes appear to be associated with immune system. The role of immunity in psoriasis is also highlighted by the FUMA gene set enrichment results (**Supplementary Figure S1**). The suggested roles of immune system related candidate genes are various. *SLC39A10*, coding zinc transporter ZIP10, expresses mostly in brain and thyroid (**Supplementary Figure S5**). It has been found modulate B-cell antigen receptor signal transduction^24^, and thus, adaptive immune response. Additionally, there’s evidence of ZIP10 expression in skin being higher in patients with atopic dermatitis, another inflammatory cutaneous disorder^23^. *COLEC10*, coding CL-L1, is a pattern recognition molecule and a member of the collectin family, expressing mainly in liver (**Supplementary Figure S6**). It is often found in circulation in complex with another collectin, *COLEC11* (CL-K1). Many collectins act in pathogen detection^30^, and *COLEC10* is also suggested to have a role in immune system. For example, its low serum level associates with infections in children^39^ and autoimmune disease systemic lupus erythematosus^31^. Additionally, *COLEC10* may affect skin immune homeostasis, as it might have a role in neural crest cell migration and therefore possibly affect melanocyte production^30^. *IL4R* is a receptor for interleukins 4 and 13^27^. Both are cytokines secreted by white blood cells and have functions concerning especially immune reaction of allergens or pathogens^40^. Additionally, both IL4 and IL13 are important in pathogenesis of atopic dermatitis^41^. The gene coding interleukin 4 has been previously found to associate with psoriasis in GWAS^5^, and the molecule has long been a target of medical treatments^42^, making its receptor a probable association. *IL4R* expresses most notably in lungs and blood (**Supplementary Figure S7**).

Candidate genes *LRRC25, TRAFD1* and *RNF216* function on the nuclear factor kappa-light-chain-enhancer of B cells (NF-κB) signaling pathway, which is known to have a major influence on psoriasis pathogenesis^4^. The NF-κB protein complex is known for its role in immune system responses. It controls for example DNA transcription, cytokine production and cell survival. Psoriatic skin contains greater levels of active NF-κB transcription factor than non-psoriatic skin^4,43^, and it takes part in psoriasis pathogenesis by altering keratinocyte and immune cell behavior. Thus, genes involved in this pathway could also have an association with psoriasis. *LRRC25*, is a leucine-rich repeat containing molecule expressing in blood (**Supplementary Figure S8**). Leucine-rich repeat containing molecules are typically involved in immune responses^44^, as is LRRC25, that has been found to inhibit NF-κB action through promoting autophagic degradation of p65/RelA, one of NF-κB’s subunits^34^. This happens through LRR subunit interacting with Rel homology domain in p65/RelA and LRRC25 enhancing the interaction between p65/RelA and p62, a cargo receptor. *TRAFD1* (also known as *FLN29*) is a negative feedback regulator gene involved in immune response^32^, mainly expressing in lymphocytes (**Supplementary Figure S9**). It inhibits toll-like receptor (TLR) 4 signaling and thereby also NF-κB^32,33^. TLRs are pathogen detectors and affect NF-κB action through several pathways^4^. TLR4, for example, detects and lipopolysaccharides found on the surface of bacteria^45^, which induces immune signaling and NF-κB activation. *RNF216* also has a role in TLR signaling. It acts as a E3 ubiquitin ligase for TLR4 and TLR9^46^. Ubiquitination is a process for marking proteins for degradation. Therefore, RNF216 controls the levels of TLR4 and TLR9, through which it affects NF-κB signaling^26,46^. The gene shows highest expression in testis (**Supplementary Figure S10**).

We report that altogether 319 phenotypes genetically correlated with psoriasis. These include multiple traits related to physical activity, anthropometry, other diseases, level of education, mood, pain, smoking, medication and blood metabolites. Out of the top traits, many, such as traits related to higher disease prevalence^47^, smoking^48^, high BMI^49^, lower level of education, and having children at a younger age^50^ are associated with lower socio-economic status, which has previously been suggested to lead to worse psoriasis symptoms^51^. Many of the phenotypes showing significant genetic correlation fit well with previous knowledge about psoriasis’ comorbidities. For example, we found multiple mood-related traits and for example major depressive disorder to be genetically correlated with psoriasis, and many mood disorders are known to be comorbid to psoriasis^52^.

We report 27 causal relationships between psoriasis and other traits. Especially high BMI and body fat mass show significant causal effect to psoriasis. This is in line with previous knowledge about psoriasis risk factors and two previous MR studies^53,54^ reporting causality between psoriasis and BMI. Additionally, our results confirm the previously detected^55^ causal relationship between inflammatory bowel disease (IBD) and psoriasis.

To the opposite direction, our results suggest psoriasis being causal to multiple mood-related symptoms, such as neuroticism, frequency of depressed mood or suffering from ‘nerves’. These results may suggest that psoriasis worsens certain mood-related symptoms, which may explain its known correlation with different psychiatric disorders. Multiple affective disorders have been previously associated with psoriasis in epidemiological settings, for example depression and anxiety are more common in psoriasis patients^56^. Common mechanisms have been detected between affective disorders and psoriasis; for example, the same pro-inflammatory cytokines levels are increased^57^ and hypothalamic-pituitary-adrenal (HPA) axis is dysregulated^58^ in both depression and psoriasis. Our results suggest that psoriasis may be causal to symptoms that may partly explain the high prevalence of psychiatric disorders in psoriasis patients, as these symptoms may be signs of multiple different psychiatric disorders. For example, both neuroticism^59^ and loneliness^60^ are associated with risk of mood and anxiety disorders. Additionally, we found multiple diagnoses psoriasis is causal to: diverticular disease of intestine, abdominal and pelvic pain and cholelithiasis. There is some previous evidence possibly suggesting a connection between diverticular disease of intestine and cholelithiasis with psoriasis: risk for developing diverticulitis may be a bit greater for psoriasis patients^61^, and incidence of gallstones has been found to be associated with psoriasis and psoriatic arthritis, independent of obesity^62^. Such evidence was not found for abdominal and pelvic pain, perhaps suggesting its correlation being linked to one or multiple other diagnoses as a symptom.

Previous work has shown the epidemiological association between lower socio-economic status and worse psoriasis symptoms^51^. Here, our results show that education-related traits form a noticeable amount of significant causal relationships to both directions. Thus, these results seem to suggest a complex relationship between psoriasis and lower education level without a definite direction for causality.

### Limitations

The study populations differed in the recognition of psoriasis in patients. The sample prevalence of psoriasis was 6,9 % in EstBB, 2,3 % in FinnGen and 0,69 % in UKBB. Possible non-identified psoriasis patients as controls may decrease statistical power to detect associated loci, and the differences in psoriasis prevalence might increase the heterogeneity in results. Additionally, the positive predictive value for psoriasis is 88.0 % in Finnish biobank registries^63^ meaning uncertain psoriasis cases may influence the accuracy of results. Some of the novel loci found in the discovery GWAS appeared to be specific to the Finnish population and thus the replication of these loci in further studies would be of high value. We did not consider psoriasis subtypes in our analysis. Subtypes have been previously suggested to have slightly different genetic backgrounds^1^. Thus, the analysis of the subtypes could have shed light on the exact associations and differences between subgroups, that now may remain uncharacterized. Considering MR results, it should be noted that we detected significant heterogeneity in multiple significant traits (**Supplementary Table S5-6**), and results should be interpreted with care.

### Conclusion

To conclude, we found 8 novel psoriasis risk loci, and, overall, the present genome-wide analyses emphasized the role of immune-regulating genes in the pathogenesis of psoriasis. The results of the genetic correlations provided genetic evidence for the close link between psoriasis and socio-economic status. With Mendelian randomization we found evidence of causal relationships between obesity and psoriasis, psoriasis and mood symptoms, and complex causal relationships between psoriasis and low education level. Our findings deepen understanding of psoriasis and may be useful in treatment development.

## Data Availability

Summary statistics will be made available through the NHGRI-EBI GWAS Catalog with GCSTxxxxxxx upon publication. The Finnish biobank data can be accessed through the Fingenious® services (https://site.fingenious.fi/en/) managed by FINBB.

## Acknowledgements

We want to acknowledge the participants and investigators of FinnGen study. Following biobanks are acknowledged for delivering biobank samples to FinnGen: Auria Biobank (www.auria.fi/biopankki), THL Biobank (www.thl.fi/biobank), Helsinki Biobank (www.helsinginbiopankki.fi), Biobank Borealis of Northern Finland (https://www.ppshp.fi/Tutkimus-ja-opetus/Biopankki/Pages/Biobank-Borealis-briefly-in-English.aspx), Finnish Clinical Biobank Tampere (www.tays.fi/en-US/Research_and_development/Finnish_Clinical_Biobank_Tampere), Biobank of Eastern Finland (www.ita-suomenbiopankki.fi/en), Central Finland Biobank (www.ksshp.fi/fi-FI/Potilaalle/Biopankki), Finnish Red Cross Blood Service Biobank (www.veripalvelu.fi/verenluovutus/biopankkitoiminta), Terveystalo Biobank (www.terveystalo.com/fi/Yritystietoa/Terveystalo-Biopankki/Biopankki/) and Arctic Biobank (https://www.oulu.fi/en/university/faculties-and-units/faculty-medicine/northern-finland-birth-cohorts-and-arctic-biobank). All Finnish Biobanks are members of BBMRI.fi infrastructure (www.bbmri.fi). Finnish Biobank Cooperative -FINBB (https://finbb.fi/) is the coordinator of BBMRI-ERIC operations in Finland.

Estonian Biobank Research Team members: Andres Metspalu, Mari Nelis, Lili Milani, Reedik Mägi & Tõnu Esko

