## Supplementary Material for "Genetic study of psoriasis highlights its close link with socio-economic status and affective symptoms"

**Supplementary Note. FinnGen DF7 ethics statement.**

FinnGen participants provided written informed consent for biobank research, based on the Finnish Biobank Act. Alternatively, separate research cohorts, collected prior the Finnish Biobank Act came into effect (in September 2013) and start of FinnGen (August 2017), were collected based on study-specific written consents and later transferred to the Finnish biobanks after approval by Fimea (Finnish Medicines Agency), the National Supervisory Authority for Welfare and Health. Recruitment protocols followed the biobank protocols approved by Fimea. The Coordinating Ethics Committee of the Hospital District of Helsinki and Uusimaa (HUS) statement number for the FinnGen study is Nr HUS/990/2017.

The FinnGen study is approved by Finnish Institute for Health and Welfare (permit numbers: THL/2031/6.02.00/2017, THL/1101/5.05.00/2017, THL/341/6.02.00/2018, THL/2222/6.02.00/2018, THL/283/6.02.00/2019, THL/1721/5.05.00/2019, THL/1524/5.05.00/2020, and THL/2364/14.02/2020), Digital and population data service agency (permit numbers: VRK43431/2017-3, VRK/6909/2018-3, VRK/4415/2019-3), the Social Insurance Institution (permit numbers: KELA 58/522/2017, KELA 131/522/2018, KELA 70/522/2019, KELA 98/522/2019, KELA 138/522/2019, KELA 2/522/2020, KELA 16/522/2020, Findata THL/2364/14.02/2020 and Statistics Finland (permit numbers: TK-53-1041-17 and TK/143/07.03.00/2020 (earlier TK-53-90-20).

The Biobank Access Decisions for FinnGen samples and data utilized in FinnGen Data Freeze 7 include: THL Biobank BB2017_55, BB2017_111, BB2018_19, BB_2018_34, BB_2018_67, BB2018_71, BB2019_7, BB2019_8, BB2019_26, BB2020_1, Finnish Red Cross Blood Service Biobank 7.12.2017, Helsinki Biobank HUS/359/2017, Auria Biobank AB17-5154 and amendment #1 (August 17 2020), Biobank Borealis of Northern Finland_2017_1013, Biobank of Eastern Finland 1186/2018 and amendment 22 § /2020, Finnish Clinical Biobank Tampere MH0004 and amendments (21.02.2020 & 06.10.2020), Central Finland Biobank 1-2017, and Terveystalo Biobank STB 2018001.

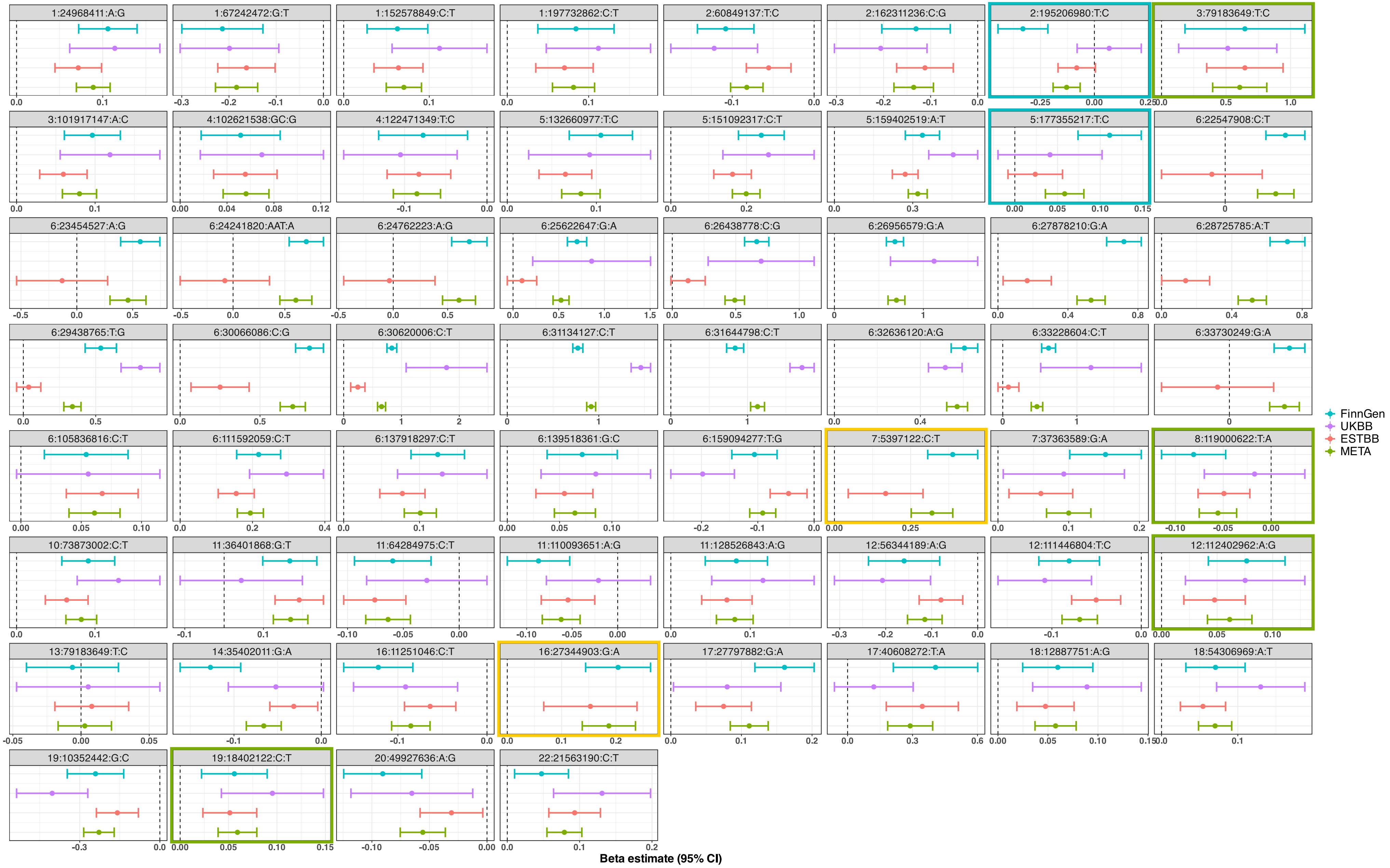
1:24968411:A:G
1:67242472:G:T
1:152578849:C:T
1:197732862:C:T
2:60849137:T:C
2:162311236:C:G
2:195206980:T:C
3:79183649:T:C
3:101917147:A:C
4:102621538:GC:G
4:122471349:T:C
5:132660977:T:C
5:151092317:C:T
5:159402519:A:T
5:177355217:T:C
6:22547908:C:T
6:23454527:A:G
6:24241820:AAT:A
6:24762223:A:G
6:25622647:G:A
6:26438778:C:G
6:26956579:G:A
6:27878210:G:A
6:28725785:A:T
6:29438765:T:G
6:30066086:C:G
6:30620006:C:T
6:31134127:C:T
6:31644798:C:T
6:32636120:A:G
6:33228604:C:T
6:33730249:G:A
6:105836816:C:T
6:111592059:C:T
6:137918297:C:T
6:139518361:G:C
6:159094277:T:G
7:5397122:C:T
7:37363589:G:A
8:119000622:T:A
10:73873002:C:T
11:36401868:G:T
11:64284975:C:T
11:110093651:A:G
11:128526843:A:G
12:56344189:A:G
12:111446804:T:C
12:112402962:A:G
13:79183649:T:C
14:35402011:G:A
16:11251046:C:T
16:27344903:G:A
17:27797882:G:A
17:40608272:T:A
18:12887751:A:G
18:54306969:A:T
19:10352442:G:C
19:18402122:C:T
20:49927636:A:G
22:21563190:C:T
FinnGen
UKBB
ESTBB
META
Beta estimate (95% CI)

**Figure S1. Forest plot showing effect estimates of meta-analysis lead variants and novel FinnGen lead variants in each study population (FinnGen, Estonian Biobank and UK Biobank) and meta-analysis.**
Every lead variant and its beta effect estimate (whiskers showing 95 % confidence interval) in each study population are shown in boxes headlined with variant position information. Populations from top down in each box: FinnGen (blue), UKBB (purple), ESTBB (red) and META (green). Boxes including novel lead variants are marked with colored frame based on population in which they are significant: variants marked with blue frame are significant only in FinnGen population (2:195206980:T:C and 5:177355217:T:C), with green only in meta-analysis (3:79183649:T:C, 8:119000622:T:A, 12:112402962:A:G and 19:18042122:C:T), and with yellow significant in both FinnGen and meta-analysis (7:5397122:C:T and 16:27344903:G:A).

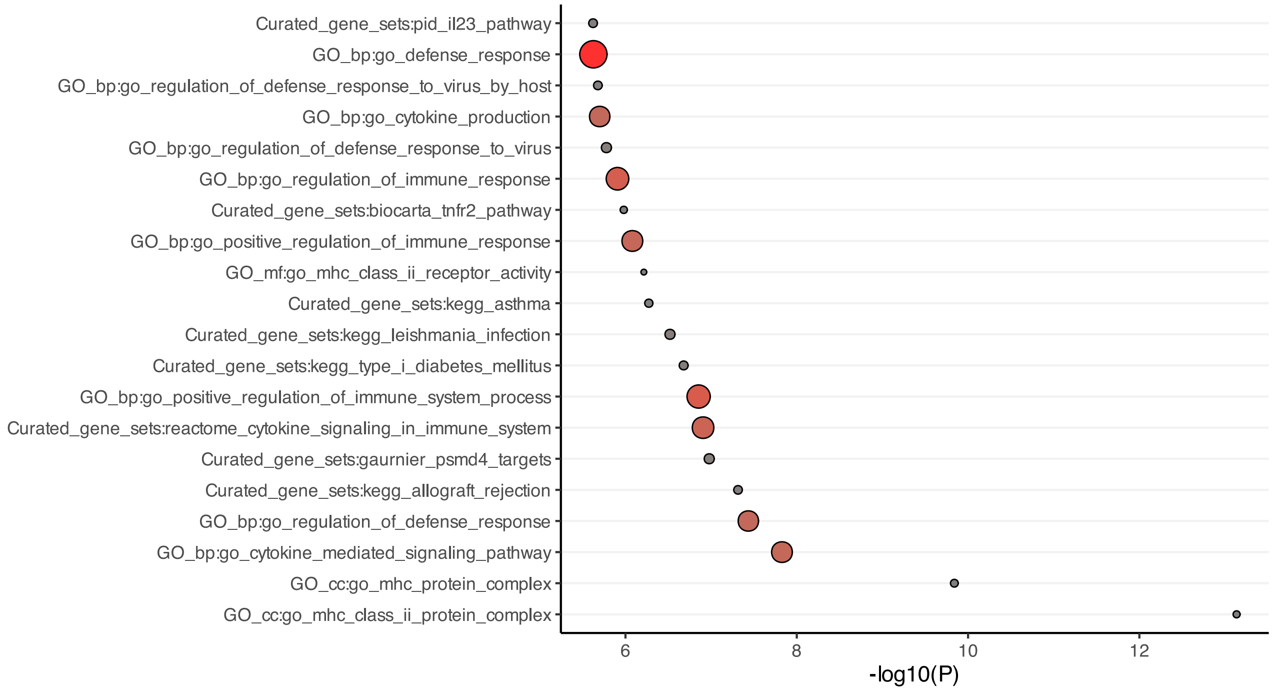

**Figure S2. MAGMA (Multi-marker Analysis of GenoMic Annotation) gene set enrichment analysis plot.**
The analysis was run in Functional Mapping and Annotation of Genome-Wide Association Studies (FUMA) pipeline. The names of the gene sets are listed on the y-axis and -log10 p-values of the enrichments are shown on x-axis. The size and color of the dot symbolize the size of the gene set: gene set smaller than 400 genes is marked with grey small dot, 400-800 genes with rust-colored medium-sized dot and greater than 800 genes with big bright red dot.

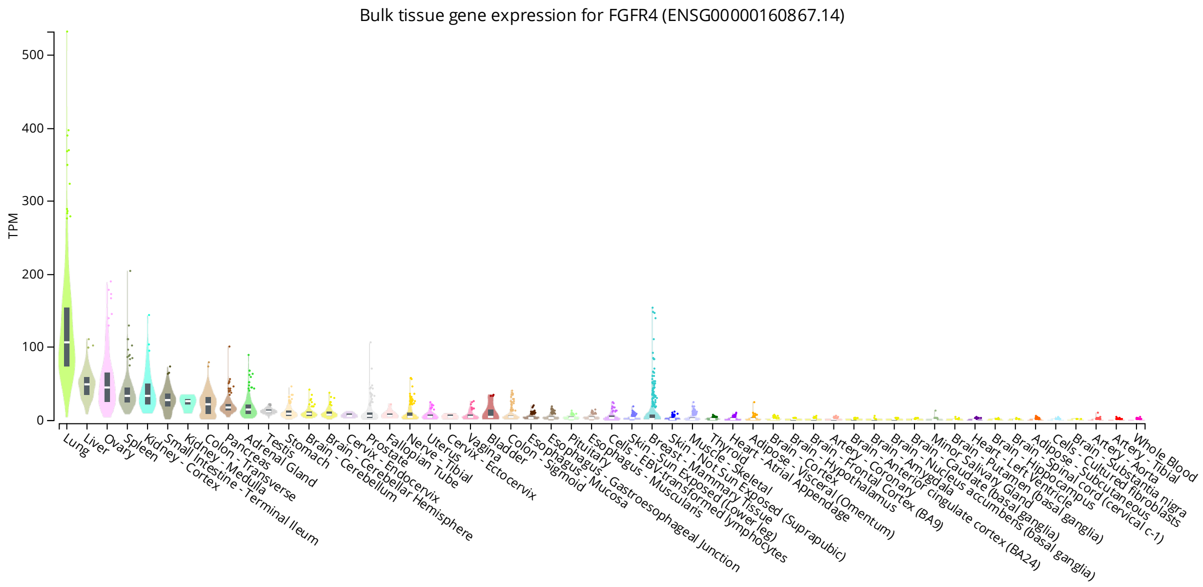

**Figure S3. *FGFR4* expression in tissues.**
Extracted from GTEx Portal 20.1.2023.

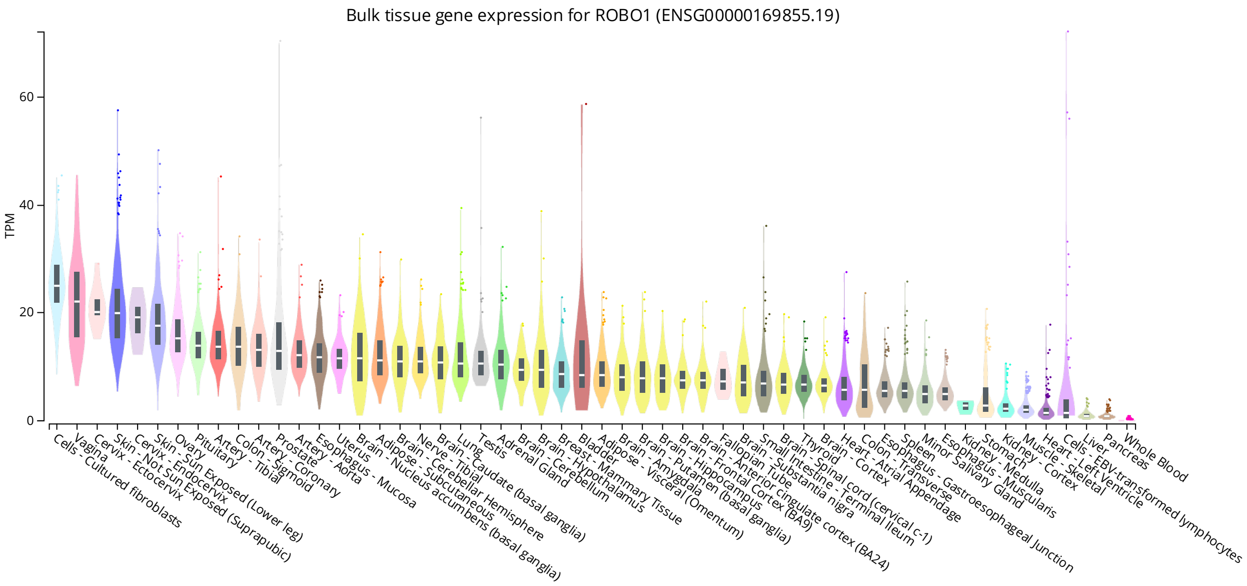

**Figure S4. *ROBO1* expression in tissues.**
Extracted from GTEx Portal 20.1.2023.

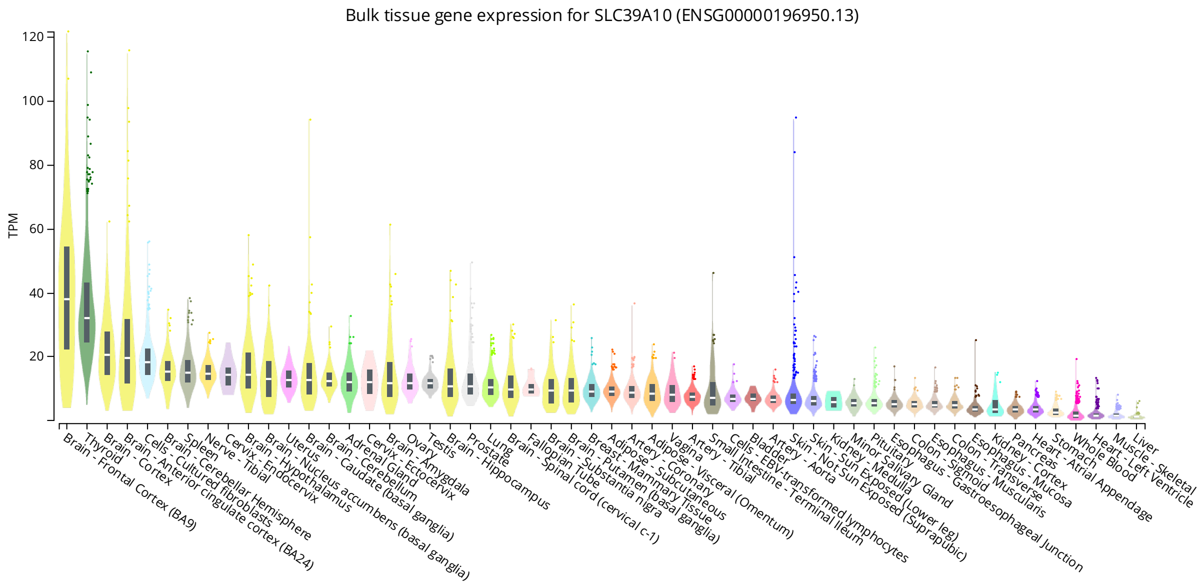

**Figure S5. *SLC39A10* expression in tissues.**
Extracted from GTEx Portal 20.1.2023.

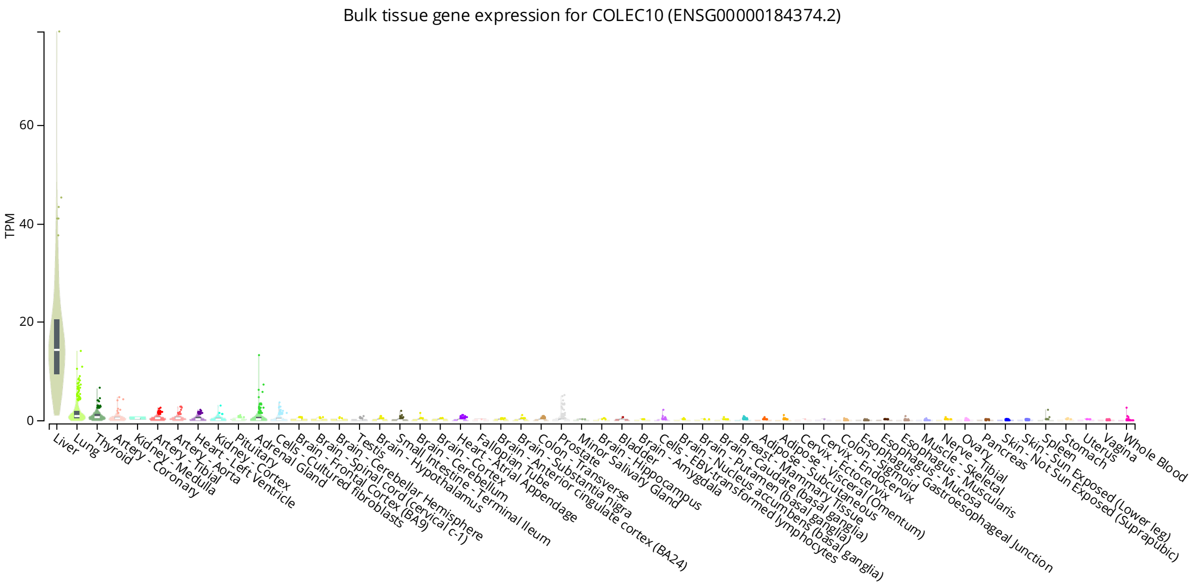

**Figure S6. *COLEC10* expression in tissues.**
Extracted from GTEx Portal 20.1.2023.

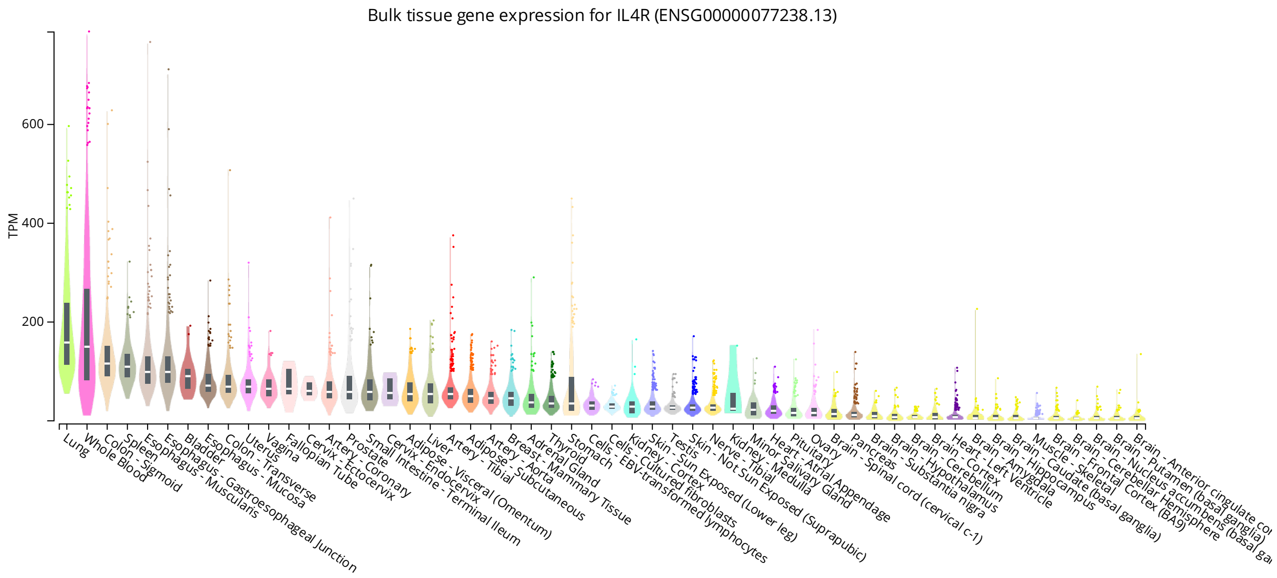

**Figure S7. *IL4R* expression in tissues.**
Extracted from GTEx Portal 20.1.2023.

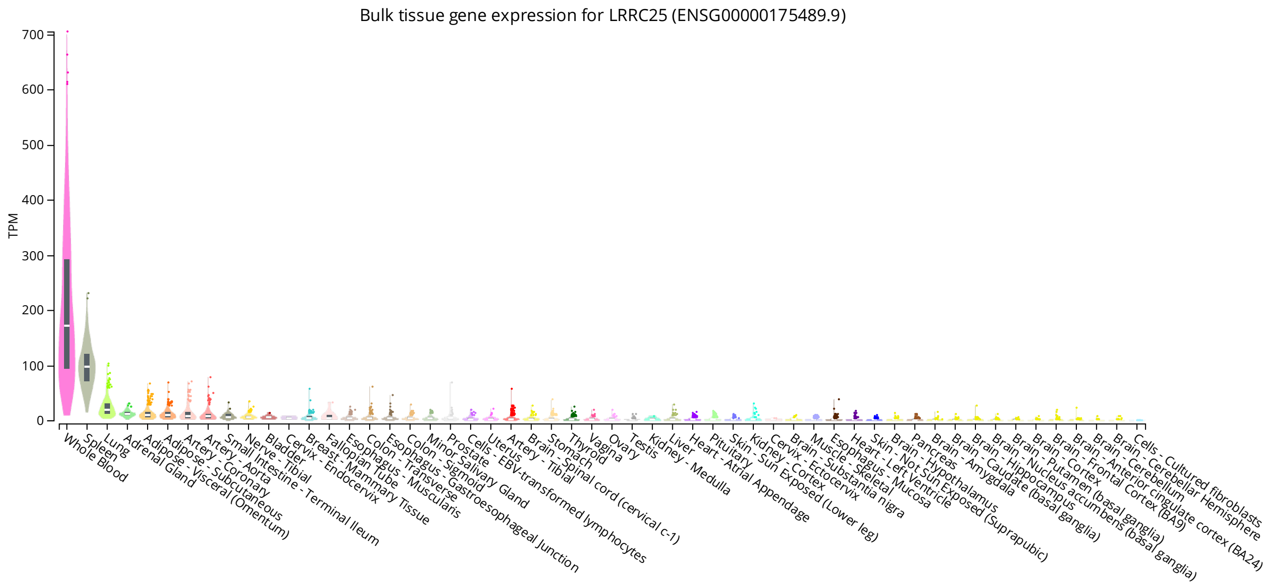

**Figure S8. *LRRC25* expression in tissues.**
Extracted from GTEx Portal 20.1.2023.

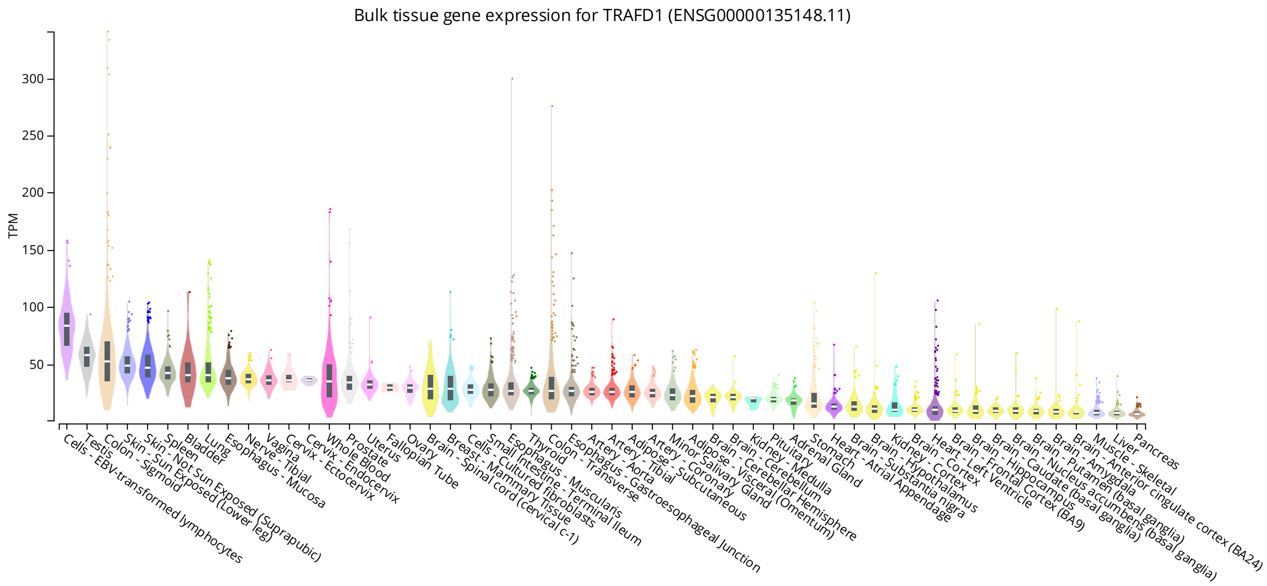

**Figure S9. *TRAFD1* expression in tissues.**
Extracted from GTEx Portal 20.1.2023.

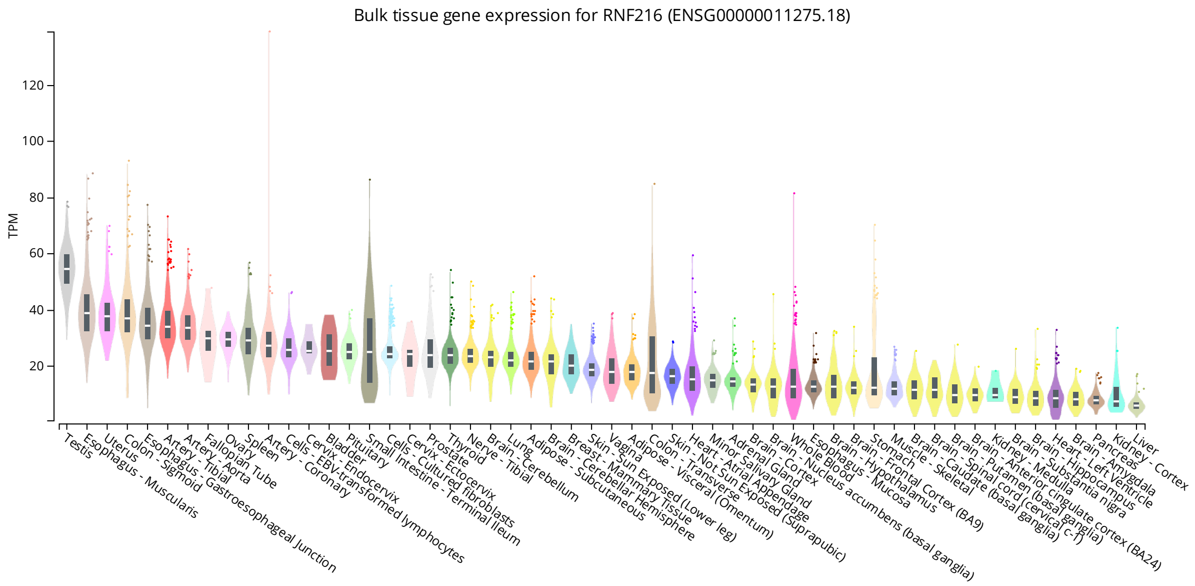

**Figure S10.** ***RNF216*** **expression in tissues.**
Extracted from GTEx Portal 20.1.2023.

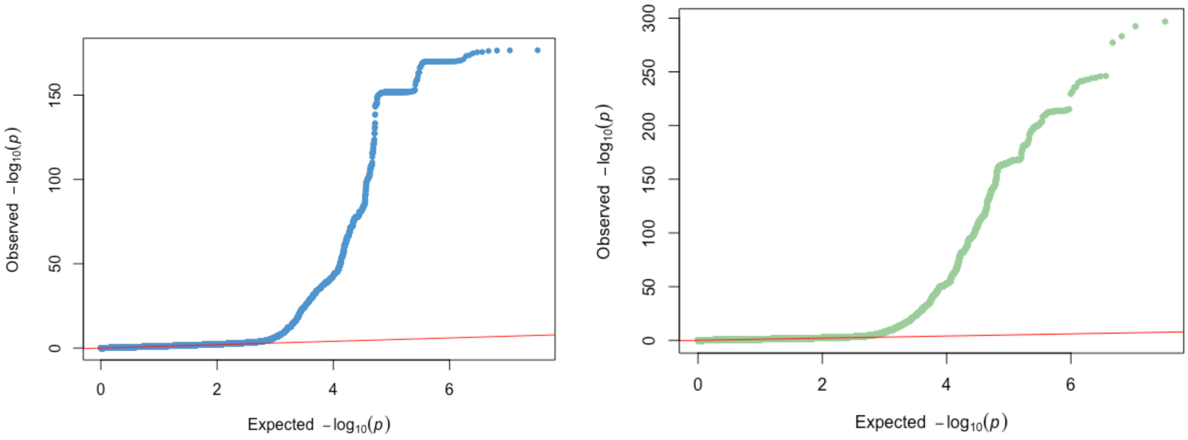

**Figure S11. Quantile-quantile (QQ) plots of p-values.**
The plot on the left with blue dots shows p-values in FinnGen discovery analysis and the plot on the right with green dots in meta-analysis in FinnGen, EstBB and UKBB.

**Table S1. Genome-wide significant loci (p < 5.10E-8) in discovery GWAS in FinnGen.** Columns in table: chr:pos, chromosome and position; rsid, rs-number; ref, reference allele; alt, alternative allele (effect allele); OR, odds ratio; 95 % CI, 95 % confidence interval; p, p-value; af _alt, alternative allele frequency; Fin enr., Finnish enrichment.

| locus | nearest gene | chr:pos | rsid | ref | alt | OR | 95 % CI | p | af_alt | Fin enr. |
|---|---|---|---|---|---|---|---|---|---|---|
| 1q12 | RUNX3 | 1:24968116 | rs7542079 | T | C | 1.11 | 1.08-1.15 | 6.05E-10 | 0.5622 | 1.136 |
| 2p16.1 | REL | 2:60890978 | rs12713428 | A | C | 1.15 | 1.11-1.19 | 7.16E-13 | 0.2485 | 1.115 |
| **2q32.3** | **SLC39A10** | **2:195206980** | **rs77509633** | **T** | **C** | **0.72** | **0.64-0.81** | **1.93E-08** | **0.0262** | **0.644** |
| 3q12.3 | NFKBIZ | 3:101937583 | rs771571 | G | C | 0.91 | 0.88-0.94 | 2.79E-08 | 0.431 | 0.698 |
| 5q31.1 | IL13 | 5:132660977 | rs847 | T | C | 1.11 | 1.07-1.15 | 6.00E-09 | 0.6352 | 0.762 |
| 5q33.1 | ANXA6 | 5:151098757 | rs17728338 | G | A | 1.27 | 1.2-1.35 | 8.21E-15 | 0.0739 | 1.274 |
| 5q33.3 | IL12B | 5:159422483 | rs10866712 | T | C | 0.84 | 0.81-0.86 | 1.40E-24 | 0.6432 | 0.935 |
| **5q35.3** | **RGS14** | **5:177355217** | **rs13153019** | **T** | **C** | **1.12** | **1.08-1.16** | **4.36E-09** | **0.2723** | **1.028** |
| 6p22.3 | HDGFL1 | 6:22547908 | rs190317150 | C | T | 1.69 | 1.43-2 | 1.35E-09 | 0.0076 | NA |
| 6p22.3 | NRSN1 | 6:23454527 | rs190749544 | A | G | 1.76 | 1.48-2.1 | 2.37E-10 | 0.0071 | NA |
| 6p22.3 | DCDC2 | 6:24241820 | rs555763495 | AAT | A | 2.03 | 1.72-2.39 | 5.34E-17 | 0.0066 | NA |
| 6p22.3 | GMNN | 6:24762223 | rs553157269 | A | G | 2.01 | 1.71-2.37 | 3.08E-17 | 0.0071 | NA |
| 6p22.2 | LRRC16A | 6:25622647 | rs151212550 | G | A | 2.01 | 1.81-2.24 | 1.35E-38 | 0.0166 | 6.585 |
| 6p22.2 | BTN3A1 | 6:26401603 | rs140614283 | G | A | 1.93 | 1.75-2.12 | 1.84E-42 | 0.0214 | 3.311 |
| 6p22.2 | HIST1H2BJ | 6:26937601 | rs145899165 | T | C | 1.97 | 1.79-2.18 | 1.95E-43 | 0.02 | 9.503 |
| 6p22.1 | HIST1H4L | 6:27878210 | rs184863525 | G | A | 2.05 | 1.86-2.27 | 3.20E-46 | 0.0189 | 8.321 |
| 6p22.1 | GPX6 | 6:28517139 | rs147333691 | G | A | 2.05 | 1.86-2.26 | 5.83E-46 | 0.0196 | 6.779 |
| 6p22.1 | OR2B3 | 6:29074437 | rs138374168 | T | G | 2.04 | 1.85-2.25 | 2.38E-44 | 0.0194 | 7.107 |
| 6p22.1 | GABBR1 | 6:29575029 | rs28986295 | T | A | 1.63 | 1.48-1.8 | 5.73E-23 | 0.0222 | 0.396 |
| 6p22.1 | RNF39 | 6:30079072 | rs142938566 | G | A | 2.15 | 1.98-2.32 | 1.63E-78 | 0.0286 | 4.014 |
| 6p21.33 | DHX16 | 6:30665484 | rs34505140 | C | T | 2.3 | 2.11-2.5 | 1.11E-82 | 0.0235 | 9.153 |
| 6p21.33 | HLA-C | 6:31299200 | rs13210419 | G | A | 2.24 | 2.12-2.37 | 2.28E-177 | 0.059 | 0.811 |
| 6p21.33 | TNXB | 6:32046679 | rs34214527 | C | T | 1.99 | 1.87-2.11 | 8.05E-111 | 0.0541 | 0.693 |
| 6p21.32 | HLA-DQA1 | 6:32638048 | rs28383348 | G | A | 1.82 | 1.71-1.93 | 2.82E-82 | 0.0569 | NA |
| 6p21.32 | RING1 | 6:33228604 | rs186014442 | C | T | 1.85 | 1.69-2.04 | 3.28E-38 | 0.0225 | 8.207 |

| locus | nearest gene | chr:pos | rsid | ref | alt | OR | 95 % CI | p | af_alt | Fin enr. |
|---|---|---|---|---|---|---|---|---|---|---|
| 6p21.31 | MLN | 6:33908113 | rs143909713 | G | C | 1.49 | 1.35-1.65 | 1.55E-14 | 0.0226 | 9.483 |
| 6p21.31 | UHRF1BP1 | 6:34852120 | rs1052297940 | A | G | 1.76 | 1.44-2.15 | 3.40E-08 | 0.0051 | NA |
| 6p21.31 | TULP1 | 6:35543106 | rs1009323567 | C | A | 1.77 | 1.45-2.16 | 2.76E-08 | 0.0049 | NA |
| 6q21 | TRAF3IP2 | 6:111592059 | rs33980500 | C | T | 1.24 | 1.17-1.32 | 2.94E-12 | 0.0726 | 0.98 |
| 6q23.3 | TNFAIP3 | 6:137895651 | rs674451 | T | C | 1.13 | 1.09-1.17 | 4.06E-12 | 0.3439 | 1.051 |
| **7p22.1** | **TNRC18** | **7:5397122** | **rs748670681** | **C** | **T** | **1.47** | **1.36-1.6** | **1.53E-20** | **0.0359** | **114.849** |
| 7p14.1 | ELMO1 | 7:37342861 | rs60600003 | T | G | 1.18 | 1.12-1.25 | 9.02E-10 | 0.1019 | 0.967 |
| 14q13.2 | NFKBIA | 14:35402011 | rs8904 | G | A | 0.88 | 0.85-0.91 | 8.89E-13 | 0.388 | 1.064 |
| 16p13.13 | RMI2 | 16:11251046 | rs2021511 | C | T | 0.89 | 0.85-0.92 | 7.14E-10 | 0.266 | 0.973 |
| **16p12.1** | **IL4R** | **16:27344903** | **rs144651842** | **G** | **A** | **1.23** | **1.16-1.3** | **2.19E-11** | **0.078** | **119.053** |
| 17q11.2 | NOS2 | 17:27797882 | rs28998802 | G | A | 1.18 | 1.13-1.23 | 4.87E-14 | 0.1849 | 1.359 |
| 20q13.13 | SPATA2 | 20:49907751 | rs636987 | A | C | 0.91 | 0.88-0.94 | 2.22E-08 | 0.618 | 1.343 |
| 22q13.31 | NUP50 | 22:45186583 | rs140716375 | A | G | 0.65 | 0.56-0.76 | 3.57E-08 | 0.0168 | 15.413 |

**Table S2. Genome-wide significant loci (p < 5.10E-8) in meta-analysis (FinnGen, EstBB, UKBB).**
Columns in table: chr:pos, chromosome and position; rsid, rs-number; ref, reference allele; alt, alternative allele (effect allele); OR, odds ratio; CI 95 %, 95 % confidence interval; p, p-value; het_p, heterogeneity between study populations p-value (Cochran's Q-test); af _alt_FinnGen, alternative allele frequency in FinnGen; af_alt_EstBB, alternative allele frequency in Estonian Biobank; Fin enr., Finnish enrichment.

| locus | nearest gene | chr:pos | rsid | ref | alt | OR | CI 95 % | p | het_p | af_alt FinnGen | af_alt EstBB | Fin enr. |
|---|---|---|---|---|---|---|---|---|---|---|---|---|
| 1p36.11 | RUNX3 | 1:24968411 | rs10903119 | A | G | 1.09 | 1.07-1.11 | 5.29E-19 | 1.77E-01 | 0.5512 | 0.5083 | 1.128 |
| 1p31.3 | IL23R | 1:67242472 | rs80174646 | G | T | 0.83 | 0.8-0.87 | 8.90E-16 | 6.12E-01 | 0.0456 | 0.054 | 0.848 |
| 1q21.3 | LCE3D | 1:152578849 | rs4112787 | C | T | 1.07 | 1.05-1.1 | 2.56E-11 | 2.79E-01 | 0.6568 | 0.6461 | 1.019 |
| 1q31.3 | DENND1B | 1:197732862 | rs16841904 | C | T | 1.08 | 1.06-1.11 | 6.08E-10 | 5.26E-01 | 0.1464 | 0.1771 | 0.687 |
| 2p16.1 | REL | 2:60849137 | rs1306395 | T | C | 0.92 | 0.9-0.94 | 4.77E-16 | 1.82E-02 | 0.4122 | 0.4317 | 0.935 |
| 2q24.2 | IFIH1 | 2:162311236 | rs17715343 | C | G | 0.87 | 0.84-0.91 | 2.40E-10 | 2.75E-01 | 0.0607 | 0.0574 | 0.655 |
| **3p12.3** | **ROBO1** | **3:79183649** | **rs540696640** | **T** | **C** | **1.83** | **1.49-2.26** | **1.35E-08** | **8.50E-01** | **0.001** | **0.0021** | **0.052** |
| 3q12.3 | NFKBIZ | 3:101917147 | rs72939372 | A | C | 1.08 | 1.06-1.11 | 4.65E-13 | 1.36E-01 | 0.3176 | 0.2801 | 1.456 |
| 4q27 | NFKB1 | 4:122471349 | rs45545833 | T | C | 0.92 | 0.89-0.95 | 5.37E-09 | 8.13E-01 | 0.4585 | 0.4859 | 0.692 |
| 4q24 | IL2 | 4:102621538 | rs57463114 | GC | G | 1.06 | 1.04-1.08 | 1.78E-08 | 8.50E-01 | 0.1144 | 0.1434 | 0.883 |
| 5q31.1 | IL13 | 5:132660977 | rs847 | T | C | 1.09 | 1.06-1.11 | 5.29E-14 | 2.27E-01 | 0.6352 | 0.6772 | 0.762 |
| 5q33.1 | TNIP1 | 5:151092317 | rs146571698 | C | T | 1.22 | 1.18-1.27 | 1.16E-26 | 9.69E-02 | 0.0739 | 0.081 | 1.259 |
| 5q33.3 | IL12B | 5:159402519 | rs12188300 | A | T | 1.38 | 1.33-1.43 | 1.93E-66 | 2.38E-03 | 0.0591 | 0.0869 | 0.621 |
| 6p22.3 | HDGFL1 | 6:22547908 | rs190317150 | C | T | 1.55 | 1.33-1.82 | 4.58E-08 | 7.60E-03 | 0.0076 | 0.0012 | NA |
| 6p22.3 | NRSN1 | 6:23454527 | rs190749544 | A | G | 1.58 | 1.35-1.86 | 2.61E-08 | 1.99E-03 | 0.0071 | 0.0013 | NA |
| 6p22.3 | DCDC2 | 6:24241820 | rs555763495 | AAT | A | 1.83 | 1.57-2.14 | 1.45E-14 | 8.31E-04 | 0.0066 | 0.0008 | NA |
| 6p22.3 | GMNN | 6:24762223 | rs553157269 | A | G | 1.83 | 1.57-2.13 | 5.54E-15 | 1.36E-03 | 0.0071 | 0.0009 | NA |
| 6p22.2 | LRRC16A | 6:25622647 | rs151212550 | G | A | 1.69 | 1.55-1.84 | 6.06E-32 | 3.99E-09 | 0.0166 | 0.0052 | 6.585 |
| 6p22.2 | BTN3A3 | 6:26438778 | rs183691101 | C | G | 1.64 | 1.51-1.77 | 4.10E-36 | 8.00E-10 | 0.0205 | 0.0074 | 4.594 |
| 6p22.2 | HIST1H2BJ | 6:26956579 | rs555043772 | G | A | 2.01 | 1.83-2.21 | 3.93E-47 | 8.75E-02 | 0.02 | NA | 9.335 |
| 6p22.1 | HIST1H4L | 6:27878210 | rs184863525 | G | A | 1.7 | 1.57-1.84 | 1.63E-38 | 1.58E-10 | 0.0189 | 0.0072 | 8.321 |
| 6p22.1 | ZBED9 | 6:28725785 | rs193201625 | A | T | 1.68 | 1.55-1.82 | 1.55E-36 | 1.66E-11 | 0.0197 | 0.0075 | 6.995 |
| 6p22.1 | OR11A1 | 6:29438765 | rs55969931 | T | G | 1.4 | 1.32-1.49 | 4.48E-29 | 1.29E-24 | 0.0175 | 0.0207 | 0.364 |

| locus | nearest gene | chr:pos | rsid | ref | alt | OR | CI 95 % | p | het_p | af_alt FinnGen | af_alt EstBB | Fin enr. |
|---|---|---|---|---|---|---|---|---|---|---|---|---|
| 6p22.1 | PPP1R11 | 6:30066086 | rs187629177 | C | G | 2.02 | 1.87-2.19 | 4.85E-69 | 5.59E-08 | 0.0232 | 0.005 | 249.63 |
| 6p21.33 | MRPS18B | 6:30620006 | rs13212442 | C | T | 1.93 | 1.8-2.06 | 4.66E-76 | 7.31E-16 | 0.0235 | 0.0081 | 8.944 |
| 6p21.33 | PSORS1C1 | 6:31134127 | rs13212902 | C | T | 2.5 | 2.38-2.63 | 1.52E-297 | 1.01E-29 | 0.0629 | NA | 0.656 |
| 6p21.33 | BAG6 | 6:31644798 | rs28732155 | C | T | 3.1 | 2.83-3.39 | 5.83E-131 | 7.63E-19 | 0.0134 | NA | NA |
| 6p21.32 | HLA-DQA1 | 6:32636120 | rs28383338 | A | G | 1.77 | 1.69-1.86 | 7.37E-118 | 7.83E-02 | 0.0551 | NA | NA |
| 6p21.32 | RING1 | 6:33228604 | rs186014442 | C | T | 1.59 | 1.47-1.71 | 1.86E-31 | 3.10E-10 | 0.0225 | 0.0067 | 8.207 |
| 6p21.31 | IP6K3 | 6:33730249 | rs1023433775 | G | A | 1.94 | 1.62-2.32 | 3.50E-13 | 1.55E-02 | 0.0073 | 0.0006 | 81.222 |
| 6p21.31 | ATG5 | 6:105836816 | rs60411028 | C | T | 1.06 | 1.04-1.09 | 1.53E-08 | 8.35E-01 | 0.3833 | 0.2917 | 1.542 |
| 6q21 | TRAF3IP2 | 6:111592059 | rs33980500 | C | T | 1.21 | 1.17-1.26 | 4.62E-26 | 3.85E-02 | 0.0726 | 0.0785 | 0.98 |
| 6q23.3 | TNFAIP3 | 6:137918297 | rs6933987 | C | T | 1.11 | 1.08-1.13 | 1.23E-20 | 8.59E-02 | 0.332 | 0.2863 | 1.249 |
| 6q24.1 | CITED2 | 6:139518361 | rs608736 | G | C | 1.07 | 1.05-1.09 | 1.05E-10 | 5.34E-01 | 0.4788 | 0.5387 | 0.861 |
| 6q25.3 | TAGAP | 6:159094277 | rs2249937 | T | G | 0.91 | 0.89-0.93 | 1.13E-14 | 1.92E-05 | 0.7814 | 0.7704 | 1.168 |
| **7p22.1** | **TNRC18** | **7:5397122** | **rs748670681** | **C** | **T** | **1.38** | **1.29-1.47** | **3.13E-20** | **3.41E-03** | **0.0359** | **0.0125** | **114.85** |
| 7p14.1 | ELMO1 | 7:37363589 | rs77801025 | G | A | 1.11 | 1.07-1.14 | 3.64E-10 | 2.93E-02 | 0.1155 | 0.0988 | 1.003 |
| **8q24.12** | **COLEC10** | **8:119000622** | **rs13264172** | **T** | **A** | **0.95** | **0.93-0.96** | **2.46E-08** | **1.08E-01** | **0.5309** | **0.5279** | **0.988** |
| 10q22.2 | CAMK2G | 10:73873002 | rs2675671 | C | T | 1.09 | 1.06-1.11 | 1.74E-16 | 7.23E-02 | 0.4519 | 0.4988 | 0.808 |
| 11p12 | PRR5L | 11:36401868 | rs77229041 | G | T | 1.18 | 1.13-1.24 | 4.56E-14 | 2.22E-01 | 0.058 | 0.0501 | 2.033 |
| 11q13.1 | GPR137 | 11:64284975 | rs2510066 | C | T | 0.94 | 0.92-0.96 | 4.77E-10 | 3.08E-01 | 0.4178 | 0.3933 | 1.106 |
| 11q22.3 | ZC3H12C | 11:110093651 | rs7119747 | A | G | 0.94 | 0.92-0.96 | 4.88E-09 | 1.19E-01 | 0.6065 | 0.6888 | 0.847 |
| 11q24.3 | ETS1 | 11:128526843 | rs7117118 | A | G | 1.08 | 1.06-1.11 | 7.77E-12 | 4.67E-01 | 0.2333 | 0.2352 | 1.027 |
| 12q13.3 | STAT2 | 12:56344189 | rs2066808 | A | G | 0.89 | 0.86-0.93 | 2.07E-09 | 4.19E-02 | 0.0545 | 0.0902 | 0.826 |
| 12q24.12 | SH2B3 | 12:111446804 | rs3184504 | T | C | 0.93 | 0.92-0.95 | 8.87E-12 | 1.16E-01 | 0.5911 | 0.5557 | 1.179 |
| **12q24.13** | **RPL6** | **12:112402962** | **rs11066283** | **A** | **G** | **1.06** | **1.04-1.08** | **1.97E-09** | **3.79E-01** | **0.379** | **0.4053** | **0.88** |
| 14q13.2 | NFKBIA | 14:35402011 | rs8904 | G | A | 0.94 | 0.92-0.96 | 1.12E-10 | 1.20E-04 | 0.388 | 0.4104 | 1.064 |
| 16p13.13 | RMI2 | 16:11251046 | rs2021511 | C | T | 0.92 | 0.9-0.94 | 7.60E-15 | 6.02E-02 | 0.266 | 0.3266 | 0.973 |
| **16p12.1** | **IL4R** | **16:27344903** | **rs144651842** | **G** | **A** | **1.21** | **1.15-1.27** | **7.21E-14** | **3.34E-01** | **0.078** | **0.0254** | **119.05** |
| 17q11.2 | NOS2 | 17:27797882 | rs28998802 | G | A | 1.12 | 1.09-1.15 | 6.24E-16 | 8.83E-03 | 0.1849 | 0.1404 | 1.359 |
| 17q21.2 | SMARCE1 | 17:40608272 | rs112401631 | T | A | 1.34 | 1.2-1.48 | 4.79E-08 | 7.81E-02 | 0.0061 | 0.0081 | 0.293 |

| locus | nearest gene | chr:pos | rsid | ref | alt | OR | CI 95 % | p | het_p | af_alt FinnGen | af_alt EstBB | Fin enr. |
|---|---|---|---|---|---|---|---|---|---|---|---|---|
| 18p11.21 | PTPN2 | 18:12887751 | rs8096327 | A | G | 1.06 | 1.04-1.08 | 3.89E-08 | 4.11E-01 | 0.3478 | 0.3459 | 0.849 |
| 18q21.2 | POLI | 18:54306969 | rs2851877 | A | T | 1.07 | 1.05-1.1 | 2.72E-10 | 7.50E-02 | 0.2404 | 0.2964 | 0.8 |
| 19p13.2 | TYK2 | 19:10352442 | rs34536443 | G | C | 0.8 | 0.75-0.84 | 4.90E-15 | 7.96E-03 | 0.0306 | 0.0334 | 0.608 |
| **19p13.11** | **LRRC25** | **19:18402122** | **rs1560117** | **C** | **T** | **1.06** | **1.04-1.08** | **6.05E-09** | **3.42E-01** | **0.5344** | **0.4935** | **1.124** |
| 20q13.13 | RNF114 | 20:49927636 | rs6125816 | A | G | 0.95 | 0.93-0.96 | 2.74E-08 | 2.49E-02 | 0.5799 | 0.5314 | 1.416 |
| 22q11.21 | UBE2L3 | 22:21563190 | rs5998509 | C | T | 1.08 | 1.06-1.11 | 1.29E-10 | 5.81E-02 | 0.293 | 0.1712 | 1.578 |

**Table S3. Genetic correlation results (FDR-corrected p-value > 0.05) from LDSC pipeline.** Results showed in order of p-values from smallest to largest. Columns: Trait, trait name as it appears in the LDSC pipeline; Group, a category for each trait as it appears in the LDSC pipeline; rg, genetic correlation estimate (varies between -1 and 1); se, standard error of rg; p, p-value of correlation; and p_fdr, FDR-corrected p-value.

| Trait | Group | rg | se | p | p_fdr |
|---|---|---|---|---|---|
| Overall health rating | activity_fitness_sleep | 0.2966 | 0.0350 | 2.51E-17 | 1.92E-14 |
| Years of schooling | education_intelligence | -0.2534 | 0.0326 | 8.32E-15 | 3.19E-12 |
| Long-standing illness, disability or infirmity | diseases | 0.3222 | 0.0418 | 1.31E-14 | 3.35E-12 |
| Types of physical activity in last 4 weeks: None of the above | activity_fitness_sleep | 0.3593 | 0.0489 | 2.06E-13 | 3.94E-11 |
| Current tobacco smoking | smoking | 0.2904 | 0.0399 | 3.43E-13 | 5.26E-11 |
| Qualifications: A levels/AS levels or equivalent | education_intelligence | -0.2561 | 0.0356 | 6.58E-13 | 8.40E-11 |
| Leg fat mass (right) | anthropometry | 0.2093 | 0.0297 | 1.70E-12 | 1.86E-10 |
| Qualifications: College or University degree | education_intelligence | -0.2481 | 0.0355 | 2.57E-12 | 2.26E-10 |
| Leg fat mass (left) | anthropometry | 0.2068 | 0.0296 | 2.65E-12 | 2.26E-10 |
| Leg fat percentage (right) | anthropometry | 0.2215 | 0.0317 | 3.00E-12 | 2.30E-10 |
| Leg fat percentage (left) | anthropometry | 0.2175 | 0.0315 | 4.79E-12 | 3.34E-10 |
| Whole body fat mass | anthropometry | 0.1979 | 0.0289 | 6.89E-12 | 4.40E-10 |
| Body fat percentage | anthropometry | 0.2010 | 0.0297 | 1.39E-11 | 8.21E-10 |
| Arm fat mass (right) | anthropometry | 0.1951 | 0.0291 | 2.14E-11 | 1.17E-09 |
| Taking other prescription medications | drugs_supplements | 0.3093 | 0.0466 | 3.11E-11 | 1.59E-09 |
| Arm fat mass (left) | anthropometry | 0.1919 | 0.0293 | 5.79E-11 | 2.77E-09 |
| Trunk fat mass | anthropometry | 0.1878 | 0.0288 | 6.64E-11 | 2.99E-09 |
| Arm fat percentage (right) | anthropometry | 0.1942 | 0.0300 | 9.92E-11 | 4.15E-09 |
| Trunk fat percentage | anthropometry | 0.1898 | 0.0294 | 1.03E-10 | 4.15E-09 |
| Body mass index (BMI) | anthropometry | 0.1947 | 0.0303 | 1.34E-10 | 5.14E-09 |
| smoking initiation | smoking | 0.2276 | 0.0356 | 1.58E-10 | 5.77E-09 |
| Waist circumference | anthropometry | 0.1885 | 0.0295 | 1.68E-10 | 5.84E-09 |
| Qualifications: None of the above | education_intelligence | 0.2319 | 0.0365 | 2.01E-10 | 6.69E-09 |
| Usual walking pace | activity_fitness_sleep | -0.2196 | 0.0349 | 3.02E-10 | 9.65E-09 |
| Smoking status: Never | smoking | -0.2454 | 0.0391 | 3.37E-10 | 1.03E-08 |
| Arm fat percentage (left) | anthropometry | 0.1869 | 0.0298 | 3.54E-10 | 1.04E-08 |
| Smoking status: Current | smoking | 0.2677 | 0.0428 | 3.84E-10 | 1.09E-08 |
| Type 2 diabetes | diseases | 0.2721 | 0.0442 | 7.23E-10 | 1.98E-08 |
| Number of treatments/medications taken | drugs_supplements | 0.2607 | 0.0429 | 1.17E-09 | 3.09E-08 |
| Health satisfaction | activity_fitness_sleep | 0.3065 | 0.0511 | 2.05E-09 | 5.24E-08 |
| Wheeze or whistling in the chest in last year | lung_function | 0.2663 | 0.0448 | 2.76E-09 | 6.82E-08 |
| Age at first live birth | gynecology_fertility | -0.2441 | 0.0422 | 7.44E-09 | 1.78E-07 |
| Types of physical activity in last 4 weeks: Walking for pleasure (not as a means of transport) | activity_fitness_sleep | -0.2502 | 0.0436 | 9.35E-09 | 2.17E-07 |

| Trait | Group | rg | se | p | p_fdr |
|---|---|---|---|---|---|
| Inflammatory bowel disease | diseases | 0.2717 | 0.0477 | 1.19E-08 | 2.67E-07 |
| Diagnoses - main ICD10: R07 Pain in throat and chest | diseases | 0.3815 | 0.0674 | 1.48E-08 | 3.25E-07 |
| Age completed full time education | education_intelligence | -0.2437 | 0.0431 | 1.62E-08 | 3.44E-07 |
| Weight | anthropometry | 0.1606 | 0.0286 | 2.04E-08 | 4.22E-07 |
| Maternal smoking around birth | smoking | 0.2532 | 0.0462 | 4.16E-08 | 8.39E-07 |
| Illness, injury, bereavement, stress in last 2 years: Financial difficulties | work_stress | 0.2567 | 0.0483 | 1.08E-07 | 2.13E-06 |
| Qualifications: Other professional qualifications eg: nursing, teaching | education_intelligence | -0.2405 | 0.0460 | 1.65E-07 | 3.16E-06 |
| Miserableness | mood_behaviour | 0.1913 | 0.0368 | 2.08E-07 | 3.88E-06 |
| Age Of Smoking Initiation | smoking | -0.2471 | 0.0477 | 2.25E-07 | 4.11E-06 |
| Frequency of tiredness / lethargy in last 2 weeks | mood_behaviour | 0.1994 | 0.0388 | 2.86E-07 | 5.10E-06 |
| Past tobacco smoking | smoking | -0.1870 | 0.0366 | 3.26E-07 | 5.67E-06 |
| Hip circumference | anthropometry | 0.1511 | 0.0296 | 3.41E-07 | 5.81E-06 |
| Ratio of linoleic acid to total fatty acids | NMR_metabo_UKBB | -0.2650 | 0.0521 | 3.71E-07 | 6.18E-06 |
| Time spent watching television (TV) | activity_fitness_sleep | 0.1956 | 0.0385 | 3.89E-07 | 6.33E-06 |
| Ratio of polyunsaturated fatty acids to total fatty acids | NMR_metabo_UKBB | -0.2235 | 0.0446 | 5.30E-07 | 8.45E-06 |
| Diabetes diagnosed by doctor | diseases | 0.2304 | 0.0463 | 6.47E-07 | 1.01E-05 |
| College completion | education_intelligence | -0.2673 | 0.0544 | 8.73E-07 | 1.34E-05 |
| Financial situation satisfaction | work_stress | 0.2922 | 0.0595 | 8.96E-07 | 1.35E-05 |
| Ever smoked | smoking | 0.1702 | 0.0347 | 9.27E-07 | 1.37E-05 |
| HDL cholesterol | biomarker | -0.1585 | 0.0325 | 1.09E-06 | 1.57E-05 |
| Mood swings | mood_behaviour | 0.1859 | 0.0382 | 1.11E-06 | 1.57E-05 |
| Pain type(s) experienced in last month: Knee pain | pain_skeletal | 0.2217 | 0.0457 | 1.20E-06 | 1.67E-05 |
| Medication for pain relief, constipation, heartburn: None of the above | drugs_supplements | -0.1985 | 0.0415 | 1.70E-06 | 2.32E-05 |
| Types of physical activity in last 4 weeks: Other exercises (eg: swimming, cycling, keep fit, bowling) | activity_fitness_sleep | -0.1907 | 0.0402 | 2.14E-06 | 2.87E-05 |
| Chest pain or discomfort | pain_skeletal | 0.2049 | 0.0434 | 2.36E-06 | 3.11E-05 |
| Father's age at death | family_memb_health | -0.2725 | 0.0584 | 3.04E-06 | 3.95E-05 |
| Age at last live birth | gynecology_fertility | -0.2188 | 0.0471 | 3.39E-06 | 4.33E-05 |
| Vascular/heart problems diagnosed by doctor: None of the above | diseases | -0.1687 | 0.0364 | 3.51E-06 | 4.41E-05 |
| Mouth/teeth dental problems: Dentures | dental | 0.1899 | 0.0411 | 3.79E-06 | 4.68E-05 |
| Other serious medical condition/disability diagnosed by doctor | diseases | 0.2542 | 0.0554 | 4.48E-06 | 5.45E-05 |
| Pack years of smoking | smoking | 0.2208 | 0.0486 | 5.61E-06 | 6.71E-05 |
| Back pain for 3+ months | pain_skeletal | 0.3194 | 0.0709 | 6.65E-06 | 7.84E-05 |
| Illness, injury, bereavement, stress in last 2 years: Serious illness, injury or assault to yourself | work_stress | 0.2886 | 0.0643 | 7.05E-06 | 8.18E-05 |

| Trait | Group | rg | se | p | p_fdr |
|---|---|---|---|---|---|
| Vascular/heart problems diagnosed by doctor: High blood pressure | diseases | 0.1632 | 0.0365 | 7.92E-06 | 9.05E-05 |
| Frequency of unenthusiasm / disinterest in last 2 weeks | mood_behaviour | 0.2027 | 0.0456 | 8.70E-06 | 9.80E-05 |
| Types of physical activity in last 4 weeks: Light DIY (eg: pruning, watering the lawn) | activity_fitness_sleep | -0.2142 | 0.0483 | 9.15E-06 | 1.02E-04 |
| Forced expiratory volume in 1-second (FEV1), predicted percentage | lung_function | -0.1824 | 0.0412 | 9.60E-06 | 1.05E-04 |
| Ratio of polyunsaturated fatty acids to monounsaturated fatty acids | NMR_metabo_UKBB | -0.1931 | 0.0438 | 1.06E-05 | 1.14E-04 |
| Frequency of tenseness / restlessness in last 2 weeks | mood_behaviour | 0.1806 | 0.0412 | 1.16E-05 | 1.22E-04 |
| Light smokers, at least 100 smokes in lifetime | smoking | 0.2587 | 0.0590 | 1.16E-05 | 1.22E-04 |
| Major depressive disorder (ICD-10 coded) | diseases | 0.3205 | 0.0735 | 1.30E-05 | 1.35E-04 |
| Seen doctor (GP) for nerves, anxiety, tension or depression | mood_behaviour | 0.1586 | 0.0364 | 1.33E-05 | 1.36E-04 |
| Diagnoses - main ICD10: R10 Abdominal and pelvic pain | diseases | 0.3527 | 0.0813 | 1.43E-05 | 1.44E-04 |
| Number of operations, self-reported | diseases | 0.1938 | 0.0450 | 1.62E-05 | 1.61E-04 |
| Qualifications: O levels/GCSEs or equivalent | education_intelligence | -0.1797 | 0.0420 | 1.87E-05 | 1.80E-04 |
| Phospholipids to total lipids ratio in large HDL | NMR_metabo_UKBB | 0.2041 | 0.0477 | 1.87E-05 | 1.80E-04 |
| Pain type(s) experienced in last month: None of the above | pain_skeletal | -0.1798 | 0.0420 | 1.88E-05 | 1.80E-04 |
| Ratio of omega-6 fatty acids to total fatty acids | NMR_metabo_UKBB | -0.2051 | 0.0481 | 2.01E-05 | 1.90E-04 |
| Alcohol usually taken with meals | alcohol | -0.1875 | 0.0440 | 2.07E-05 | 1.93E-04 |
| Blood clot, DVT, bronchitis, emphysema, asthma, rhinitis, eczema, allergy diagnosed by doctor: Emphysema/chronic bronchitis | diseases | 0.3215 | 0.0761 | 2.38E-05 | 2.20E-04 |
| Job involves heavy manual or physical work | activity_fitness_sleep | 0.1776 | 0.0424 | 2.78E-05 | 2.53E-04 |
| Pain type(s) experienced in last month: Pain all over the body | pain_skeletal | 0.3467 | 0.0829 | 2.92E-05 | 2.63E-04 |
| Sleeplessness / insomnia | work_stress | 0.1725 | 0.0414 | 3.09E-05 | 2.75E-04 |
| Exposure to tobacco smoke outside home | smoking | 0.2058 | 0.0496 | 3.34E-05 | 2.94E-04 |
| Vascular/heart problems diagnosed by doctor: Angina | diseases | 0.2366 | 0.0575 | 3.82E-05 | 3.29E-04 |
| Waist-to-hip ratio | anthropometry | 0.1923 | 0.0467 | 3.82E-05 | 3.29E-04 |
| triglycerides | biomarker | 0.1486 | 0.0363 | 4.18E-05 | 3.56E-04 |
| Seen a psychiatrist for nerves, anxiety, tension or depression | mood_behaviour | 0.1890 | 0.0463 | 4.37E-05 | 3.68E-04 |
| Pain type(s) experienced in last month: Stomach or abdominal pain | pain_skeletal | 0.2376 | 0.0582 | 4.42E-05 | 3.68E-04 |
| Shortness of breath walking on level ground | activity_fitness_sleep | 0.2680 | 0.0658 | 4.67E-05 | 3.84E-04 |
| Mouth/teeth dental problems: None of the above | dental | -0.1724 | 0.0424 | 4.78E-05 | 3.90E-04 |
| Falls in the last year | activity_fitness_sleep | 0.1966 | 0.0485 | 4.96E-05 | 4.00E-04 |
| Cholesteryl esters to total lipids ratio in large HDL | NMR_metabo_UKBB | -0.1879 | 0.0465 | 5.30E-05 | 4.22E-04 |
| Intelligence | education_intelligence | -0.1499 | 0.0372 | 5.51E-05 | 4.35E-04 |
| Loneliness, isolation | mood_behaviour | 0.1664 | 0.0413 | 5.67E-05 | 4.39E-04 |

| Trait | Group | rg | se | p | p_fdr |
|---|---|---|---|---|---|
| Medication for cholesterol, blood pressure, diabetes, or take exogenous hormones: Cholesterol lowering medication | drugs_supplements | 0.2165 | 0.0538 | 5.67E-05 | 4.39E-04 |
| Smoking status: Previous | smoking | 0.1824 | 0.0454 | 5.84E-05 | 4.48E-04 |
| Ratio of monounsaturated fatty acids to total fatty acids | NMR_metabo_UKBB | 0.1727 | 0.0433 | 6.70E-05 | 5.08E-04 |
| apolipoprotein A-I | biomarker | -0.1357 | 0.0342 | 7.14E-05 | 5.36E-04 |
| Types of transport used (excluding work): Walk | activity_fitness_sleep | -0.1745 | 0.0441 | 7.71E-05 | 5.73E-04 |
| Fed-up feelings | mood_behaviour | 0.1464 | 0.0373 | 8.72E-05 | 6.42E-04 |
| Ever stopped smoking for 6+ months | smoking | 0.4456 | 0.1140 | 9.32E-05 | 6.80E-04 |
| Arm fat-free mass (left) | anthropometry | 0.1149 | 0.0295 | 9.52E-05 | 6.88E-04 |
| Frequency of depressed mood in last 2 weeks | mood_behaviour | 0.1681 | 0.0432 | 1.01E-04 | 7.20E-04 |
| Medication for pain relief, constipation, heartburn: Laxatives (e.g. Dulcolax, Senokot) | drugs_supplements | 0.2721 | 0.0701 | 1.02E-04 | 7.26E-04 |
| Coronary artery disease | diseases | 0.1555 | 0.0402 | 1.10E-04 | 7.72E-04 |
| Cholesterol to total lipids ratio in medium LDL | NMR_metabo_UKBB | -0.2713 | 0.0702 | 1.11E-04 | 7.72E-04 |
| Mouth/teeth dental problems: Loose teeth | dental | 0.2685 | 0.0695 | 1.12E-04 | 7.72E-04 |
| Leg pain on walking | pain_skeletal | 0.2428 | 0.0630 | 1.16E-04 | 7.91E-04 |
| Triglycerides to total lipids ratio in large LDL | NMR_metabo_UKBB | 0.2478 | 0.0643 | 1.17E-04 | 7.91E-04 |
| Fractured bone site(s): Ankle | pain_skeletal | 0.3889 | 0.1010 | 1.18E-04 | 7.94E-04 |
| Phospholipids to total lipids ratio in medium HDL | NMR_metabo_UKBB | 0.1765 | 0.0459 | 1.22E-04 | 8.12E-04 |
| Medication for cholesterol, blood pressure, diabetes, or take exogenous hormones: None of the above | drugs_supplements | -0.1780 | 0.0466 | 1.32E-04 | 8.74E-04 |
| Smoking/smokers in household | smoking | 0.2708 | 0.0713 | 1.45E-04 | 9.51E-04 |
| Forced expiratory volume in 1-second (FEV1) | lung_function | -0.1363 | 0.0360 | 1.52E-04 | 9.78E-04 |
| Medication for pain relief, constipation, heartburn: Omeprazole (e.g. Zanprol) | drugs_supplements | 0.2075 | 0.0548 | 1.52E-04 | 9.78E-04 |
| Triglycerides to total lipids ratio in IDL | NMR_metabo_UKBB | 0.2016 | 0.0535 | 1.64E-04 | 1.04E-03 |
| Pulse wave peak to peak time | cardiovascular | -0.2545 | 0.0675 | 1.64E-04 | 1.04E-03 |
| Forced vital capacity (FVC) | lung_function | -0.1328 | 0.0353 | 1.69E-04 | 1.04E-03 |
| Glycoprotein acetyls | NMR_metabo_UKBB | 0.1771 | 0.0471 | 1.70E-04 | 1.04E-03 |
| Cigarettes per Day | smoking | 0.1693 | 0.0451 | 1.71E-04 | 1.04E-03 |
| Cholesterol to total lipids ratio in large HDL | NMR_metabo_UKBB | -0.1767 | 0.0470 | 1.71E-04 | 1.04E-03 |
| Pain type(s) experienced in last month: Back pain | pain_skeletal | 0.1686 | 0.0449 | 1.72E-04 | 1.04E-03 |
| Cigarettes smoked per day | smoking | 0.1692 | 0.0450 | 1.72E-04 | 1.04E-03 |
| Illness, injury, bereavement, stress in last 2 years: None of the above | work_stress | -0.2051 | 0.0549 | 1.88E-04 | 1.12E-03 |
| Noisy workplace | work_stress | 0.2217 | 0.0594 | 1.89E-04 | 1.12E-03 |
| Qualifications: CSEs or equivalent | education_intelligence | 0.1969 | 0.0529 | 1.97E-04 | 1.16E-03 |
| Cholesterol to total lipids ratio in large LDL | NMR_metabo_UKBB | -0.2547 | 0.0685 | 1.99E-04 | 1.16E-03 |
| Blood clot, DVT, bronchitis, emphysema, asthma, rhinitis, eczema, allergy diagnosed by doctor: Blood clot in the leg (DVT) | diseases | 0.2753 | 0.0745 | 2.18E-04 | 1.27E-03 |

| Trait | Group | rg | se | p | p_fdr |
|---|---|---|---|---|---|
| Basal metabolic rate | anthropometry | 0.1076 | 0.0292 | 2.28E-04 | 1.31E-03 |
| Eye problems/disorders: Diabetes related eye disease | diseases | 0.4061 | 0.1104 | 2.34E-04 | 1.34E-03 |
| Chest pain or discomfort walking normally | pain_skeletal | 0.2982 | 0.0812 | 2.38E-04 | 1.35E-03 |
| Ever had bowel cancer screening | diseases | 0.2211 | 0.0606 | 2.64E-04 | 1.48E-03 |
| Types of physical activity in last 4 weeks: Strenuous sports | activity_fitness_sleep | -0.1809 | 0.0496 | 2.65E-04 | 1.48E-03 |
| Ratio of saturated fatty acids to total fatty acids | NMR_metabo_UKBB | 0.2014 | 0.0559 | 3.14E-04 | 1.73E-03 |
| Forced expiratory volume in 1-second (FEV1), Best measure | lung_function | -0.1300 | 0.0361 | 3.14E-04 | 1.73E-03 |
| Leg fat-free mass (left) | anthropometry | 0.1061 | 0.0295 | 3.17E-04 | 1.73E-03 |
| Triglycerides to total lipids ratio in medium LDL | NMR_metabo_UKBB | 0.2303 | 0.0642 | 3.32E-04 | 1.80E-03 |
| Pulse wave Arterial Stiffness index | cardiovascular | 0.2462 | 0.0687 | 3.41E-04 | 1.84E-03 |
| Alcohol intake frequency. | alcohol | 0.1258 | 0.0353 | 3.62E-04 | 1.94E-03 |
| Townsend deprivation index at recruitment | work_stress | 0.1764 | 0.0495 | 3.64E-04 | 1.94E-03 |
| Medication for pain relief, constipation, heartburn: Aspirin | drugs_supplements | 0.2023 | 0.0570 | 3.83E-04 | 2.02E-03 |
| Pain type(s) experienced in last month: Neck or shoulder pain | pain_skeletal | 0.1685 | 0.0475 | 3.90E-04 | 2.05E-03 |
| Job involves mainly walking or standing | activity_fitness_sleep | 0.1505 | 0.0427 | 4.17E-04 | 2.17E-03 |
| Loud music exposure frequency | eye_ear | 0.2582 | 0.0736 | 4.53E-04 | 2.34E-03 |
| Cholesteryl esters to total lipids ratio in very small VLDL | NMR_metabo_UKBB | -0.1769 | 0.0506 | 4.72E-04 | 2.43E-03 |
| Transport type for commuting to job workplace: Cycle | activity_fitness_sleep | -0.1821 | 0.0531 | 6.05E-04 | 3.09E-03 |
| Diagnoses - main ICD10: M25 Other joint disorders, not elsewhere classified | diseases | 0.4868 | 0.1421 | 6.10E-04 | 3.09E-03 |
| Average weekly red wine intake | alcohol | -0.1537 | 0.0449 | 6.14E-04 | 3.09E-03 |
| Degree of unsaturation | NMR_metabo_UKBB | -0.1721 | 0.0503 | 6.21E-04 | 3.11E-03 |
| Cholesterol to total lipids ratio in very small VLDL | NMR_metabo_UKBB | -0.1739 | 0.0510 | 6.50E-04 | 3.23E-03 |
| Cholesterol to total lipids ratio in medium HDL | NMR_metabo_UKBB | -0.1548 | 0.0455 | 6.61E-04 | 3.26E-03 |
| Diagnoses - main ICD10: I25 Chronic ischaemic heart disease | diseases | 0.2111 | 0.0620 | 6.64E-04 | 3.26E-03 |
| Had other major operations | diseases | 0.1980 | 0.0583 | 6.86E-04 | 3.35E-03 |
| Forced vital capacity (FVC), Best measure | lung_function | -0.1197 | 0.0354 | 7.27E-04 | 3.53E-03 |
| Neuroticism score | mood_behaviour | 0.1146 | 0.0340 | 7.56E-04 | 3.63E-03 |
| Cholesteryl esters to total lipids ratio in medium HDL | NMR_metabo_UKBB | -0.1535 | 0.0456 | 7.58E-04 | 3.63E-03 |
| Free cholesterol to total lipids ratio in large LDL | NMR_metabo_UKBB | -0.1518 | 0.0455 | 8.41E-04 | 4.00E-03 |
| Cholesterol to total lipids ratio in IDL | NMR_metabo_UKBB | -0.1820 | 0.0546 | 8.52E-04 | 4.03E-03 |
| Average weekly fortified wine intake | alcohol | -0.2652 | 0.0797 | 8.75E-04 | 4.11E-03 |
| Diagnoses - secondary ICD10: M06.99 Rheumatoid arthritis, unspecified (Site unspecified) | diseases | 0.4613 | 0.1388 | 8.87E-04 | 4.14E-03 |
| Cholesteryl esters in very large HDL | NMR_metabo_UKBB | -0.1533 | 0.0462 | 9.00E-04 | 4.18E-03 |
| Sensitivity / hurt feelings | mood_behaviour | 0.1263 | 0.0384 | 9.94E-04 | 4.59E-03 |
| Parental longevity (combined parental age at death) | family_memb_health | -0.2087 | 0.0635 | 1.00E-03 | 4.60E-03 |
| Transport type for commuting to job workplace: Car/motor vehicle | activity_fitness_sleep | 0.2245 | 0.0685 | 1.04E-03 | 4.74E-03 |
| Triglycerides to total lipids ratio in very small VLDL | NMR_metabo_UKBB | 0.1668 | 0.0509 | 1.05E-03 | 4.75E-03 |

| Trait | Group | rg | se | p | p_fdr |
|---|---|---|---|---|---|
| Phospholipids to total lipids ratio in very small VLDL | NMR_metabo_UKBB | 0.1772 | 0.0541 | 1.06E-03 | 4.78E-03 |
| Types of transport used (excluding work): Cycle | activity_fitness_sleep | -0.1648 | 0.0505 | 1.09E-03 | 4.86E-03 |
| Free cholesterol to total lipids ratio in medium LDL | NMR_metabo_UKBB | -0.1507 | 0.0461 | 1.09E-03 | 4.86E-03 |
| Triglycerides in large LDL | NMR_metabo_UKBB | 0.1487 | 0.0457 | 1.13E-03 | 5.01E-03 |
| Arm fat-free mass (right) | anthropometry | 0.0964 | 0.0297 | 1.15E-03 | 5.07E-03 |
| HOMA-IR | biomarker | 0.3026 | 0.0935 | 1.22E-03 | 5.33E-03 |
| Cholesterol in very large HDL | NMR_metabo_UKBB | -0.1524 | 0.0472 | 1.25E-03 | 5.45E-03 |
| Free cholesterol to total lipids ratio in small LDL | NMR_metabo_UKBB | -0.1600 | 0.0497 | 1.29E-03 | 5.56E-03 |
| Average weekly spirits intake | alcohol | 0.1727 | 0.0537 | 1.31E-03 | 5.62E-03 |
| Triglycerides to total lipids ratio in small LDL | NMR_metabo_UKBB | 0.1604 | 0.0506 | 1.51E-03 | 6.46E-03 |
| Depressive symptoms | mood_behaviour | 0.1708 | 0.0540 | 1.54E-03 | 6.55E-03 |
| Triglycerides in small HDL | NMR_metabo_UKBB | 0.1327 | 0.0419 | 1.55E-03 | 6.55E-03 |
| Alcohol intake versus 10 years previously | alcohol | 0.1524 | 0.0482 | 1.56E-03 | 6.55E-03 |
| Leg fat-free mass (right) | anthropometry | 0.0927 | 0.0294 | 1.59E-03 | 6.67E-03 |
| Concentration of very large HDL particles | NMR_metabo_UKBB | -0.1513 | 0.0481 | 1.66E-03 | 6.92E-03 |
| Pain type(s) experienced in last month: Hip pain | pain_skeletal | 0.1545 | 0.0494 | 1.77E-03 | 7.32E-03 |
| Mother's age at death | family_memb_health | -0.2066 | 0.0661 | 1.78E-03 | 7.34E-03 |
| Impedance of arm (left) | anthropometry | -0.0920 | 0.0295 | 1.81E-03 | 7.40E-03 |
| Triglycerides in LDL | NMR_metabo_UKBB | 0.1388 | 0.0447 | 1.90E-03 | 7.75E-03 |
| Maximum heart rate during fitness test | cardiovascular | -0.1867 | 0.0603 | 1.96E-03 | 7.95E-03 |
| Diagnoses - main ICD10: K52 Other non-infective gastro-enteritis and colitis | diseases | 0.4020 | 0.1301 | 2.01E-03 | 8.09E-03 |
| Triglycerides in IDL | NMR_metabo_UKBB | 0.1379 | 0.0447 | 2.03E-03 | 8.14E-03 |
| Histidine | NMR_metabo_UKBB | -0.2370 | 0.0771 | 2.10E-03 | 8.37E-03 |
| Total lipids in very large HDL | NMR_metabo_UKBB | -0.1420 | 0.0463 | 2.17E-03 | 8.62E-03 |
| Cholesteryl esters in large HDL | NMR_metabo_UKBB | -0.1338 | 0.0437 | 2.18E-03 | 8.62E-03 |
| Triglycerides to total lipids ratio in small HDL | NMR_metabo_UKBB | 0.1324 | 0.0433 | 2.23E-03 | 8.77E-03 |
| Transport type for commuting to job workplace: Walk | activity_fitness_sleep | -0.2415 | 0.0793 | 2.31E-03 | 9.02E-03 |
| Reason for reducing amount of alcohol drunk: Illness or ill health | alcohol | 0.3651 | 0.1201 | 2.37E-03 | 9.23E-03 |
| Duration walking for pleasure | activity_fitness_sleep | -0.1673 | 0.0551 | 2.40E-03 | 9.27E-03 |
| Medication for cholesterol, blood pressure, diabetes, or take exogenous hormones: Blood pressure medication | drugs_supplements | 0.1269 | 0.0419 | 2.49E-03 | 9.58E-03 |
| Medication for pain relief, constipation, heartburn: Paracetamol | drugs_supplements | 0.1370 | 0.0455 | 2.61E-03 | 1.00E-02 |
| Cholesterol in large HDL | NMR_metabo_UKBB | -0.1319 | 0.0438 | 2.63E-03 | 1.00E-02 |
| Cholesteryl esters to total lipids ratio in medium VLDL | NMR_metabo_UKBB | -0.1422 | 0.0475 | 2.74E-03 | 1.04E-02 |
| Triglycerides in medium LDL | NMR_metabo_UKBB | 0.1308 | 0.0438 | 2.84E-03 | 1.07E-02 |
| Diagnoses - main ICD10: I20 Angina pectoris | diseases | 0.2614 | 0.0877 | 2.87E-03 | 1.08E-02 |
| Phospholipids to total lipids ratio in large VLDL | NMR_metabo_UKBB | 0.1389 | 0.0466 | 2.90E-03 | 1.08E-02 |
| Phospholipids in very large HDL | NMR_metabo_UKBB | -0.1348 | 0.0457 | 3.18E-03 | 1.18E-02 |

| Trait | Group | rg | se | p | p_fdr |
|---|---|---|---|---|---|
| Ever used hormone-replacement therapy (HRT) | gynecology_fertility | 0.1681 | 0.0570 | 3.20E-03 | 1.18E-02 |
| Cholesteryl esters to total lipids ratio in very large VLDL | NMR_metabo_UKBB | -0.1413 | 0.0480 | 3.26E-03 | 1.20E-02 |
| Free cholesterol to total lipids ratio in small VLDL | NMR_metabo_UKBB | -0.1513 | 0.0515 | 3.31E-03 | 1.21E-02 |
| Triglycerides to total lipids ratio in very large HDL | NMR_metabo_UKBB | 0.1284 | 0.0440 | 3.51E-03 | 1.28E-02 |
| Former vs current smoker | smoking | -0.2757 | 0.0945 | 3.52E-03 | 1.28E-02 |
| Whole body fat-free mass | anthropometry | 0.0861 | 0.0295 | 3.56E-03 | 1.29E-02 |
| Ratio of triglycerides to phosphoglycerides | NMR_metabo_UKBB | 0.1216 | 0.0419 | 3.72E-03 | 1.33E-02 |
| Cholesteryl esters to total lipids ratio in IDL | NMR_metabo_UKBB | -0.1741 | 0.0600 | 3.73E-03 | 1.33E-02 |
| Concentration of large HDL particles | NMR_metabo_UKBB | -0.1302 | 0.0449 | 3.73E-03 | 1.33E-02 |
| Cholesterol to total lipids ratio in small HDL | NMR_metabo_UKBB | -0.1640 | 0.0566 | 3.78E-03 | 1.33E-02 |
| Free cholesterol in very large HDL | NMR_metabo_UKBB | -0.1423 | 0.0491 | 3.79E-03 | 1.33E-02 |
| Triglycerides in very small VLDL | NMR_metabo_UKBB | 0.1255 | 0.0433 | 3.79E-03 | 1.33E-02 |
| Bilateral oophorectomy (both ovaries removed) | gynecology_fertility | 0.2235 | 0.0773 | 3.85E-03 | 1.34E-02 |
| Neck/shoulder pain for 3+ months | pain_skeletal | 0.2603 | 0.0901 | 3.86E-03 | 1.34E-02 |
| Diagnoses - main ICD10: K21 Gastro-oesophageal reflux disease | diseases | 0.2866 | 0.0993 | 3.88E-03 | 1.35E-02 |
| Free cholesterol to total lipids ratio in IDL | NMR_metabo_UKBB | -0.1502 | 0.0521 | 3.96E-03 | 1.36E-02 |
| Cholesterol to total lipids ratio in medium VLDL | NMR_metabo_UKBB | -0.1380 | 0.0479 | 3.97E-03 | 1.36E-02 |
| Triglycerides to total lipids ratio in medium HDL | NMR_metabo_UKBB | 0.1290 | 0.0448 | 3.98E-03 | 1.36E-02 |
| Cholesteryl esters to total lipids ratio in large VLDL | NMR_metabo_UKBB | -0.1405 | 0.0489 | 4.02E-03 | 1.37E-02 |
| Whole body water mass | anthropometry | 0.0847 | 0.0295 | 4.12E-03 | 1.40E-02 |
| Free cholesterol to total lipids ratio in medium HDL | NMR_metabo_UKBB | -0.1412 | 0.0494 | 4.25E-03 | 1.43E-02 |
| Free cholesterol to total lipids ratio in very large HDL | NMR_metabo_UKBB | 0.1353 | 0.0475 | 4.34E-03 | 1.46E-02 |
| Phospholipids to total lipids ratio in small VLDL | NMR_metabo_UKBB | -0.1512 | 0.0531 | 4.38E-03 | 1.46E-02 |
| Childhood obesity | anthropometry | 0.1856 | 0.0652 | 4.41E-03 | 1.46E-02 |
| Alcohol drinker status: Previous | alcohol | 0.2095 | 0.0736 | 4.41E-03 | 1.46E-02 |
| Neuroticism | mood_behaviour | 0.1037 | 0.0365 | 4.48E-03 | 1.48E-02 |
| Blood clot, DVT, bronchitis, emphysema, asthma, rhinitis, eczema, allergy diagnosed by doctor: Blood clot in the lung | diseases | 0.3174 | 0.1122 | 4.67E-03 | 1.54E-02 |
| Monounsaturated fatty acids | NMR_metabo_UKBB | 0.1238 | 0.0438 | 4.71E-03 | 1.54E-02 |
| Comparative body size at age 10 | anthropometry | 0.0992 | 0.0351 | 4.73E-03 | 1.54E-02 |
| Cholesterol to total lipids ratio in very large VLDL | NMR_metabo_UKBB | -0.1393 | 0.0495 | 4.87E-03 | 1.58E-02 |
| Fluid intelligence score | education_intelligence | -0.1153 | 0.0411 | 5.04E-03 | 1.63E-02 |
| Transport type for commuting to job workplace: Public transport | activity_fitness_sleep | -0.1775 | 0.0633 | 5.06E-03 | 1.63E-02 |
| Cholesterol in chylomicrons and extremely large VLDL | NMR_metabo_UKBB | 0.1218 | 0.0436 | 5.19E-03 | 1.66E-02 |
| Tinnitus: Yes, but not now, but have in the past | eye_ear | 0.2833 | 0.1015 | 5.23E-03 | 1.67E-02 |
| Age started oral contraceptive pill | gynecology_fertility | -0.1532 | 0.0550 | 5.36E-03 | 1.70E-02 |
| Phospholipids in chylomicrons and extremely large VLDL | NMR_metabo_UKBB | 0.1236 | 0.0444 | 5.39E-03 | 1.71E-02 |
| Free cholesterol in large HDL | NMR_metabo_UKBB | -0.1264 | 0.0455 | 5.48E-03 | 1.73E-02 |
| Hearing difficulty/problems with background noise | eye_ear | 0.1060 | 0.0382 | 5.55E-03 | 1.74E-02 |

| Trait | Group | rg | se | p | p_fdr |
|---|---|---|---|---|---|
| Impedance of arm (right) | anthropometry | -0.0808 | 0.0292 | 5.59E-03 | 1.75E-02 |
| Ratio of docosahexaenoic acid to total fatty acids | NMR_metabo_UKBB | -0.1392 | 0.0503 | 5.70E-03 | 1.77E-02 |
| Concentration of chylomicrons and extremely large VLDL particles | NMR_metabo_UKBB | 0.1237 | 0.0448 | 5.72E-03 | 1.77E-02 |
| Free cholesterol in chylomicrons and extremely large VLDL | NMR_metabo_UKBB | 0.1236 | 0.0448 | 5.78E-03 | 1.78E-02 |
| Cholesteryl esters in chylomicrons and extremely large VLDL | NMR_metabo_UKBB | 0.1182 | 0.0430 | 6.00E-03 | 1.85E-02 |
| Impedance of whole body | anthropometry | -0.0793 | 0.0290 | 6.30E-03 | 1.93E-02 |
| Number of full brothers | family_memb_health | 0.1751 | 0.0642 | 6.37E-03 | 1.95E-02 |
| Diagnoses - main ICD10: J44 Other chronic obstructive pulmonary disease | diseases | 0.3455 | 0.1269 | 6.48E-03 | 1.97E-02 |
| Triglycerides to total lipids ratio in large HDL | NMR_metabo_UKBB | 0.1260 | 0.0464 | 6.66E-03 | 2.00E-02 |
| Fractured bone site(s): Other bones | pain_skeletal | 0.2133 | 0.0786 | 6.66E-03 | 2.00E-02 |
| Triglycerides to total lipids ratio in medium VLDL | NMR_metabo_UKBB | 0.1334 | 0.0492 | 6.67E-03 | 2.00E-02 |
| Fractured/broken bones in last 5 years | pain_skeletal | 0.1567 | 0.0579 | 6.79E-03 | 2.03E-02 |
| Diagnoses - main ICD10: K20 Oesophagitis | diseases | 0.3191 | 0.1182 | 6.94E-03 | 2.07E-02 |
| HOMA-B | biomarker | 0.2360 | 0.0875 | 6.97E-03 | 2.07E-02 |
| Creatinine (enzymatic) in urine | biomarker | 0.1117 | 0.0416 | 7.26E-03 | 2.14E-02 |
| Average weekly beer plus cider intake | alcohol | 0.1255 | 0.0468 | 7.27E-03 | 2.14E-02 |
| Diagnoses - main ICD10: K57 Diverticular disease of intestine | diseases | 0.2094 | 0.0781 | 7.36E-03 | 2.15E-02 |
| Reason for reducing amount of alcohol drunk: Financial reasons | alcohol | 0.2884 | 0.1076 | 7.37E-03 | 2.15E-02 |
| Number of live births | gynecology_fertility | 0.1322 | 0.0494 | 7.40E-03 | 2.15E-02 |
| Total lipids in chylomicrons and extremely large VLDL | NMR_metabo_UKBB | 0.1204 | 0.0451 | 7.62E-03 | 2.21E-02 |
| Total lipids in large HDL | NMR_metabo_UKBB | -0.1210 | 0.0454 | 7.71E-03 | 2.23E-02 |
| Duration of vigorous activity | activity_fitness_sleep | 0.1459 | 0.0551 | 8.14E-03 | 2.34E-02 |
| Types of physical activity in last 4 weeks: Heavy DIY (eg: weeding, lawn mowing, carpentry, digging) | activity_fitness_sleep | -0.1257 | 0.0476 | 8.24E-03 | 2.36E-02 |
| Number of cigarettes previously smoked daily | smoking | 0.1521 | 0.0576 | 8.27E-03 | 2.36E-02 |
| Triglycerides in medium HDL | NMR_metabo_UKBB | 0.1198 | 0.0455 | 8.47E-03 | 2.41E-02 |
| Triglycerides in very large VLDL | NMR_metabo_UKBB | 0.1119 | 0.0426 | 8.51E-03 | 2.42E-02 |
| Cholesteryl esters in HDL | NMR_metabo_UKBB | -0.1210 | 0.0461 | 8.64E-03 | 2.44E-02 |
| Average diameter for HDL particles | NMR_metabo_UKBB | -0.1209 | 0.0460 | 8.66E-03 | 2.44E-02 |
| Triglycerides in small LDL | NMR_metabo_UKBB | 0.1128 | 0.0430 | 8.72E-03 | 2.45E-02 |
| HDL cholesterol | NMR_metabo_UKBB | -0.1202 | 0.0463 | 9.45E-03 | 2.64E-02 |
| Number of full sisters | family_memb_health | 0.1804 | 0.0696 | 9.57E-03 | 2.66E-02 |
| Sodium in urine | biomarker | 0.0974 | 0.0376 | 9.60E-03 | 2.66E-02 |
| Reason for glasses/contact lenses: For short-sightedness, i.e. only or mainly for distance viewing such as driving, cinema etc (called 'myopia') | eye_ear | -0.1545 | 0.0598 | 9.74E-03 | 2.69E-02 |
| Diagnoses - main ICD10: K80 Cholelithiasis | diseases | 0.2226 | 0.0866 | 1.01E-02 | 2.79E-02 |

| Trait | Group | rg | se | p | p_fdr |
|---|---|---|---|---|---|
| Sleep duration | activity_fitness_sleep | -0.0913 | 0.0356 | 1.03E-02 | 2.82E-02 |
| ECG, heart rate | cardiovascular | -0.1487 | 0.0581 | 1.04E-02 | 2.86E-02 |
| Hearing difficulty/problems: Yes | eye_ear | 0.1338 | 0.0525 | 1.08E-02 | 2.94E-02 |
| Total triglycerides | NMR_metabo_UKBB | 0.1080 | 0.0424 | 1.08E-02 | 2.94E-02 |
| Cholesterol to total lipids ratio in small LDL | NMR_metabo_UKBB | -0.1639 | 0.0647 | 1.13E-02 | 3.04E-02 |
| Pain type(s) experienced in last month: Facial pain | pain_skeletal | 0.2445 | 0.0965 | 1.13E-02 | 3.04E-02 |
| Mouth/teeth dental problems: Painful gums | dental | 0.2180 | 0.0861 | 1.13E-02 | 3.04E-02 |
| Job involves shift work | work_stress | 0.1735 | 0.0686 | 1.14E-02 | 3.04E-02 |
| Ischemic stroke | diseases | 0.1660 | 0.0656 | 1.14E-02 | 3.04E-02 |
| Triglycerides to total lipids ratio in very large VLDL | NMR_metabo_UKBB | 0.1349 | 0.0534 | 1.15E-02 | 3.06E-02 |
| Concentration of very large VLDL particles | NMR_metabo_UKBB | 0.1074 | 0.0426 | 1.18E-02 | 3.11E-02 |
| Free cholesterol to total lipids ratio in medium VLDL | NMR_metabo_UKBB | -0.1244 | 0.0496 | 1.21E-02 | 3.21E-02 |
| Celiac disease | diseases | 0.2514 | 0.1006 | 1.24E-02 | 3.27E-02 |
| Total lipids in very large VLDL | NMR_metabo_UKBB | 0.1065 | 0.0429 | 1.30E-02 | 3.41E-02 |
| Smoking behaviors : Smoking cessation | smoking | -0.2297 | 0.0927 | 1.32E-02 | 3.45E-02 |
| Phospholipids in very large VLDL | NMR_metabo_UKBB | 0.1070 | 0.0432 | 1.33E-02 | 3.45E-02 |
| Types of transport used (excluding work): Public transport | activity_fitness_sleep | -0.1393 | 0.0563 | 1.33E-02 | 3.45E-02 |
| Triglycerides to total lipids ratio in small VLDL | NMR_metabo_UKBB | 0.1407 | 0.0569 | 1.34E-02 | 3.47E-02 |
| Snoring | activity_fitness_sleep | -0.0964 | 0.0391 | 1.37E-02 | 3.52E-02 |
| Alcohol drinker status: Current | alcohol | -0.1342 | 0.0545 | 1.37E-02 | 3.53E-02 |
| Parkinson's disease | neurology | -0.1428 | 0.0580 | 1.38E-02 | 3.53E-02 |
| Free cholesterol to total lipids ratio in very large VLDL | NMR_metabo_UKBB | -0.1230 | 0.0502 | 1.42E-02 | 3.62E-02 |
| Free cholesterol to total lipids ratio in small HDL | NMR_metabo_UKBB | -0.1405 | 0.0573 | 1.42E-02 | 3.62E-02 |
| Average diameter for LDL particles | NMR_metabo_UKBB | -0.1650 | 0.0674 | 1.44E-02 | 3.65E-02 |
| Cholesteryl esters to total lipids ratio in small HDL | NMR_metabo_UKBB | -0.1427 | 0.0585 | 1.47E-02 | 3.72E-02 |
| Triglycerides in HDL | NMR_metabo_UKBB | 0.1130 | 0.0466 | 1.53E-02 | 3.84E-02 |
| Triglycerides in chylomicrons and extremely large VLDL | NMR_metabo_UKBB | 0.1113 | 0.0459 | 1.53E-02 | 3.84E-02 |
| Time spent using computer | activity_fitness_sleep | -0.0791 | 0.0328 | 1.59E-02 | 3.97E-02 |
| Average weekly champagne plus white wine intake | alcohol | -0.1443 | 0.0603 | 1.66E-02 | 4.15E-02 |
| Average diameter for VLDL particles | NMR_metabo_UKBB | 0.1026 | 0.0430 | 1.69E-02 | 4.21E-02 |
| Difference in height between adolescence and adulthood | anthropometry | -0.3066 | 0.1284 | 1.70E-02 | 4.21E-02 |
| Phospholipids in large VLDL | NMR_metabo_UKBB | 0.1004 | 0.0422 | 1.73E-02 | 4.28E-02 |
| Tinnitus: Yes, now most or all of the time | eye_ear | 0.1964 | 0.0829 | 1.78E-02 | 4.38E-02 |
| Qualifications: NVQ or HND or HNC or equivalent | education_intelligence | 0.1424 | 0.0604 | 1.83E-02 | 4.50E-02 |
| Suffer from 'nerves' | mood_behaviour | 0.1018 | 0.0434 | 1.89E-02 | 4.63E-02 |
| Trunk fat-free mass | anthropometry | 0.0697 | 0.0297 | 1.91E-02 | 4.65E-02 |
| Phospholipids in large HDL | NMR_metabo_UKBB | -0.1085 | 0.0463 | 1.91E-02 | 4.65E-02 |
| Diagnoses - main ICD10: G56 Mononeuropathies of upper limb | diseases | 0.1587 | 0.0679 | 1.95E-02 | 4.72E-02 |
| Triglycerides in VLDL | NMR_metabo_UKBB | 0.0988 | 0.0425 | 2.03E-02 | 4.89E-02 |

| Trait | Group | rg | se | p | p_fdr |
|---|---|---|---|---|---|
| Free cholesterol in very large VLDL | NMR_metabo_UKBB | 0.1004 | 0.0433 | 2.03E-02 | 4.89E-02 |
| Isoleucine | NMR_metabo_UKBB | 0.1624 | 0.0702 | 2.07E-02 | 4.97E-02 |

**Table S4. List of traits tested in Mendelian randomization.**
After picking the key traits to be tested in Mendelian randomization (MR), we searched each trait on the IEU Open GWAS Project website (https://gwas.mrcieu.ac.uk/) and picked the GWAS with greatest sample size to use in MR. The table contains information for GWASes picked as it appears on the IEU Open GWAS Project website (24.1.2023). For some traits, we have included further information on trait in "Trait description" column; the descriptions are based on UKBB data notes of traits (https://biobank.ndph.ox.ac.uk/ukb/index.cgi) unless otherwise referenced.

| GWAS ID | Year | Trait | Consortium | Sample size | Number of SNPs | Trait description |
|---|---|---|---|---|---|---|
| ukb-b-6134 | 2018 | Age completed full time education | MRC-IEU | 307,897 | 9,851,867 | |
| ukb-b-5779 | 2018 | Alcohol intake frequency. | MRC-IEU | 462,346 | 9,851,867 | |
| ieu-b-107 | 2020 | apolipoprotein A-I | UK Biobank | 393,193 | 12,321,875 | |
| ukb-b-11411 | 2018 | Blood clot, DVT, bronchitis, emphysema, asthma, rhinitis, eczema, allergy diagnosed by doctor: Blood clot in the leg (DVT) | MRC-IEU | 462,013 | 9,851,867 | |
| ukb-b-13704 | 2018 | Blood clot, DVT, bronchitis, emphysema, asthma, rhinitis, eczema, allergy diagnosed by doctor: Blood clot in the lung | MRC-IEU | 462,013 | 9,851,867 | |
| ukb-b-16207 | 2018 | Blood clot, DVT, bronchitis, emphysema, asthma, rhinitis, eczema, allergy diagnosed by doctor: Emphysema/chronic bronchitis | MRC-IEU | 462,013 | 9,851,867 | |
| ukb-b-19953 | 2018 | Body mass index (BMI) | MRC-IEU | 461,460 | 9,851,867 | |
| ieu-a-1058 | 2011 | Celiac disease | NA | 24,269 | 38,037 | |
| ieu-a-836 | 2013 | College completion | SSGAC | 95,427 | 2,321,511 | |
| ebi-a-GCST005195 | 2017 | Coronary artery disease | NA | 547,261 | 7,934,254 | |
| ukb-a-333 | 2017 | Creatinine (enzymatic) in urine | Neale Lab | 327,525 | 10,894,596 | |
| ukb-b-223 | 2018 | Current tobacco smoking | MRC-IEU | 462,434 | 9,851,867 | |

| | | | | | | |
|---|---|---|---|---|---|---|
| ieu-a-1000 | 2016 | Depressive symptoms | SSGAC | 161,460 | 6,524,475 | Meta-analysis of two major depressive disorder GWASes (Psychiatric Genomics Consortium and Resource for Genetic Epidemiology Research on Aging cohort) and UKBB continous phenotype constructed by combining answers to two questions asking about experiencing feelings of unenthusiasim or disinterest and depression or hopelessness[1]. |
| ukb-b-10753 | 2018 | Diabetes diagnosed by doctor | MRC-IEU | 461,578 | 9,851,867 | |
| ukb-a-528 | 2017 | Diagnoses - main ICD10: G56 Mononeuropathies of upper limb | Neale Lab | 337,199 | 10,894,596 | |
| ukb-a-532 | 2017 | Diagnoses - main ICD10: I20 Angina pectoris | Neale Lab | 337,199 | 10,894,596 | |
| ukb-a-534 | 2017 | Diagnoses - main ICD10: I25 Chronic ischaemic heart disease | Neale Lab | 337,199 | 10,894,596 | |
| ukb-a-543 | 2017 | Diagnoses - main ICD10: J44 Other chronic obstructive pulmonary disease | Neale Lab | 337,199 | 10,894,596 | |
| ukb-b-19354 | 2018 | Diagnoses - main ICD10: K20 Oesophagitis | MRC-IEU | 463,010 | 9,851,867 | |
| ukb-a-545 | 2017 | Diagnoses - main ICD10: K21 Gastro-oesophageal reflux disease | Neale Lab | 337,199 | 10,894,596 | |
| ukb-a-554 | 2017 | Diagnoses - main ICD10: K52 Other non-infective gastro-enteritis and colitis | Neale Lab | 337,199 | 10,894,596 | |
| ukb-a-555 | 2017 | Diagnoses - main ICD10: K57 Diverticular disease of intestine | Neale Lab | 337,199 | 10,894,596 | |
| ukb-a-559 | 2017 | Diagnoses - main ICD10: K80 Cholelithiasis | Neale Lab | 337,199 | 10,894,596 | |
| ukb-d-M25 | 2018 | Diagnoses - main ICD10: M25 Other joint disorders, not elsewhere classified | NA | 361,194 | 12,784,029 | |
| ukb-a-581 | 2017 | Diagnoses - main ICD10: R07 Pain in throat and chest | Neale Lab | 337,199 | 10,894,596 | |
| ukb-a-582 | 2017 | Diagnoses - main ICD10: R10 Abdominal and pelvic pain | Neale Lab | 337,199 | 10,894,596 | |

| | | | | | | |
|---|---|---|---|---|---|---|
| ukb-b-11874 | 2018 | Diagnoses - secondary ICD10: M06.99 Rheumatoid arthritis, unspecified (Site unspecified) | MRC-IEU | 463,010 | 9,851,867 | |
| ukb-b-11361 | 2018 | ECG, heart rate | MRC-IEU | 68,160 | 9,851,867 | |
| ukb-b-17001 | 2018 | Ever had bowel cancer screening | MRC-IEU | 455,259 | 9,851,867 | |
| ukb-b-10181 | 2018 | Eye problems/disorders: Diabetes related eye disease | MRC-IEU | 150,642 | 9,851,867 | |
| ukb-b-19809 | 2018 | Fed-up feelings | MRC-IEU | 453,071 | 9,851,867 | Participants were asked "Do you often feel "fed-up"? (yes, no, do not know, prefer not to answer) |
| ukb-b-5238 | 2018 | Fluid intelligence score | MRC-IEU | 149,051 | 9,851,867 | Sum of correct answers given to 13 different questions measuring fluid intelligence. |
| ukb-b-19657 | 2018 | Forced expiratory volume in 1-second (FEV1) | MRC-IEU | 421,986 | 9,851,867 | |
| ukb-b-11141 | 2018 | Forced expiratory volume in 1-second (FEV1), Best measure | MRC-IEU | 345,665 | 9,851,867 | |
| ukb-b-13405 | 2018 | Forced expiratory volume in 1-second (FEV1), predicted percentage | MRC-IEU | 148,653 | 9,851,867 | |
| ukb-b-7953 | 2018 | Forced vital capacity (FVC) | MRC-IEU | 421,986 | 9,851,867 | |
| ukb-b-14713 | 2018 | Forced vital capacity (FVC), Best measure | MRC-IEU | 345,665 | 9,851,867 | |
| ukb-b-3822 | 2018 | Frequency of depressed mood in last 2 weeks | MRC-IEU | 442,840 | 9,851,867 | |
| ukb-b-5664 | 2018 | Frequency of tenseness / restlessness in last 2 weeks | MRC-IEU | 445,194 | 9,851,867 | |
| ukb-b-929 | 2018 | Frequency of tiredness / lethargy in last 2 weeks | MRC-IEU | 449,019 | 9,851,867 | |
| ukb-b-1419 | 2018 | Frequency of unenthusiasm / disinterest in last 2 weeks | MRC-IEU | 447,403 | 9,851,867 | |
| ukb-b-15378 | 2018 | Had other major operations | MRC-IEU | 248,845 | 9,851,867 | |
| ieu-b-109 | 2020 | HDL cholesterol | UK Biobank | 403,943 | 12,321,875 | |
| ukb-b-18275 | 2018 | Hearing difficulty/problems with background noise | MRC-IEU | 453,482 | 9,851,867 | |
| ukb-a-257 | 2017 | Hearing difficulty/problems: Yes | Neale Lab | 323,978 | 10,894,596 | |

| | | | | | | |
|---|---|---|---|---|---|---|
| ieu-b-117 | 2011 | HOMA-B | MAGIC | 36,466 | 2,454,220 | Homeostasis model assesment of beta-cell function from paired fasting glucose measurement[2] |
| ieu-b-118 | 2011 | HOMA-IR | MAGIC | 37,037 | 2,455,342 | Homeostasis model assesment of insulin resistance from insulin measurement[2] |
| ebi-a-GCST004131 | 2017 | Inflammatory bowel disease | NA | 59,957 | 9,619,016 | |
| ebi-a-GCST006250 | 2018 | Intelligence | NA | 269,867 | 9,276,181 | Meta-analysis of 14 cohorts with varying intelligence phenotypes, including e.g. verbal and mathematical reasoning, neuropsychological tests and memory[3]. |
| ebi-a-GCST005843 | 2018 | Ischemic stroke | NA | 440,328 | 7,537,579 | |
| ukb-b-8476 | 2018 | Loneliness, isolation | MRC-IEU | 455,364 | 9,851,867 | Participants were asked "Do you often feel lonely?" (yes, no, do not know, prefer not to answer) |
| ukb-b-13764 | 2018 | Long-standing illness, disability or infirmity | MRC-IEU | 451,893 | 9,851,867 | Participants were asked "Do you have any long-standing illness, disability or infirmity?" (yes, no, do not know, prefer not to answer) |
| ukb-b-5436 | 2018 | Loud music exposure frequency | MRC-IEU | 150,898 | 9,851,867 | |
| ebi-a-GCST005903 | 2018 | Major depressive disorder (ICD-10 coded) | NA | 217,584 | 7,640,987 | |
| ukb-b-14461 | 2018 | Maximum heart rate during fitness test | MRC-IEU | 68,409 | 9,851,867 | |
| ukb-b-18994 | 2018 | Miserableness | MRC-IEU | 454,982 | 9,851,867 | |
| ukb-b-14180 | 2018 | Mood swings | MRC-IEU | 451,619 | 9,851,867 | |
| ebi-a-GCST005232 | 2017 | Neuroticism | NA | 329,821 | 18,436,568 | Neuroticism was measured by total score on the 12-item Eysenck Personality Questionnaire–Revised Short Form (EPQ-R-S)[4]. |

| | | | | | | |
|---|---|---|---|---|---|---|
| ukb-b-4630 | 2018 | Neuroticism score | MRC-IEU | 374,323 | 9,851,867 | Neuroticism score is based on a questionnaire with 12 domains of neurotic behaviour. This variable has come from Professor Jill Pell from the Institute of Health & Wellbeing, University of Glasgow. |
| ukb-b-4733 | 2018 | Number of operations, self-reported | MRC-IEU | 462,933 | 9,851,867 | |
| ukb-b-14961 | 2018 | Other serious medical condition/disability diagnosed by doctor | MRC-IEU | 454,253 | 9,851,867 | |
| ieu-b-7 | 2019 | Parkinson's disease | International Parkinson's Disease Genomics Consortium | 482,730 | 17,891,936 | |
| ukb-b-11971 | 2018 | Pulse wave Arterial Stiffness index | MRC-IEU | 151,053 | 9,851,867 | |
| ukb-b-8778 | 2018 | Pulse wave peak to peak time | MRC-IEU | 151,466 | 9,851,867 | |
| ukb-b-11615 | 2018 | Qualifications: A levels/AS levels or equivalent | MRC-IEU | 458,079 | 9,851,867 | Answer to question "Which of the following qualifications do you have? (You can select more than one)" |
| ukb-b-16489 | 2018 | Qualifications: College or University degree | MRC-IEU | 458,079 | 9,851,867 | Answer to question "Which of the following qualifications do you have? (You can select more than one)" |
| ukb-b-10505 | 2018 | Qualifications: CSEs or equivalent | MRC-IEU | 458,079 | 9,851,867 | Answer to question "Which of the following qualifications do you have? (You can select more than one)" |
| ukb-b-17729 | 2018 | Qualifications: None of the above | MRC-IEU | 458,079 | 9,851,867 | Answer to question "Which of the following qualifications do you have? (You can select more than one)" |
| ukb-b-7926 | 2018 | Qualifications: NVQ or HND or HNC or equivalent | MRC-IEU | 458,079 | 9,851,867 | Answer to question "Which of the following qualifications do you have? (You can select more than one)" |

| | | | | | | |
|---|---|---|---|---|---|---|
| ukb-b-18099 | 2018 | Qualifications: O levels/GCSEs or equivalent | MRC-IEU | 458,079 | 9,851,867 | Answer to question "Which of the following qualifications do you have? (You can select more than one)" |
| ukb-b-13799 | 2018 | Qualifications: Other professional qualifications eg: nursing, teaching | MRC-IEU | 458,079 | 9,851,867 | Answer to question "Which of the following qualifications do you have? (You can select more than one)" |
| ukb-b-6353 | 2018 | Reason for glasses/contact lenses: For short-sightedness, i.e. only or mainly for distance viewing such as driving, cinema etc (called 'myopia') | MRC-IEU | 460,536 | 9,851,867 | |
| ukb-b-18336 | 2018 | Seen a psychiatrist for nerves, anxiety, tension or depression | MRC-IEU | 460,702 | 9,851,867 | |
| ukb-b-6991 | 2018 | Seen doctor (GP) for nerves, anxiety, tension or depression | MRC-IEU | 459,560 | 9,851,867 | |
| ukb-b-9981 | 2018 | Sensitivity / hurt feelings | MRC-IEU | 449,419 | 9,851,867 | Participants were asked "Are your feelings easily hurt?" (yes, no, do not know, prefer not to answer) |
| ukb-b-3957 | 2018 | Sleeplessness / insomnia | MRC-IEU | 462,341 | 9,851,867 | |
| ukb-a-335 | 2017 | Sodium in urine | Neale Lab | 326,831 | 10,894,596 | |
| ukb-b-19957 | 2018 | Suffer from 'nerves' | MRC-IEU | 445,809 | 9,851,867 | Participants were asked "Do you suffer from 'nerves'?" (yes, no, do not know, prefer not to answer) |
| ukb-d-4803_14 | 2018 | Tinnitus: Yes, but not now, but have in the past | NA | 117,882 | 13,567,874 | |
| ukb-d-4803_11 | 2018 | Tinnitus: Yes, now most or all of the time | NA | 117,882 | 12,817,923 | |
| ieu-b-111 | 2020 | triglycerides | UK Biobank | 441,016 | 12,321,875 | |
| ebi-a-GCST006867 | 2018 | Type 2 diabetes | NA | 655,666 | 5,030,727 | |
| ukb-b-8468 | 2018 | Vascular/heart problems diagnosed by doctor: Angina | MRC-IEU | 461,880 | 9,851,867 | |
| ukb-b-14177 | 2018 | Vascular/heart problems diagnosed by doctor: High blood pressure | MRC-IEU | 461,880 | 9,851,867 | |

| | | | | | |
|---|---|---|---|---|---|
| ukb-b-13352 | 2018 | Vascular/heart problems diagnosed by doctor: None of the above | MRC-IEU | 461,880 | 9,851,867 |
| ieu-a-72 | 2015 | Waist-to-hip ratio | GIANT | 224,459 | 2,562,516 |
| ukb-b-18335 | 2018 | Wheeze or whistling in the chest in last year | MRC-IEU | 453,959 | 9,851,867 |
| ukb-b-19393 | 2018 | Whole body fat mass | MRC-IEU | 454,137 | 9,851,867 |
| ukb-b-13354 | 2018 | Whole body fat-free mass | MRC-IEU | 454,850 | 9,851,867 |
| ieu-a-1239 | 2018 | Years of schooling | SSGAC | 766,345 | 10,101,242 |

**Table S5. Mendelian randomization results (exposure -> psoriasis).**
Columns: nsnp, number of valid SNPs used for analysis; pval, p-value; or, odds ratio; or lci95; odds ratio lower 95 % confidence interval; or uci95, odds ratio upper 95 % confidence interval; Q pval, Q-test p-value (for testing heterogeneity); egger intercept, MR Egger intercept; and ple pval, pleiotropy p-value.

| exposure | method | nsnp | pval | or | or lci95 | or uci95 | Q pval | egger intercept | ple pval |
|---|---|---|---|---|---|---|---|---|---|
| Age completed full time education \|\| id:ukb-b-6134 | MR Egger | 36 | 3.82E-01 | 3.04E+00 | 2.59E-01 | 3.57E+01 | 2.53E-02 | -0.0279 | 0.1319 |
| Age completed full time education \|\| id:ukb-b-6134 | Inverse variance weighted | 36 | 6.80E-03 | 4.59E-01 | 2.62E-01 | 8.07E-01 | 1.50E-02 | | |
| Alcohol intake frequency. \|\| id:ukb-b-5779 | MR Egger | 88 | 3.70E-01 | 1.85E+00 | 4.85E-01 | 7.08E+00 | 3.09E-64 | -0.0043 | 0.7893 |
| Alcohol intake frequency. \|\| id:ukb-b-5779 | Inverse variance weighted | 88 | 4.47E-02 | 1.56E+00 | 1.01E+00 | 2.40E+00 | 6.39E-64 | | |
| apolipoprotein A-I \|\| id:ieu-b-107 | MR Egger | 250 | 6.12E-01 | 1.04E+00 | 8.84E-01 | 1.23E+00 | 6.33E-06 | -0.0040 | 0.1224 |
| apolipoprotein A-I \|\| id:ieu-b-107 | Inverse variance weighted | 250 | 2.65E-01 | 9.42E-01 | 8.49E-01 | 1.05E+00 | 4.38E-06 | | |
| Blood clot, DVT, bronchitis, emphysema, asthma, rhinitis, eczema, allergy diagnosed by doctor: Blood clot in the leg (DVT) \|\| id:ukb-b-11411 | MR Egger | 11 | 3.26E-01 | 2.43E-02 | 2.18E-05 | 2.70E+01 | 1.66E-01 | 0.0101 | 0.4737 |
| Blood clot, DVT, bronchitis, emphysema, asthma, rhinitis, eczema, allergy diagnosed by doctor: Blood clot in the leg (DVT) \|\| id:ukb-b-11411 | Inverse variance weighted | 11 | 4.48E-01 | 2.17E-01 | 4.18E-03 | 1.12E+01 | 1.86E-01 | | |
| Blood clot, DVT, bronchitis, emphysema, asthma, rhinitis, eczema, allergy diagnosed by doctor: Blood clot in the lung \|\| id:ukb-b-13704 | MR Egger | 5 | 8.24E-01 | 4.26E-05 | 1.92E-40 | 9.47E+30 | 1.08E-01 | 0.0286 | 0.7233 |
| Blood clot, DVT, bronchitis, emphysema, asthma, rhinitis, eczema, allergy diagnosed by doctor: Blood clot in the lung \|\| id:ukb-b-13704 | Inverse variance weighted | 5 | 3.75E-01 | 3.39E+02 | 8.65E-04 | 1.33E+08 | 1.72E-01 | | |

| exposure | method | nsnp | pval | or | or lci95 | or uci95 | Q pval | egger intercept | ple pval |
|---|---|---|---|---|---|---|---|---|---|
| Body mass index (BMI) \|\| id:ukb-b-19953 | MR Egger | 398 | 5.73E-03 | 1.68E+00 | 1.17E+00 | 2.43E+00 | 2.51E-14 | -0.0021 | 0.5377 |
| Body mass index (BMI) \|\| id:ukb-b-19953 | Inverse variance weighted | 398 | 4.51E-09 | 1.51E+00 | 1.32E+00 | 1.73E+00 | 2.85E-14 | | |
| Celiac disease \|\| id:ieu-a-1058 | MR Egger | 15 | 6.77E-01 | 1.02E+00 | 9.31E-01 | 1.12E+00 | 4.55E-11 | 0.0089 | 0.6638 |
| Celiac disease \|\| id:ieu-a-1058 | Inverse variance weighted | 15 | 2.49E-01 | 1.04E+00 | 9.75E-01 | 1.10E+00 | 6.96E-11 | | |
| College completion \|\| id:ieu-a-836 | MR Egger | 3 | 3.81E-01 | 7.42E-02 | 2.29E-03 | 2.41E+00 | 7.69E-01 | 0.1587 | 0.4284 |
| College completion \|\| id:ieu-a-836 | Inverse variance weighted | 3 | 1.64E-02 | 6.82E-01 | 4.99E-01 | 9.32E-01 | 4.36E-01 | | |
| Coronary artery disease \|\| id:ebi-a-GCST005195 | MR Egger | 60 | 3.22E-02 | 1.22E+00 | 1.02E+00 | 1.45E+00 | 9.29E-02 | -0.0111 | 0.0979 |
| Coronary artery disease \|\| id:ebi-a-GCST005195 | Inverse variance weighted | 60 | 1.39E-01 | 1.07E+00 | 9.79E-01 | 1.16E+00 | 6.51E-02 | | |
| Creatinine (enzymatic) in urine \|\| id:ukb-a-333 | MR Egger | 21 | 1.75E-01 | 2.09E+02 | 1.24E-01 | 3.51E+05 | 5.93E-46 | -0.0821 | 0.2736 |
| Creatinine (enzymatic) in urine \|\| id:ukb-a-333 | Inverse variance weighted | 21 | 1.96E-01 | 3.30E+00 | 5.39E-01 | 2.02E+01 | 5.12E-49 | | |
| Current tobacco smoking \|\| id:ukb-b-223 | MR Egger | 33 | 4.85E-01 | 4.05E+00 | 8.38E-02 | 1.95E+02 | 2.26E-02 | -0.0098 | 0.5506 |
| Current tobacco smoking \|\| id:ukb-b-223 | Inverse variance weighted | 33 | 6.30E-01 | 1.28E+00 | 4.72E-01 | 3.46E+00 | 2.62E-02 | | |
| Diabetes diagnosed by doctor \|\| id:ukb-b-10753 | MR Egger | 62 | 9.10E-01 | 8.58E-01 | 6.13E-02 | 1.20E+01 | 4.68E-01 | 0.0026 | 0.6726 |
| Diabetes diagnosed by doctor \|\| id:ukb-b-10753 | Inverse variance weighted | 62 | 5.29E-01 | 1.44E+00 | 4.64E-01 | 4.45E+00 | 4.98E-01 | | |
| Diagnoses - main ICD10: G56 Mononeuropathies of upper limb \|\| id:ukb-a-528 | MR Egger | 5 | 9.93E-01 | 1.18E+00 | 1.28E-14 | 1.08E+14 | 3.57E-01 | -0.0137 | 0.7797 |
| Diagnoses - main ICD10: G56 Mononeuropathies of upper limb \|\| id:ukb-a-528 | Inverse variance weighted | 5 | 2.74E-01 | 9.39E-03 | 2.20E-06 | 4.01E+01 | 5.04E-01 | | |
| Diagnoses - main ICD10: I20 Angina pectoris \|\| id:ukb-a-532 | Inverse variance weighted | 2 | 8.98E-01 | 2.25E+00 | 1.00E-05 | 5.04E+05 | 5.27E-01 | NA | NA |

| exposure | method | nsnp | pval | or | or lci95 | or uci95 | Q pval | egger intercept | ple pval |
|---|---|---|---|---|---|---|---|---|---|
| Diagnoses - main ICD10: I25 Chronic ischaemic heart disease \|\| id:ukb-a-534 | MR Egger | 15 | 3.23E-01 | 4.38E+01 | 3.21E-02 | 5.97E+04 | 1.84E-01 | -0.0122 | 0.4156 |
| Diagnoses - main ICD10: I25 Chronic ischaemic heart disease \|\| id:ukb-a-534 | Inverse variance weighted | 15 | 5.42E-01 | 2.85E+00 | 9.88E-02 | 8.22E+01 | 1.94E-01 | | |
| Diagnoses - main ICD10: J44 Other chronic obstructive pulmonary disease \|\| id:ukb-a-543 | Inverse variance weighted | 2 | 5.40E-01 | 2.12E-04 | 3.66E-16 | 1.23E+08 | 9.53E-01 | NA | NA |
| Diagnoses - main ICD10: K57 Diverticular disease of intestine \|\| id:ukb-a-555 | MR Egger | 3 | 4.14E-01 | 3.19E+03 | 1.91E-02 | 5.33E+08 | 6.07E-01 | -0.0236 | 0.4923 |
| Diagnoses - main ICD10: K57 Diverticular disease of intestine \|\| id:ukb-a-555 | Inverse variance weighted | 3 | 4.08E-01 | 1.88E+01 | 1.81E-02 | 1.96E+04 | 5.18E-01 | | |
| Diagnoses - main ICD10: K80 Cholelithiasis \|\| id:ukb-a-559 | MR Egger | 10 | 5.02E-01 | 1.49E-01 | 7.33E-04 | 3.03E+01 | 5.79E-02 | 0.0030 | 0.8161 |
| Diagnoses - main ICD10: K80 Cholelithiasis \|\| id:ukb-a-559 | Inverse variance weighted | 10 | 4.22E-01 | 2.38E-01 | 7.11E-03 | 7.93E+00 | 8.63E-02 | | |
| Diagnoses - secondary ICD10: M06.99 Rheumatoid arthritis, unspecified (Site unspecified) \|\| id:ukb-b-11874 | Inverse variance weighted | 2 | 7.90E-01 | 1.18E+01 | 1.53E-07 | 9.15E+08 | 2.51E-01 | NA | NA |
| ECG, heart rate \|\| id:ukb-b-11361 | MR Egger | 9 | 8.03E-01 | 1.24E+00 | 2.42E-01 | 6.36E+00 | 3.25E-06 | -0.0164 | 0.7289 |
| ECG, heart rate \|\| id:ukb-b-11361 | Inverse variance weighted | 9 | 8.43E-01 | 9.41E-01 | 5.15E-01 | 1.72E+00 | 5.96E-06 | | |
| Eye problems/disorders: Diabetes related eye disease \|\| id:ukb-b-10181 | Inverse variance weighted | 2 | 1.07E-01 | 9.51E-05 | 1.21E-09 | 7.51E+00 | 3.97E-02 | NA | NA |
| Fed-up feelings \|\| id:ukb-b-19809 | MR Egger | 51 | 6.21E-01 | 2.44E+00 | 7.29E-02 | 8.15E+01 | 9.33E-01 | -0.0003 | 0.9803 |
| Fed-up feelings \|\| id:ukb-b-19809 | Inverse variance weighted | 51 | 2.42E-02 | 2.33E+00 | 1.12E+00 | 4.88E+00 | 9.46E-01 | | |
| Fluid intelligence score \|\| id:ukb-b-5238 | MR Egger | 69 | 4.57E-01 | 8.38E-01 | 5.27E-01 | 1.33E+00 | 8.51E-02 | 0.0047 | 0.7078 |
| Fluid intelligence score \|\| id:ukb-b-5238 | Inverse variance weighted | 69 | 6.53E-02 | 9.14E-01 | 8.31E-01 | 1.01E+00 | 9.66E-02 | | |
| Forced expiratory volume in 1-second (FEV1) \|\| id:ukb-b-19657 | MR Egger | 234 | 3.44E-01 | 1.32E+00 | 7.43E-01 | 2.35E+00 | 3.49E-02 | -0.0080 | 0.0734 |
| Forced expiratory volume in 1-second (FEV1) \|\| id:ukb-b-19657 | Inverse variance weighted | 234 | 2.27E-02 | 8.02E-01 | 6.64E-01 | 9.70E-01 | 2.71E-02 | | |
| Forced expiratory volume in 1-second (FEV1), Best measure \|\| id:ukb-b-11141 | MR Egger | 204 | 5.16E-01 | 1.23E+00 | 6.59E-01 | 2.29E+00 | 1.48E-03 | -0.0083 | 0.1140 |

| exposure | method | nsnp | pval | or | or lci95 | or uci95 | Q pval | egger intercept | ple pval |
|---|---|---|---|---|---|---|---|---|---|
| Forced expiratory volume in 1-second (FEV1), Best measure \|\| id:ukb-b-11141 | Inverse variance weighted | 204 | 6.75E-03 | 7.62E-01 | 6.25E-01 | 9.27E-01 | 1.10E-03 | | |
| Forced expiratory volume in 1-second (FEV1), predicted percentage \|\| id:ukb-b-13405 | MR Egger | 80 | 1.82E-01 | 1.82E+00 | 7.60E-01 | 4.38E+00 | 2.60E-02 | -0.0189 | 0.1149 |
| Forced expiratory volume in 1-second (FEV1), predicted percentage \|\| id:ukb-b-13405 | Inverse variance weighted | 80 | 3.21E-01 | 9.09E-01 | 7.54E-01 | 1.10E+00 | 1.84E-02 | | |
| Forced vital capacity (FVC) \|\| id:ukb-b-7953 | MR Egger | 283 | 6.48E-01 | 1.12E+00 | 6.95E-01 | 1.79E+00 | 5.86E-03 | -0.0050 | 0.1791 |
| Forced vital capacity (FVC) \|\| id:ukb-b-7953 | Inverse variance weighted | 283 | 3.02E-02 | 8.25E-01 | 6.93E-01 | 9.82E-01 | 5.21E-03 | | |
| Forced vital capacity (FVC), Best measure \|\| id:ukb-b-14713 | MR Egger | 263 | 7.74E-01 | 1.07E+00 | 6.75E-01 | 1.70E+00 | 2.43E-03 | -0.0029 | 0.4636 |
| Forced vital capacity (FVC), Best measure \|\| id:ukb-b-14713 | Inverse variance weighted | 263 | 2.86E-01 | 9.11E-01 | 7.69E-01 | 1.08E+00 | 2.55E-03 | | |
| Frequency of depressed mood in last 2 weeks \|\| id:ukb-b-3822 | MR Egger | 11 | 3.98E-01 | 1.38E+02 | 2.58E-03 | 7.39E+06 | 6.80E-01 | -0.0398 | 0.4300 |
| Frequency of depressed mood in last 2 weeks \|\| id:ukb-b-3822 | Inverse variance weighted | 11 | 5.73E-01 | 1.45E+00 | 4.00E-01 | 5.23E+00 | 6.99E-01 | | |
| Frequency of tenseness / restlessness in last 2 weeks \|\| id:ukb-b-5664 | MR Egger | 15 | 9.23E-04 | 2.62E+04 | 2.44E+02 | 2.81E+06 | 5.80E-01 | -0.0832 | 0.0016 |
| Frequency of tenseness / restlessness in last 2 weeks \|\| id:ukb-b-5664 | Inverse variance weighted | 15 | 2.27E-01 | 2.76E+00 | 5.33E-01 | 1.43E+01 | 1.87E-02 | | |
| Frequency of tiredness / lethargy in last 2 weeks \|\| id:ukb-b-929 | MR Egger | 38 | 6.57E-02 | 1.20E+01 | 9.22E-01 | 1.56E+02 | 4.13E-01 | -0.0175 | 0.2647 |
| Frequency of tiredness / lethargy in last 2 weeks \|\| id:ukb-b-929 | Inverse variance weighted | 38 | 1.37E-04 | 2.81E+00 | 1.65E+00 | 4.79E+00 | 4.00E-01 | | |
| Frequency of unenthusiasm / disinterest in last 2 weeks \|\| id:ukb-b-1419 | MR Egger | 13 | 1.94E-01 | 3.50E+11 | 1.57E-05 | 7.78E+27 | 1.44E-77 | -0.1984 | 0.2712 |
| Frequency of unenthusiasm / disinterest in last 2 weeks \|\| id:ukb-b-1419 | Inverse variance weighted | 13 | 2.07E-01 | 1.16E+02 | 7.27E-02 | 1.84E+05 | 5.84E-87 | | |
| Had other major operations \|\| id:ukb-b-15378 | Inverse variance weighted | 2 | 9.33E-01 | 1.22E+00 | 1.08E-02 | 1.39E+02 | 5.95E-02 | NA | NA |

| exposure | method | nsnp | pval | or | or lci95 | or uci95 | Q pval | egger intercept | ple pval |
|---|---|---|---|---|---|---|---|---|---|
| HDL cholesterol \|\| id:ieu-b-109 | MR Egger | 298 | 1.51E-01 | 8.94E-01 | 7.67E-01 | 1.04E+00 | 2.20E-05 | 0.0006 | 0.7767 |
| HDL cholesterol \|\| id:ieu-b-109 | Inverse variance weighted | 298 | 5.96E-02 | 9.09E-01 | 8.23E-01 | 1.00E+00 | 2.56E-05 | | |
| Hearing difficulty/problems with background noise \|\| id:ukb-b-18275 | MR Egger | 30 | 1.21E-01 | 2.12E-03 | 1.11E-06 | 4.04E+00 | 8.47E-03 | 0.0498 | 0.0764 |
| Hearing difficulty/problems with background noise \|\| id:ukb-b-18275 | Inverse variance weighted | 30 | 2.17E-01 | 2.30E+00 | 6.12E-01 | 8.63E+00 | 2.58E-03 | | |
| Hearing difficulty/problems: Yes \|\| id:ukb-a-257 | MR Egger | 22 | 5.53E-01 | 3.09E+00 | 7.94E-02 | 1.20E+02 | 4.52E-01 | 0.0004 | 0.9765 |
| Hearing difficulty/problems: Yes \|\| id:ukb-a-257 | Inverse variance weighted | 22 | 2.28E-02 | 3.26E+00 | 1.18E+00 | 9.01E+00 | 5.15E-01 | | |
| HOMA-B \|\| id:ieu-b-117 | Inverse variance weighted | 2 | 9.38E-01 | 9.64E-01 | 3.81E-01 | 2.44E+00 | 2.61E-01 | NA | NA |
| Inflammatory bowel disease \|\| id:ebi-a-GCST004131 | MR Egger | 94 | 3.22E-01 | 1.05E+00 | 9.55E-01 | 1.15E+00 | 3.59E-31 | 0.0091 | 0.1826 |
| Inflammatory bowel disease \|\| id:ebi-a-GCST004131 | Inverse variance weighted | 94 | 4.19E-04 | 1.11E+00 | 1.05E+00 | 1.17E+00 | 5.65E-32 | | |
| Intelligence \|\| id:ebi-a-GCST006250 | MR Egger | 137 | 1.11E-01 | 2.00E+00 | 8.57E-01 | 4.67E+00 | 2.63E-01 | -0.0177 | 0.0401 |
| Intelligence \|\| id:ebi-a-GCST006250 | Inverse variance weighted | 137 | 3.26E-02 | 8.31E-01 | 7.01E-01 | 9.85E-01 | 2.01E-01 | | |
| Ischemic stroke \|\| id:ebi-a-GCST005843 | MR Egger | 16 | 8.62E-01 | 1.14E+00 | 2.61E-01 | 5.00E+00 | 1.70E-05 | -0.0026 | 0.9576 |
| Ischemic stroke \|\| id:ebi-a-GCST005843 | Inverse variance weighted | 16 | 4.95E-01 | 1.10E+00 | 8.40E-01 | 1.43E+00 | 3.25E-05 | | |
| Loneliness, isolation \|\| id:ukb-b-8476 | MR Egger | 16 | 9.55E-01 | 1.27E+00 | 3.97E-04 | 4.06E+03 | 6.85E-01 | 0.0043 | 0.8485 |
| Loneliness, isolation \|\| id:ukb-b-8476 | Inverse variance weighted | 16 | 2.56E-01 | 2.77E+00 | 4.77E-01 | 1.61E+01 | 7.49E-01 | | |
| Long-standing illness, disability or infirmity \|\| id:ukb-b-13764 | MR Egger | 28 | 5.59E-01 | 4.78E+00 | 2.70E-02 | 8.48E+02 | 5.53E-02 | -0.0029 | 0.8746 |
| Long-standing illness, disability or infirmity \|\| id:ukb-b-13764 | Inverse variance weighted | 28 | 6.63E-02 | 3.18E+00 | 9.25E-01 | 1.09E+01 | 7.09E-02 | | |
| Maximum heart rate during fitness test \|\| id:ukb-b-14461 | MR Egger | 5 | 1.39E-01 | 1.50E-01 | 2.35E-02 | 9.61E-01 | 1.84E-01 | 0.1092 | 0.1158 |
| Maximum heart rate during fitness test \|\| id:ukb-b-14461 | Inverse variance weighted | 5 | 7.68E-01 | 1.11E+00 | 5.52E-01 | 2.23E+00 | 1.34E-02 | | |
| Miserableness \|\| id:ukb-b-18994 | MR Egger | 39 | 2.63E-01 | 1.53E+01 | 1.39E-01 | 1.70E+03 | 2.61E-01 | -0.0134 | 0.4453 |
| Miserableness \|\| id:ukb-b-18994 | Inverse variance weighted | 39 | 4.90E-02 | 2.49E+00 | 1.00E+00 | 6.19E+00 | 2.75E-01 | | |

| exposure | method | nsnp | pval | or | or lci95 | or uci95 | Q pval | egger intercept | ple pval |
|---|---|---|---|---|---|---|---|---|---|
| Mood swings \|\| id:ukb-b-14180 | MR Egger | 54 | 2.12E-01 | 8.36E+01 | 8.70E-02 | 8.03E+04 | 4.14E-08 | -0.0273 | 0.2873 |
| Mood swings \|\| id:ukb-b-14180 | Inverse variance weighted | 54 | 2.12E-01 | 2.03E+00 | 6.67E-01 | 6.18E+00 | 2.79E-08 | | |
| Neuroticism \|\| id:ebi-a-GCST005232 | MR Egger | 66 | 9.33E-01 | 9.28E-01 | 1.64E-01 | 5.24E+00 | 3.39E-21 | 0.0088 | 0.7818 |
| Neuroticism \|\| id:ebi-a-GCST005232 | Inverse variance weighted | 66 | 1.81E-01 | 1.18E+00 | 9.24E-01 | 1.52E+00 | 5.89E-21 | | |
| Neuroticism score \|\| id:ukb-b-4630 | MR Egger | 107 | 1.56E-01 | 1.60E+00 | 8.40E-01 | 3.05E+00 | 8.19E-25 | -0.0198 | 0.2571 |
| Neuroticism score \|\| id:ukb-b-4630 | Inverse variance weighted | 107 | 1.04E-01 | 1.11E+00 | 9.79E-01 | 1.25E+00 | 3.61E-25 | | |
| Number of operations, self-reported \|\| id:ukb-b-4733 | MR Egger | 21 | 5.53E-01 | 4.69E-01 | 4.02E-02 | 5.48E+00 | 4.07E-01 | 0.0111 | 0.4565 |
| Number of operations, self-reported \|\| id:ukb-b-4733 | Inverse variance weighted | 21 | 6.79E-01 | 1.17E+00 | 5.65E-01 | 2.40E+00 | 4.34E-01 | | |
| Other serious medical condition/disability diagnosed by doctor \|\| id:ukb-b-14961 | MR Egger | 7 | 6.39E-01 | 1.39E-01 | 5.89E-05 | 3.26E+02 | 1.50E-01 | 0.0131 | 0.6687 |
| Other serious medical condition/disability diagnosed by doctor \|\| id:ukb-b-14961 | Inverse variance weighted | 7 | 8.24E-01 | 7.36E-01 | 4.96E-02 | 1.09E+01 | 2.07E-01 | | |
| Parkinson's disease \|\| id:ieu-b-7 | MR Egger | 20 | 1.78E-01 | 8.61E-01 | 6.98E-01 | 1.06E+00 | 3.04E-01 | 0.0119 | 0.4669 |
| Parkinson's disease \|\| id:ieu-b-7 | Inverse variance weighted | 20 | 2.45E-02 | 9.29E-01 | 8.70E-01 | 9.90E-01 | 3.28E-01 | | |
| Pulse wave Arterial Stiffness index \|\| id:ukb-b-11971 | MR Egger | 3 | 2.23E-01 | 4.87E-08 | 2.87E-13 | 8.25E-03 | 7.48E-01 | 0.5146 | 0.2123 |
| Pulse wave Arterial Stiffness index \|\| id:ukb-b-11971 | Inverse variance weighted | 3 | 2.99E-01 | 2.36E+00 | 4.67E-01 | 1.19E+01 | 1.47E-02 | | |
| Pulse wave peak to peak time \|\| id:ukb-b-8778 | MR Egger | 3 | 5.72E-01 | 1.20E+06 | 1.33E-09 | 1.08E+21 | 3.09E-02 | -0.4404 | 0.5523 |
| Pulse wave peak to peak time \|\| id:ukb-b-8778 | Inverse variance weighted | 3 | 2.64E-01 | 4.14E-01 | 8.81E-02 | 1.95E+00 | 1.82E-02 | | |
| Qualifications: A levels/AS levels or equivalent \|\| id:ukb-b-11615 | MR Egger | 81 | 6.87E-01 | 2.40E+00 | 3.47E-02 | 1.65E+02 | 2.01E-02 | -0.0087 | 0.5299 |
| Qualifications: A levels/AS levels or equivalent \|\| id:ukb-b-11615 | Inverse variance weighted | 81 | 2.29E-01 | 6.27E-01 | 2.93E-01 | 1.34E+00 | 2.22E-02 | | |
| Qualifications: College or University degree \|\| id:ukb-b-16489 | MR Egger | 228 | 8.12E-01 | 8.32E-01 | 1.82E-01 | 3.80E+00 | 9.88E-03 | -0.0048 | 0.4016 |

| exposure | method | nsnp | pval | or | or lci95 | or uci95 | Q pval | egger intercept | ple pval |
|---|---|---|---|---|---|---|---|---|---|
| Qualifications: College or University degree \|\| id:ukb-b-16489 | Inverse variance weighted | 228 | 2.68E-05 | 4.43E-01 | 3.03E-01 | 6.48E-01 | 1.01E-02 | | |
| Qualifications: CSEs or equivalent \|\| id:ukb-b-10505 | MR Egger | 6 | 8.16E-01 | 3.99E+00 | 7.41E-05 | 2.15E+05 | 9.92E-01 | -0.0017 | 0.9579 |
| Qualifications: CSEs or equivalent \|\| id:ukb-b-10505 | Inverse variance weighted | 6 | 4.81E-01 | 2.96E+00 | 1.45E-01 | 6.05E+01 | 9.98E-01 | | |
| Qualifications: None of the above \|\| id:ukb-b-17729 | MR Egger | 86 | 1.73E-01 | 1.71E+01 | 2.99E-01 | 9.82E+02 | 1.43E-01 | -0.0090 | 0.4471 |
| Qualifications: None of the above \|\| id:ukb-b-17729 | Inverse variance weighted | 86 | 1.09E-03 | 3.64E+00 | 1.68E+00 | 7.91E+00 | 1.49E-01 | | |
| Qualifications: O levels/GCSEs or equivalent \|\| id:ukb-b-18099 | MR Egger | 19 | 8.89E-01 | 2.49E-01 | 1.12E-09 | 5.53E+07 | 2.78E-08 | -0.0034 | 0.9607 |
| Qualifications: O levels/GCSEs or equivalent \|\| id:ukb-b-18099 | Inverse variance weighted | 19 | 1.36E-01 | 1.53E-01 | 1.30E-02 | 1.81E+00 | 5.78E-08 | | |
| Qualifications: Other professional qualifications eg: nursing, teaching \|\| id:ukb-b-13799 | MR Egger | 17 | 6.54E-01 | 1.54E-03 | 1.40E-15 | 1.69E+09 | 3.73E-27 | 0.0205 | 0.8266 |
| Qualifications: Other professional qualifications eg: nursing, teaching \|\| id:ukb-b-13799 | Inverse variance weighted | 17 | 1.49E-01 | 3.44E-02 | 3.54E-04 | 3.34E+00 | 9.83E-27 | | |
| Reason for glasses/contact lenses: For short-sightedness, i.e. only or mainly for distance viewing such as driving, cinema etc (called 'myopia') \|\| id:ukb-b-6353 | MR Egger | 26 | 8.28E-01 | 4.93E-01 | 8.79E-04 | 2.76E+02 | 9.99E-03 | 0.0048 | 0.7623 |
| Reason for glasses/contact lenses: For short-sightedness, i.e. only or mainly for distance viewing such as driving, cinema etc (called 'myopia') \|\| id:ukb-b-6353 | Inverse variance weighted | 26 | 8.33E-01 | 1.25E+00 | 1.56E-01 | 1.00E+01 | 1.35E-02 | | |
| Seen a psychiatrist for nerves, anxiety, tension or depression \|\| id:ukb-b-18336 | MR Egger | 4 | 1.62E-01 | 7.86E+09 | 8.92E+00 | 6.92E+18 | 8.57E-01 | -0.0784 | 0.2413 |
| Seen a psychiatrist for nerves, anxiety, tension or depression \|\| id:ukb-b-18336 | Inverse variance weighted | 4 | 6.18E-03 | 3.41E+02 | 5.25E+00 | 2.22E+04 | 3.88E-01 | | |
| Seen doctor (GP) for nerves, anxiety, tension or depression \|\| id:ukb-b-6991 | MR Egger | 37 | 4.87E-01 | 7.96E+00 | 2.42E-02 | 2.61E+03 | 7.58E-01 | -0.0095 | 0.6267 |

| exposure | method | nsnp | pval | or | or lci95 | or uci95 | Q pval | egger intercept | ple pval |
|---|---|---|---|---|---|---|---|---|---|
| Seen doctor (GP) for nerves, anxiety, tension or depression \|\| id:ukb-b-6991 | Inverse variance weighted | 37 | 1.87E-01 | 1.90E+00 | 7.31E-01 | 4.95E+00 | 7.85E-01 | | |
| Sensitivity / hurt feelings \|\| id:ukb-b-9981 | MR Egger | 36 | 6.24E-01 | 2.77E-01 | 1.73E-03 | 4.44E+01 | 8.79E-02 | 0.0155 | 0.4091 |
| Sensitivity / hurt feelings \|\| id:ukb-b-9981 | Inverse variance weighted | 36 | 1.07E-01 | 2.31E+00 | 8.35E-01 | 6.40E+00 | 9.15E-02 | | |
| Sleeplessness / insomnia \|\| id:ukb-b-3957 | MR Egger | 38 | 7.84E-01 | 1.29E+00 | 2.15E-01 | 7.72E+00 | 1.22E-01 | 0.0076 | 0.4734 |
| Sleeplessness / insomnia \|\| id:ukb-b-3957 | Inverse variance weighted | 38 | 4.35E-03 | 2.40E+00 | 1.31E+00 | 4.38E+00 | 1.32E-01 | | |
| Sodium in urine \|\| id:ukb-a-335 | MR Egger | 29 | 4.09E-01 | 3.37E-01 | 2.63E-02 | 4.30E+00 | 5.71E-03 | 0.0293 | 0.2131 |
| Sodium in urine \|\| id:ukb-a-335 | Inverse variance weighted | 29 | 6.81E-02 | 1.69E+00 | 9.62E-01 | 2.99E+00 | 3.74E-03 | | |
| Suffer from 'nerves' \|\| id:ukb-b-19957 | MR Egger | 21 | 2.89E-01 | 2.50E+02 | 1.24E-02 | 5.02E+06 | 9.52E-02 | -0.0352 | 0.2546 |
| Suffer from 'nerves' \|\| id:ukb-b-19957 | Inverse variance weighted | 21 | 7.00E-01 | 7.16E-01 | 1.31E-01 | 3.91E+00 | 7.99E-02 | | |
| triglycerides \|\| id:ieu-b-111 | MR Egger | 261 | 1.02E-01 | 1.14E+00 | 9.75E-01 | 1.33E+00 | 3.38E-11 | 0.0013 | 0.6046 |
| triglycerides \|\| id:ieu-b-111 | Inverse variance weighted | 261 | 3.17E-03 | 1.17E+00 | 1.06E+00 | 1.31E+00 | 4.01E-11 | | |
| Type 2 diabetes \|\| id:ebi-a-GCST006867 | MR Egger | 114 | 9.10E-01 | 9.89E-01 | 8.15E-01 | 1.20E+00 | 2.20E-17 | 0.0034 | 0.6312 |
| Type 2 diabetes \|\| id:ebi-a-GCST006867 | Inverse variance weighted | 114 | 4.39E-01 | 1.03E+00 | 9.52E-01 | 1.12E+00 | 2.96E-17 | | |
| Vascular/heart problems diagnosed by doctor: Angina \|\| id:ukb-b-8468 | MR Egger | 21 | 2.71E-02 | 1.62E+04 | 5.80E+00 | 4.52E+07 | 4.21E-01 | -0.0287 | 0.0355 |
| Vascular/heart problems diagnosed by doctor: Angina \|\| id:ukb-b-8468 | Inverse variance weighted | 21 | 4.74E-01 | 3.16E+00 | 1.35E-01 | 7.38E+01 | 2.08E-01 | | |
| Vascular/heart problems diagnosed by doctor: High blood pressure \|\| id:ukb-b-14177 | MR Egger | 207 | 4.19E-01 | 6.24E-01 | 1.99E-01 | 1.96E+00 | 8.98E-08 | 0.0059 | 0.2335 |
| Vascular/heart problems diagnosed by doctor: High blood pressure \|\| id:ukb-b-14177 | Inverse variance weighted | 207 | 4.00E-01 | 1.19E+00 | 7.90E-01 | 1.81E+00 | 7.34E-08 | | |
| Vascular/heart problems diagnosed by doctor: None of the above \|\| id:ukb-b-13352 | MR Egger | 199 | 7.48E-01 | 1.22E+00 | 3.68E-01 | 4.03E+00 | 1.01E-09 | -0.0032 | 0.5445 |

| exposure | method | nsnp | pval | or | or lci95 | or uci95 | Q pval | egger intercept | ple pval |
|---|---|---|---|---|---|---|---|---|---|
| Vascular/heart problems diagnosed by doctor: None of the above \|\| id:ukb-b-13352 | Inverse variance weighted | 199 | 5.00E-01 | 8.61E-01 | 5.58E-01 | 1.33E+00 | 1.17E-09 | | |
| Waist-to-hip ratio \|\| id:ieu-a-72 | MR Egger | 29 | 9.26E-01 | 9.16E-01 | 1.46E-01 | 5.75E+00 | 7.18E-04 | 0.0061 | 0.7942 |
| Waist-to-hip ratio \|\| id:ieu-a-72 | Inverse variance weighted | 29 | 4.52E-01 | 1.17E+00 | 7.82E-01 | 1.74E+00 | 1.04E-03 | | |
| Wheeze or whistling in the chest in last year \|\| id:ukb-b-18335 | MR Egger | 44 | 1.61E-01 | 9.18E-03 | 1.45E-05 | 5.81E+00 | 2.75E-10 | 0.0358 | 0.1168 |
| Wheeze or whistling in the chest in last year \|\| id:ukb-b-18335 | Inverse variance weighted | 44 | 5.92E-01 | 1.53E+00 | 3.21E-01 | 7.34E+00 | 3.42E-11 | | |
| Whole body fat mass \|\| id:ukb-b-19393 | MR Egger | 387 | 9.49E-02 | 1.36E+00 | 9.49E-01 | 1.95E+00 | 2.03E-06 | 0.0005 | 0.8832 |
| Whole body fat mass \|\| id:ukb-b-19393 | Inverse variance weighted | 387 | 4.41E-07 | 1.40E+00 | 1.23E+00 | 1.59E+00 | 2.38E-06 | | |
| Whole body fat-free mass \|\| id:ukb-b-13354 | MR Egger | 486 | 2.50E-01 | 8.03E-01 | 5.54E-01 | 1.17E+00 | 5.34E-10 | 0.0044 | 0.0888 |
| Whole body fat-free mass \|\| id:ukb-b-13354 | Inverse variance weighted | 486 | 3.69E-01 | 1.08E+00 | 9.17E-01 | 1.26E+00 | 3.32E-10 | | |
| Years of schooling \|\| id:ieu-a-1239 | MR Egger | 287 | 2.48E-01 | 6.67E-01 | 3.36E-01 | 1.32E+00 | 1.12E-01 | -0.0022 | 0.6536 |
| Years of schooling \|\| id:ieu-a-1239 | Inverse variance weighted | 287 | 6.12E-10 | 5.73E-01 | 4.80E-01 | 6.84E-01 | 1.18E-01 | | |

**Table S6. Mendelian randomization results (psoriasis -> outcome).**
Columns: nsnp, number of valid SNPs used for analysis; pval, p-value; b, beta estimate; lo ci; lower 95 % confidence interval; up ci, upper 95 % confidence interval; Q pval, Q-test p-value (for testing heterogeneity); egger intercept, MR Egger intercept; and ple pval, pleiotropy p-value.

| outcome | method | nsnp | pval | b | lo ci | up ci | Q pval | egger intercept | ple pval |
|---|---|---|---|---|---|---|---|---|---|
| Age completed full time education \|\| id:ukb-b-6134 | MR Egger | 21 | 2.15E-02 | -1.77E-02 | -3.15E-02 | -3.85E-03 | 2.32E-02 | 1.46E-03 | 0.3063 |
| Age completed full time education \|\| id:ukb-b-6134 | Inverse variance weighted | 21 | 3.38E-03 | -1.15E-02 | -1.92E-02 | -3.81E-03 | 1.98E-02 | | |
| Alcohol intake frequency. \|\| id:ukb-b-5779 | MR Egger | 21 | 1.01E-01 | 2.40E-02 | -3.26E-03 | 5.13E-02 | 4.05E-06 | -3.43E-03 | 0.2263 |
| Alcohol intake frequency. \|\| id:ukb-b-5779 | Inverse variance weighted | 21 | 2.23E-01 | 9.53E-03 | -5.80E-03 | 2.49E-02 | 1.26E-06 | | |
| apolipoprotein A-I \|\| id:ieu-b-107 | MR Egger | 21 | 5.46E-02 | 2.69E-02 | 1.16E-03 | 5.27E-02 | 4.91E-18 | -1.83E-03 | 0.4887 |
| apolipoprotein A-I \|\| id:ieu-b-107 | Inverse variance weighted | 21 | 7.64E-03 | 1.92E-02 | 5.09E-03 | 3.33E-02 | 3.08E-18 | | |
| Blood clot, DVT, bronchitis, emphysema, asthma, rhinitis, eczema, allergy diagnosed by doctor: Blood clot in the leg (DVT) \|\| id:ukb-b-11411 | MR Egger | 20 | 1.54E-02 | -2.06E-03 | -3.57E-03 | -5.52E-04 | 8.89E-01 | 3.25E-04 | 0.0449 |
| Blood clot, DVT, bronchitis, emphysema, asthma, rhinitis, eczema, allergy diagnosed by doctor: Blood clot in the leg (DVT) \|\| id:ukb-b-11411 | Inverse variance weighted | 20 | 1.12E-01 | -6.81E-04 | -1.52E-03 | 1.59E-04 | 6.72E-01 | | |
| Blood clot, DVT, bronchitis, emphysema, asthma, rhinitis, eczema, allergy diagnosed by doctor: Blood clot in the lung \|\| id:ukb-b-13704 | MR Egger | 15 | 6.63E-01 | -6.05E-04 | -3.26E-03 | 2.05E-03 | 6.77E-01 | 1.01E-04 | 0.6207 |
| Blood clot, DVT, bronchitis, emphysema, asthma, rhinitis, eczema, | Inverse variance weighted | 15 | 8.86E-01 | 5.48E-05 | -6.97E-04 | 8.06E-04 | 7.28E-01 | | |

| outcome | method | nsnp | pval | b | lo ci | up ci | Q pval | egger intercept | ple pval |
|---|---|---|---|---|---|---|---|---|---|
| allergy diagnosed by doctor: Blood clot in the lung \|\| id:ukb-b-13704 | | | | | | | | | |
| Blood clot, DVT, bronchitis, emphysema, asthma, rhinitis, eczema, allergy diagnosed by doctor: Emphysema/chronic bronchitis \|\| id:ukb-b-16207 | MR Egger | 19 | 7.71E-01 | -2.99E-04 | -2.28E-03 | 1.68E-03 | 7.53E-03 | 4.46E-05 | 0.8225 |
| Blood clot, DVT, bronchitis, emphysema, asthma, rhinitis, eczema, allergy diagnosed by doctor: Emphysema/chronic bronchitis \|\| id:ukb-b-16207 | Inverse variance weighted | 19 | 8.44E-01 | -1.08E-04 | -1.19E-03 | 9.70E-04 | 1.10E-02 | | |
| Body mass index (BMI) \|\| id:ukb-b-19953 | MR Egger | 21 | 3.25E-01 | 9.21E-03 | -8.66E-03 | 2.71E-02 | 3.28E-06 | -2.11E-03 | 0.2546 |
| Body mass index (BMI) \|\| id:ukb-b-19953 | Inverse variance weighted | 21 | 9.53E-01 | 3.03E-04 | -9.71E-03 | 1.03E-02 | 1.24E-06 | | |
| Celiac disease \|\| id:ieu-a-1058 | MR Egger | 7 | 2.53E-01 | -1.79E+00 | -4.52E+00 | 9.31E-01 | 3.14E-66 | 2.99E-01 | 0.2214 |
| Celiac disease \|\| id:ieu-a-1058 | Inverse variance weighted | 7 | 9.28E-01 | 4.36E-02 | -9.03E-01 | 9.90E-01 | 8.20E-92 | | |
| College completion \|\| id:ieu-a-836 | MR Egger | 12 | 9.04E-02 | -1.55E-01 | -3.18E-01 | 7.13E-03 | 2.69E-01 | 2.04E-02 | 0.1229 |
| College completion \|\| id:ieu-a-836 | Inverse variance weighted | 12 | 4.09E-01 | -2.19E-02 | -7.40E-02 | 3.01E-02 | 1.52E-01 | | |
| Coronary artery disease \|\| id:ebi-a-GCST005195 | MR Egger | 18 | 9.54E-01 | 1.73E-03 | -5.60E-02 | 5.95E-02 | 5.76E-04 | -3.26E-03 | 0.5928 |
| Coronary artery disease \|\| id:ebi-a-GCST005195 | Inverse variance weighted | 18 | 4.93E-01 | -1.14E-02 | -4.40E-02 | 2.12E-02 | 7.52E-04 | | |
| Creatinine (enzymatic) in urine \|\| id:ukb-a-333 | MR Egger | 20 | 2.93E-03 | 2.19E-02 | 9.43E-03 | 3.44E-02 | 3.21E-01 | -1.93E-03 | 0.1389 |
| Creatinine (enzymatic) in urine \|\| id:ukb-a-333 | Inverse variance weighted | 20 | 2.42E-04 | 1.38E-02 | 6.44E-03 | 2.12E-02 | 2.42E-01 | | |
| Current tobacco smoking \|\| id:ukb-b-223 | MR Egger | 21 | 1.56E-01 | 5.22E-03 | -1.70E-03 | 1.21E-02 | 9.83E-02 | -5.51E-04 | 0.4384 |
| Current tobacco smoking \|\| id:ukb-b-223 | Inverse variance weighted | 21 | 1.36E-01 | 2.89E-03 | -9.10E-04 | 6.69E-03 | 1.05E-01 | | |

| outcome | method | nsnp | pval | b | lo ci | up ci | Q pval | egger intercept | ple pval |
|---|---|---|---|---|---|---|---|---|---|
| Depressive symptoms \|\| id:ieu-a-1000 | MR Egger | 17 | 6.68E-01 | 1.27E-02 | -4.42E-02 | 6.95E-02 | 1.92E-01 | -1.62E-03 | 0.7030 |
| Depressive symptoms \|\| id:ieu-a-1000 | Inverse variance weighted | 17 | 8.24E-01 | 1.96E-03 | -1.53E-02 | 1.93E-02 | 2.34E-01 | | |
| Diabetes diagnosed by doctor \|\| id:ukb-b-10753 | MR Egger | 21 | 8.87E-01 | 4.48E-04 | -5.63E-03 | 6.52E-03 | 4.41E-20 | 1.93E-04 | 0.7559 |
| Diabetes diagnosed by doctor \|\| id:ukb-b-10753 | Inverse variance weighted | 21 | 4.53E-01 | 1.26E-03 | -2.03E-03 | 4.56E-03 | 8.85E-20 | | |
| Diagnoses - main ICD10: G56 Mononeuropathies of upper limb \|\| id:ukb-a-528 | MR Egger | 20 | 8.69E-01 | 1.39E-04 | -1.50E-03 | 1.77E-03 | 7.40E-01 | -9.12E-05 | 0.5826 |
| Diagnoses - main ICD10: G56 Mononeuropathies of upper limb \|\| id:ukb-a-528 | Inverse variance weighted | 20 | 6.07E-01 | -2.44E-04 | -1.18E-03 | 6.87E-04 | 7.75E-01 | | |
| Diagnoses - main ICD10: I20 Angina pectoris \|\| id:ukb-a-532 | MR Egger | 20 | 6.71E-01 | -3.27E-04 | -1.81E-03 | 1.16E-03 | 6.65E-01 | -6.72E-05 | 0.6550 |
| Diagnoses - main ICD10: I20 Angina pectoris \|\| id:ukb-a-532 | Inverse variance weighted | 20 | 1.57E-01 | -6.10E-04 | -1.45E-03 | 2.36E-04 | 7.12E-01 | | |
| Diagnoses - main ICD10: I25 Chronic ischaemic heart disease \|\| id:ukb-a-534 | MR Egger | 20 | 5.40E-01 | -7.27E-04 | -3.01E-03 | 1.55E-03 | 1.55E-01 | 6.15E-05 | 0.7899 |
| Diagnoses - main ICD10: I25 Chronic ischaemic heart disease \|\| id:ukb-a-534 | Inverse variance weighted | 20 | 4.68E-01 | -4.69E-04 | -1.74E-03 | 7.98E-04 | 1.92E-01 | | |
| Diagnoses - main ICD10: J44 Other chronic obstructive pulmonary disease \|\| id:ukb-a-543 | MR Egger | 20 | 6.52E-01 | -1.50E-04 | -7.94E-04 | 4.93E-04 | 4.25E-01 | -4.48E-06 | 0.9451 |
| Diagnoses - main ICD10: J44 Other chronic obstructive pulmonary disease \|\| id:ukb-a-543 | Inverse variance weighted | 20 | 3.59E-01 | -1.69E-04 | -5.31E-04 | 1.93E-04 | 4.91E-01 | | |
| Diagnoses - main ICD10: K20 Oesophagitis \|\| id:ukb-b-19354 | MR Egger | 17 | 8.79E-01 | -8.86E-05 | -1.21E-03 | 1.03E-03 | 7.02E-01 | -9.25E-06 | 0.9338 |
| Diagnoses - main ICD10: K20 Oesophagitis \|\| id:ukb-b-19354 | Inverse variance weighted | 17 | 6.92E-01 | -1.28E-04 | -7.63E-04 | 5.06E-04 | 7.64E-01 | | |

| outcome | method | nsnp | pval | b | lo ci | up ci | Q pval | egger intercept | ple pval |
|---|---|---|---|---|---|---|---|---|---|
| Diagnoses - main ICD10: K21 Gastro-oesophageal reflux disease \|\| id:ukb-a-545 | MR Egger | 20 | 4.61E-01 | -6.98E-04 | -2.51E-03 | 1.12E-03 | 6.20E-01 | 2.23E-04 | 0.2330 |
| Diagnoses - main ICD10: K21 Gastro-oesophageal reflux disease \|\| id:ukb-a-545 | Inverse variance weighted | 20 | 6.47E-01 | 2.41E-04 | -7.93E-04 | 1.28E-03 | 5.81E-01 | | |
| Diagnoses - main ICD10: K52 Other non-infective gastro-enteritis and colitis \|\| id:ukb-a-554 | MR Egger | 20 | 8.82E-01 | 1.32E-04 | -1.58E-03 | 1.84E-03 | 6.01E-01 | 8.11E-06 | 0.9626 |
| Diagnoses - main ICD10: K52 Other non-infective gastro-enteritis and colitis \|\| id:ukb-a-554 | Inverse variance weighted | 20 | 7.39E-01 | 1.66E-04 | -8.09E-04 | 1.14E-03 | 6.65E-01 | | |
| Diagnoses - main ICD10: K57 Diverticular disease of intestine \|\| id:ukb-a-555 | MR Egger | 20 | 2.53E-01 | 1.39E-03 | -9.18E-04 | 3.70E-03 | 2.96E-02 | 1.57E-04 | 0.5046 |
| Diagnoses - main ICD10: K57 Diverticular disease of intestine \|\| id:ukb-a-555 | Inverse variance weighted | 20 | 1.94E-03 | 2.05E-03 | 7.54E-04 | 3.35E-03 | 3.39E-02 | | |
| Diagnoses - main ICD10: K80 Cholelithiasis \|\| id:ukb-a-559 | MR Egger | 20 | 5.23E-02 | 1.93E-03 | 1.09E-04 | 3.75E-03 | 3.97E-01 | -1.17E-04 | 0.5285 |
| Diagnoses - main ICD10: K80 Cholelithiasis \|\| id:ukb-a-559 | Inverse variance weighted | 20 | 5.73E-03 | 1.44E-03 | 4.18E-04 | 2.46E-03 | 4.35E-01 | | |
| Diagnoses - main ICD10: M25 Other joint disorders, not elsewhere classified \|\| id:ukb-d-M25 | MR Egger | 20 | 9.12E-01 | -1.09E-04 | -2.03E-03 | 1.81E-03 | 2.30E-01 | -1.14E-04 | 0.5567 |
| Diagnoses - main ICD10: M25 Other joint disorders, not elsewhere classified \|\| id:ukb-d-M25 | Inverse variance weighted | 20 | 2.81E-01 | -5.90E-04 | -1.66E-03 | 4.83E-04 | 2.61E-01 | | |
| Diagnoses - main ICD10: R07 Pain in throat and chest \|\| id:ukb-a-581 | MR Egger | 20 | 5.36E-01 | 8.68E-04 | -1.83E-03 | 3.56E-03 | 4.88E-01 | -2.83E-04 | 0.3070 |
| Diagnoses - main ICD10: R07 Pain in throat and chest \|\| id:ukb-a-581 | Inverse variance weighted | 20 | 6.82E-01 | -3.21E-04 | -1.86E-03 | 1.21E-03 | 4.82E-01 | | |

| outcome | method | nsnp | pval | b | lo ci | up ci | Q pval | egger intercept | ple pval |
|---|---|---|---|---|---|---|---|---|---|
| Diagnoses - main ICD10: R10 Abdominal and pelvic pain \|\| id:ukb-a-582 | MR Egger | 20 | 1.51E-02 | 3.92E-03 | 1.06E-03 | 6.78E-03 | 1.72E-01 | -3.72E-04 | 0.2085 |
| Diagnoses - main ICD10: R10 Abdominal and pelvic pain \|\| id:ukb-a-582 | Inverse variance weighted | 20 | 5.41E-03 | 2.36E-03 | 6.96E-04 | 4.02E-03 | 1.38E-01 | | |
| Diagnoses - secondary ICD10: M06.99 Rheumatoid arthritis, unspecified (Site unspecified) \|\| id:ukb-b-11874 | MR Egger | 10 | 6.94E-02 | 6.07E-03 | 3.93E-04 | 1.17E-02 | 1.63E-14 | -7.57E-04 | 0.1218 |
| Diagnoses - secondary ICD10: M06.99 Rheumatoid arthritis, unspecified (Site unspecified) \|\| id:ukb-b-11874 | Inverse variance weighted | 10 | 1.79E-01 | 1.29E-03 | -5.91E-04 | 3.17E-03 | 3.29E-20 | | |
| ECG, heart rate \|\| id:ukb-b-11361 | MR Egger | 21 | 3.46E-01 | 1.43E-02 | -1.47E-02 | 4.32E-02 | 2.81E-01 | -1.18E-03 | 0.6894 |
| ECG, heart rate \|\| id:ukb-b-11361 | Inverse variance weighted | 21 | 2.47E-01 | 9.28E-03 | -6.45E-03 | 2.50E-02 | 3.27E-01 | | |
| Ever had bowel cancer screening \|\| id:ukb-b-17001 | MR Egger | 21 | 2.82E-01 | 3.51E-03 | -2.71E-03 | 9.73E-03 | 5.44E-02 | -1.23E-04 | 0.8463 |
| Ever had bowel cancer screening \|\| id:ukb-b-17001 | Inverse variance weighted | 21 | 8.13E-02 | 2.99E-03 | -3.72E-04 | 6.36E-03 | 7.22E-02 | | |
| Eye problems/disorders: Diabetes related eye disease \|\| id:ukb-b-10181 | MR Egger | 14 | 1.02E-01 | 1.49E-02 | -1.59E-03 | 3.15E-02 | 3.65E-09 | -1.76E-03 | 0.1783 |
| Eye problems/disorders: Diabetes related eye disease \|\| id:ukb-b-10181 | Inverse variance weighted | 14 | 1.78E-01 | 3.39E-03 | -1.54E-03 | 8.31E-03 | 8.46E-11 | | |
| Fed-up feelings \|\| id:ukb-b-19809 | MR Egger | 21 | 3.76E-03 | 9.69E-03 | 3.93E-03 | 1.54E-02 | 2.29E-01 | -1.10E-03 | 0.0720 |
| Fed-up feelings \|\| id:ukb-b-19809 | Inverse variance weighted | 21 | 3.65E-03 | 5.04E-03 | 1.64E-03 | 8.43E-03 | 1.19E-01 | | |
| Fluid intelligence score \|\| id:ukb-b-5238 | MR Egger | 21 | 2.10E-01 | -4.35E-02 | -1.09E-01 | 2.23E-02 | 1.70E-05 | 1.74E-03 | 0.7951 |
| Fluid intelligence score \|\| id:ukb-b-5238 | Inverse variance weighted | 21 | 4.68E-02 | -3.62E-02 | -7.18E-02 | -5.11E-04 | 2.81E-05 | | |
| Forced expiratory volume in 1-second (FEV1) \|\| id:ukb-b-19657 | MR Egger | 21 | 9.54E-01 | -6.97E-04 | -2.40E-02 | 2.26E-02 | 3.14E-19 | 6.13E-04 | 0.7968 |
| Forced expiratory volume in 1-second (FEV1) \|\| id:ukb-b-19657 | Inverse variance weighted | 21 | 7.70E-01 | 1.89E-03 | -1.07E-02 | 1.45E-02 | 6.90E-19 | | |

| outcome | method | nsnp | pval | b | lo ci | up ci | Q pval | egger intercept | ple pval |
|---|---|---|---|---|---|---|---|---|---|
| Forced expiratory volume in 1-second (FEV1), Best measure \|\| id:ukb-b-11141 | MR Egger | 21 | 9.92E-01 | 1.29E-04 | -2.38E-02 | 2.41E-02 | 3.93E-16 | 6.21E-04 | 0.7993 |
| Forced expiratory volume in 1-second (FEV1), Best measure \|\| id:ukb-b-11141 | Inverse variance weighted | 21 | 6.78E-01 | 2.75E-03 | -1.02E-02 | 1.57E-02 | 8.40E-16 | | |
| Forced expiratory volume in 1-second (FEV1), predicted percentage \|\| id:ukb-b-13405 | MR Egger | 21 | 6.06E-01 | 8.08E-03 | -2.21E-02 | 3.83E-02 | 2.83E-05 | -2.03E-04 | 0.9473 |
| Forced expiratory volume in 1-second (FEV1), predicted percentage \|\| id:ukb-b-13405 | Inverse variance weighted | 21 | 3.86E-01 | 7.23E-03 | -9.11E-03 | 2.36E-02 | 4.96E-05 | | |
| Forced vital capacity (FVC) \|\| id:ukb-b-7953 | MR Egger | 21 | 5.40E-01 | -7.18E-03 | -2.97E-02 | 1.54E-02 | 2.41E-20 | 1.00E-03 | 0.6628 |
| Forced vital capacity (FVC) \|\| id:ukb-b-7953 | Inverse variance weighted | 21 | 6.38E-01 | -2.94E-03 | -1.52E-02 | 9.31E-03 | 3.55E-20 | | |
| Forced vital capacity (FVC), Best measure \|\| id:ukb-b-14713 | MR Egger | 21 | 7.11E-01 | -4.46E-03 | -2.77E-02 | 1.88E-02 | 2.38E-17 | 7.89E-04 | 0.7395 |
| Forced vital capacity (FVC), Best measure \|\| id:ukb-b-14713 | Inverse variance weighted | 21 | 8.60E-01 | -1.13E-03 | -1.37E-02 | 1.15E-02 | 4.52E-17 | | |
| Frequency of depressed mood in last 2 weeks \|\| id:ukb-b-3822 | MR Egger | 21 | 1.86E-03 | 1.26E-02 | 5.75E-03 | 1.94E-02 | 3.67E-01 | -1.14E-03 | 0.1123 |
| Frequency of depressed mood in last 2 weeks \|\| id:ukb-b-3822 | Inverse variance weighted | 21 | 1.19E-04 | 7.76E-03 | 3.80E-03 | 1.17E-02 | 2.67E-01 | | |
| Frequency of tenseness / restlessness in last 2 weeks \|\| id:ukb-b-5664 | MR Egger | 21 | 7.06E-03 | 1.45E-02 | 5.07E-03 | 2.38E-02 | 5.18E-03 | -1.94E-03 | 0.0539 |
| Frequency of tenseness / restlessness in last 2 weeks \|\| id:ukb-b-5664 | Inverse variance weighted | 21 | 2.85E-02 | 6.27E-03 | 6.58E-04 | 1.19E-02 | 5.84E-04 | | |
| Frequency of tiredness / lethargy in last 2 weeks \|\| id:ukb-b-929 | MR Egger | 21 | 9.94E-02 | 8.91E-03 | -1.17E-03 | 1.90E-02 | 1.71E-01 | -6.44E-04 | 0.5330 |
| Frequency of tiredness / lethargy in last 2 weeks \|\| id:ukb-b-929 | Inverse variance weighted | 21 | 2.76E-02 | 6.20E-03 | 6.85E-04 | 1.17E-02 | 1.94E-01 | | |

| outcome | method | nsnp | pval | b | lo ci | up ci | Q pval | egger intercept | ple pval |
|---|---|---|---|---|---|---|---|---|---|
| Frequency of unenthusiasm / disinterest in last 2 weeks \|\| id:ukb-b-1419 | MR Egger | 21 | 4.61E-03 | 1.43E-02 | 5.58E-03 | 2.31E-02 | 1.41E-02 | -1.90E-03 | 0.0442 |
| Frequency of unenthusiasm / disinterest in last 2 weeks \|\| id:ukb-b-1419 | Inverse variance weighted | 21 | 1.88E-02 | 6.33E-03 | 1.05E-03 | 1.16E-02 | 1.76E-03 | | |
| Had other major operations \|\| id:ukb-b-15378 | MR Egger | 21 | 6.10E-01 | 1.78E-03 | -4.96E-03 | 8.53E-03 | 8.48E-01 | -6.22E-04 | 0.3712 |
| Had other major operations \|\| id:ukb-b-15378 | Inverse variance weighted | 21 | 6.61E-01 | -8.38E-04 | -4.58E-03 | 2.91E-03 | 8.47E-01 | | |
| HDL cholesterol \|\| id:ieu-b-109 | MR Egger | 21 | 9.20E-02 | 2.46E-02 | -2.57E-03 | 5.17E-02 | 2.56E-22 | -2.41E-03 | 0.3871 |
| HDL cholesterol \|\| id:ieu-b-109 | Inverse variance weighted | 21 | 5.99E-02 | 1.44E-02 | -5.97E-04 | 2.93E-02 | 4.79E-23 | | |
| Hearing difficulty/problems with background noise \|\| id:ukb-b-18275 | MR Egger | 21 | 7.84E-01 | -1.12E-03 | -9.07E-03 | 6.82E-03 | 5.43E-04 | 7.97E-04 | 0.3308 |
| Hearing difficulty/problems with background noise \|\| id:ukb-b-18275 | Inverse variance weighted | 21 | 3.19E-01 | 2.24E-03 | -2.16E-03 | 6.65E-03 | 4.10E-04 | | |
| Hearing difficulty/problems: Yes \|\| id:ukb-a-257 | MR Egger | 20 | 9.64E-01 | 1.93E-04 | -8.02E-03 | 8.41E-03 | 2.69E-03 | 5.55E-05 | 0.9468 |
| Hearing difficulty/problems: Yes \|\| id:ukb-a-257 | Inverse variance weighted | 20 | 8.54E-01 | 4.27E-04 | -4.13E-03 | 4.98E-03 | 4.16E-03 | | |
| HOMA-B \|\| id:ieu-b-117 | MR Egger | 11 | 5.97E-01 | -1.71E-02 | -7.84E-02 | 4.41E-02 | 9.53E-01 | 3.42E-03 | 0.4481 |
| HOMA-B \|\| id:ieu-b-117 | Inverse variance weighted | 11 | 4.56E-01 | 6.63E-03 | -1.08E-02 | 2.40E-02 | 9.52E-01 | | |
| HOMA-IR \|\| id:ieu-b-118 | MR Egger | 11 | 1.31E-01 | -5.12E-02 | -1.12E-01 | 9.16E-03 | 7.69E-01 | 8.91E-03 | 0.0817 |
| HOMA-IR \|\| id:ieu-b-118 | Inverse variance weighted | 11 | 5.25E-01 | 6.12E-03 | -1.27E-02 | 2.50E-02 | 4.82E-01 | | |
| Inflammatory bowel disease \|\| id:ebi-a-GCST004131 | MR Egger | 18 | 2.06E-01 | 1.73E-01 | -8.39E-02 | 4.29E-01 | 3.03E-41 | -1.42E-02 | 0.5918 |
| Inflammatory bowel disease \|\| id:ebi-a-GCST004131 | Inverse variance weighted | 18 | 1.21E-01 | 1.14E-01 | -3.03E-02 | 2.59E-01 | 1.49E-41 | | |
| Intelligence \|\| id:ebi-a-GCST006250 | MR Egger | 18 | 2.30E-01 | -1.34E-02 | -3.45E-02 | 7.64E-03 | 8.08E-03 | 1.88E-04 | 0.9305 |
| Intelligence \|\| id:ebi-a-GCST006250 | Inverse variance weighted | 18 | 3.31E-02 | -1.26E-02 | -2.43E-02 | -1.01E-03 | 1.22E-02 | | |
| Ischemic stroke \|\| id:ebi-a-GCST005843 | MR Egger | 20 | 9.09E-01 | -3.78E-03 | -6.80E-02 | 6.04E-02 | 2.64E-01 | 2.55E-03 | 0.6541 |

| outcome | method | nsnp | pval | b | lo ci | up ci | Q pval | egger intercept | ple pval |
|---|---|---|---|---|---|---|---|---|---|
| Ischemic stroke \|\| id:ebi-a-GCST005843 | Inverse variance weighted | 20 | 5.68E-01 | 9.14E-03 | -2.22E-02 | 4.05E-02 | 3.06E-01 | | |
| Loneliness, isolation \|\| id:ukb-b-8476 | MR Egger | 21 | 6.49E-03 | 7.19E-03 | 2.58E-03 | 1.18E-02 | 1.85E-01 | -7.57E-04 | 0.1191 |
| Loneliness, isolation \|\| id:ukb-b-8476 | Inverse variance weighted | 21 | 3.25E-03 | 4.00E-03 | 1.33E-03 | 6.66E-03 | 1.16E-01 | | |
| Long-standing illness, disability or infirmity \|\| id:ukb-b-13764 | MR Egger | 21 | 9.21E-01 | -6.73E-04 | -1.38E-02 | 1.24E-02 | 7.53E-19 | 7.11E-04 | 0.5955 |
| Long-standing illness, disability or infirmity \|\| id:ukb-b-13764 | Inverse variance weighted | 21 | 5.22E-01 | 2.33E-03 | -4.81E-03 | 9.46E-03 | 8.45E-19 | | |
| Loud music exposure frequency \|\| id:ukb-b-5436 | MR Egger | 21 | 5.45E-01 | -4.27E-03 | -1.78E-02 | 9.30E-03 | 6.92E-01 | 1.97E-03 | 0.1655 |
| Loud music exposure frequency \|\| id:ukb-b-5436 | Inverse variance weighted | 21 | 2.93E-01 | 4.04E-03 | -3.49E-03 | 1.16E-02 | 6.17E-01 | | |
| Major depressive disorder (ICD-10 coded) \|\| id:ebi-a-GCST005903 | MR Egger | 20 | 4.38E-01 | 1.46E-03 | -2.14E-03 | 5.06E-03 | 9.59E-02 | -1.16E-04 | 0.7498 |
| Major depressive disorder (ICD-10 coded) \|\| id:ebi-a-GCST005903 | Inverse variance weighted | 20 | 3.43E-01 | 9.68E-04 | -1.03E-03 | 2.97E-03 | 1.21E-01 | | |
| Maximum heart rate during fitness test \|\| id:ukb-b-14461 | MR Egger | 21 | 7.21E-01 | 4.97E-03 | -2.19E-02 | 3.18E-02 | 4.37E-01 | 2.97E-04 | 0.9136 |
| Maximum heart rate during fitness test \|\| id:ukb-b-14461 | Inverse variance weighted | 21 | 4.09E-01 | 6.22E-03 | -8.55E-03 | 2.10E-02 | 5.01E-01 | | |
| Miserableness \|\| id:ukb-b-18994 | MR Egger | 21 | 5.52E-05 | 1.54E-02 | 9.54E-03 | 2.12E-02 | 1.98E-01 | -1.42E-03 | 0.0261 |
| Miserableness \|\| id:ukb-b-18994 | Inverse variance weighted | 21 | 3.27E-07 | 9.40E-03 | 5.79E-03 | 1.30E-02 | 5.16E-02 | | |
| Mood swings \|\| id:ukb-b-14180 | MR Egger | 21 | 9.44E-03 | 1.09E-02 | 3.50E-03 | 1.83E-02 | 7.53E-03 | -1.45E-03 | 0.0659 |
| Mood swings \|\| id:ukb-b-14180 | Inverse variance weighted | 21 | 3.29E-02 | 4.77E-03 | 3.86E-04 | 9.15E-03 | 1.24E-03 | | |
| Neuroticism \|\| id:ebi-a-GCST005232 | MR Egger | 19 | 1.43E-03 | 3.53E-02 | 1.71E-02 | 5.35E-02 | 1.97E-01 | -4.31E-03 | 0.0967 |
| Neuroticism \|\| id:ebi-a-GCST005232 | Inverse variance weighted | 19 | 1.53E-04 | 2.22E-02 | 1.07E-02 | 3.37E-02 | 1.08E-01 | | |
| Neuroticism score \|\| id:ukb-b-4630 | MR Egger | 21 | 4.48E-03 | 7.74E-02 | 3.03E-02 | 1.24E-01 | 5.26E-02 | -9.24E-03 | 0.0658 |
| Neuroticism score \|\| id:ukb-b-4630 | Inverse variance weighted | 21 | 6.98E-03 | 3.84E-02 | 1.05E-02 | 6.63E-02 | 1.56E-02 | | |

| outcome | method | nsnp | pval | b | lo ci | up ci | Q pval | egger intercept | ple pval |
|---|---|---|---|---|---|---|---|---|---|
| Number of operations, self-reported \|\| id:ukb-b-4733 | MR Egger | 21 | 2.27E-01 | 5.49E-03 | -3.13E-03 | 1.41E-02 | 3.47E-01 | -1.34E-03 | 0.1393 |
| Number of operations, self-reported \|\| id:ukb-b-4733 | Inverse variance weighted | 21 | 9.51E-01 | -1.56E-04 | -5.10E-03 | 4.78E-03 | 2.68E-01 | | |
| Other serious medical condition/disability diagnosed by doctor \|\| id:ukb-b-14961 | MR Egger | 21 | 2.09E-01 | -4.20E-03 | -1.05E-02 | 2.13E-03 | 2.27E-03 | 1.03E-03 | 0.1231 |
| Other serious medical condition/disability diagnosed by doctor \|\| id:ukb-b-14961 | Inverse variance weighted | 21 | 9.40E-01 | 1.39E-04 | -3.51E-03 | 3.79E-03 | 6.16E-04 | | |
| Parkinson's disease \|\| id:ieu-b-7 | MR Egger | 21 | 7.62E-01 | 1.59E-02 | -8.55E-02 | 1.17E-01 | 8.32E-01 | -8.37E-03 | 0.3951 |
| Parkinson's disease \|\| id:ieu-b-7 | Inverse variance weighted | 21 | 4.56E-01 | -2.15E-02 | -7.80E-02 | 3.50E-02 | 8.36E-01 | | |
| Pulse wave Arterial Stiffness index \|\| id:ukb-b-11971 | MR Egger | 21 | 4.66E-01 | 7.99E-03 | -1.31E-02 | 2.90E-02 | 1.28E-01 | -1.44E-03 | 0.5060 |
| Pulse wave Arterial Stiffness index \|\| id:ukb-b-11971 | Inverse variance weighted | 21 | 7.42E-01 | 1.94E-03 | -9.58E-03 | 1.35E-02 | 1.44E-01 | | |
| Pulse wave peak to peak time \|\| id:ukb-b-8778 | MR Egger | 21 | 3.99E-01 | -9.35E-03 | -3.06E-02 | 1.19E-02 | 1.43E-01 | 1.44E-03 | 0.5090 |
| Pulse wave peak to peak time \|\| id:ukb-b-8778 | Inverse variance weighted | 21 | 5.79E-01 | -3.29E-03 | -1.49E-02 | 8.33E-03 | 1.60E-01 | | |
| Qualifications: A levels/AS levels or equivalent \|\| id:ukb-b-11615 | MR Egger | 21 | 4.76E-02 | -6.04E-03 | -1.16E-02 | -4.50E-04 | 9.77E-02 | 4.50E-04 | 0.4329 |
| Qualifications: A levels/AS levels or equivalent \|\| id:ukb-b-11615 | Inverse variance weighted | 21 | 8.29E-03 | -4.14E-03 | -7.21E-03 | -1.07E-03 | 1.04E-01 | | |
| Qualifications: College or University degree \|\| id:ukb-b-16489 | MR Egger | 21 | 6.15E-02 | -7.49E-03 | -1.49E-02 | -1.04E-04 | 7.61E-04 | 3.85E-04 | 0.6109 |
| Qualifications: College or University degree \|\| id:ukb-b-16489 | Inverse variance weighted | 21 | 4.24E-03 | -5.87E-03 | -9.89E-03 | -1.85E-03 | 1.00E-03 | | |
| Qualifications: CSEs or equivalent \|\| id:ukb-b-10505 | MR Egger | 21 | 6.45E-01 | 8.53E-04 | -2.72E-03 | 4.43E-03 | 4.54E-01 | 1.29E-04 | 0.7241 |
| Qualifications: CSEs or equivalent \|\| id:ukb-b-10505 | Inverse variance weighted | 21 | 1.67E-01 | 1.40E-03 | -5.84E-04 | 3.38E-03 | 5.11E-01 | | |
| Qualifications: None of the above \|\| id:ukb-b-17729 | MR Egger | 21 | 1.19E-02 | 7.23E-03 | 2.13E-03 | 1.23E-02 | 3.80E-02 | -6.81E-04 | 0.1995 |

| outcome | method | nsnp | pval | b | lo ci | up ci | Q pval | egger intercept | ple pval |
|---|---|---|---|---|---|---|---|---|---|
| Qualifications: None of the above \|\| id:ukb-b-17729 | Inverse variance weighted | 21 | 3.03E-03 | 4.35E-03 | 1.48E-03 | 7.23E-03 | 2.52E-02 | | |
| Qualifications: NVQ or HND or HNC or equivalent \|\| id:ukb-b-7926 | MR Egger | 21 | 1.03E-01 | 4.84E-03 | -7.01E-04 | 1.04E-02 | 1.50E-02 | -5.40E-04 | 0.3445 |
| Qualifications: NVQ or HND or HNC or equivalent \|\| id:ukb-b-7926 | Inverse variance weighted | 21 | 1.02E-01 | 2.56E-03 | -5.10E-04 | 5.62E-03 | 1.35E-02 | | |
| Qualifications: O levels/GCSEs or equivalent \|\| id:ukb-b-18099 | MR Egger | 21 | 5.63E-03 | -9.06E-03 | -1.48E-02 | -3.37E-03 | 2.70E-01 | 1.16E-03 | 0.0564 |
| Qualifications: O levels/GCSEs or equivalent \|\| id:ukb-b-18099 | Inverse variance weighted | 21 | 1.65E-02 | -4.15E-03 | -7.55E-03 | -7.58E-04 | 1.31E-01 | | |
| Qualifications: Other professional qualifications eg: nursing, teaching \|\| id:ukb-b-13799 | MR Egger | 21 | 1.70E-02 | -7.01E-03 | -1.23E-02 | -1.76E-03 | 2.39E-01 | 5.99E-04 | 0.2711 |
| Qualifications: Other professional qualifications eg: nursing, teaching \|\| id:ukb-b-13799 | Inverse variance weighted | 21 | 2.74E-03 | -4.48E-03 | -7.42E-03 | -1.55E-03 | 2.21E-01 | | |
| Reason for glasses/contact lenses: For short-sightedness, i.e. only or mainly for distance viewing such as driving, cinema etc (called 'myopia') \|\| id:ukb-b-6353 | MR Egger | 21 | 2.68E-01 | -2.13E-03 | -5.78E-03 | 1.53E-03 | 4.40E-02 | 3.22E-04 | 0.3917 |
| Reason for glasses/contact lenses: For short-sightedness, i.e. only or mainly for distance viewing such as driving, cinema etc (called 'myopia') \|\| id:ukb-b-6353 | Inverse variance weighted | 21 | 4.55E-01 | -7.67E-04 | -2.78E-03 | 1.25E-03 | 4.44E-02 | | |
| Seen a psychiatrist for nerves, anxiety, tension or depression \|\| id:ukb-b-18336 | MR Egger | 21 | 4.66E-02 | 4.89E-03 | 3.88E-04 | 9.40E-03 | 1.97E-02 | -4.81E-04 | 0.3018 |
| Seen a psychiatrist for nerves, anxiety, tension or depression \|\| id:ukb-b-18336 | Inverse variance weighted | 21 | 2.52E-02 | 2.86E-03 | 3.57E-04 | 5.37E-03 | 1.65E-02 | | |

| outcome | method | nsnp | pval | b | lo ci | up ci | Q pval | egger intercept | ple pval |
|---|---|---|---|---|---|---|---|---|---|
| Seen doctor (GP) for nerves, anxiety, tension or depression \|\| id:ukb-b-6991 | MR Egger | 21 | 5.79E-04 | 1.26E-02 | 6.29E-03 | 1.77E-02 | 1.54E-01 | -1.29E-03 | 0.0364 |
| Seen doctor (GP) for nerves, anxiety, tension or depression \|\| id:ukb-b-6991 | Inverse variance weighted | 21 | 2.20E-04 | 6.54E-03 | 3.07E-03 | 1.00E-02 | 4.40E-02 | | |
| Sensitivity / hurt feelings \|\| id:ukb-b-9981 | MR Egger | 21 | 1.82E-02 | 7.40E-03 | 1.79E-03 | 1.30E-02 | 2.78E-01 | -6.91E-04 | 0.2358 |
| Sensitivity / hurt feelings \|\| id:ukb-b-9981 | Inverse variance weighted | 21 | 5.30E-03 | 4.49E-03 | 1.33E-03 | 7.64E-03 | 2.48E-01 | | |
| Sleeplessness / insomnia \|\| id:ukb-b-3957 | MR Egger | 21 | 9.64E-01 | -1.88E-04 | -8.36E-03 | 7.99E-03 | 2.65E-01 | -3.05E-04 | 0.7146 |
| Sleeplessness / insomnia \|\| id:ukb-b-3957 | Inverse variance weighted | 21 | 5.14E-01 | -1.48E-03 | -5.91E-03 | 2.96E-03 | 3.11E-01 | | |
| Sodium in urine \|\| id:ukb-a-335 | MR Egger | 20 | 1.66E-02 | 2.13E-02 | 5.49E-03 | 3.71E-02 | 3.18E-02 | -1.63E-03 | 0.3140 |
| Sodium in urine \|\| id:ukb-a-335 | Inverse variance weighted | 20 | 1.71E-03 | 1.44E-02 | 5.41E-03 | 2.35E-02 | 2.78E-02 | | |
| Suffer from 'nerves' \|\| id:ukb-b-19957 | MR Egger | 21 | 3.61E-02 | 5.05E-03 | 6.63E-04 | 9.44E-03 | 5.13E-01 | -1.03E-04 | 0.8182 |
| Suffer from 'nerves' \|\| id:ukb-b-19957 | Inverse variance weighted | 21 | 2.02E-04 | 4.62E-03 | 2.18E-03 | 7.05E-03 | 5.74E-01 | | |
| Tinnitus: Yes, but not now, but have in the past \|\| id:ukb-d-4803_14 | MR Egger | 20 | 8.19E-01 | 8.49E-04 | -6.33E-03 | 8.03E-03 | 2.76E-01 | -3.51E-04 | 0.6298 |
| Tinnitus: Yes, but not now, but have in the past \|\| id:ukb-d-4803_14 | Inverse variance weighted | 20 | 7.59E-01 | -6.28E-04 | -4.64E-03 | 3.38E-03 | 3.17E-01 | | |
| Tinnitus: Yes, now most or all of the time \|\| id:ukb-d-4803_11 | MR Egger | 20 | 9.46E-01 | 1.83E-04 | -5.02E-03 | 5.39E-03 | 5.00E-01 | 2.17E-04 | 0.6800 |
| Tinnitus: Yes, now most or all of the time \|\| id:ukb-d-4803_11 | Inverse variance weighted | 20 | 4.68E-01 | 1.10E-03 | -1.87E-03 | 4.06E-03 | 5.55E-01 | | |
| triglycerides \|\| id:ieu-b-111 | MR Egger | 21 | 3.91E-01 | 1.67E-02 | -2.05E-02 | 5.38E-02 | 5.67E-44 | -1.53E-03 | 0.6863 |
| triglycerides \|\| id:ieu-b-111 | Inverse variance weighted | 21 | 3.22E-01 | 1.02E-02 | -9.99E-03 | 3.04E-02 | 7.33E-44 | | |
| Type 2 diabetes \|\| id:ebi-a-GCST006867 | MR Egger | 12 | 7.82E-01 | 2.24E-02 | -1.32E-01 | 1.77E-01 | 8.39E-01 | -6.00E-03 | 0.5903 |
| Type 2 diabetes \|\| id:ebi-a-GCST006867 | Inverse variance weighted | 12 | 3.09E-01 | -2.01E-02 | -5.88E-02 | 1.86E-02 | 8.72E-01 | | |

| outcome | method | nsnp | pval | b | lo ci | up ci | Q pval | egger intercept | ple pval |
|---|---|---|---|---|---|---|---|---|---|
| Vascular/heart problems diagnosed by doctor: Angina \|\| id:ukb-b-8468 | MR Egger | 21 | 9.21E-01 | 1.07E-04 | -1.99E-03 | 2.21E-03 | 1.82E-01 | -3.66E-05 | 0.8644 |
| Vascular/heart problems diagnosed by doctor: Angina \|\| id:ukb-b-8468 | Inverse variance weighted | 21 | 9.35E-01 | -4.72E-05 | -1.18E-03 | 1.09E-03 | 2.24E-01 | | |
| Vascular/heart problems diagnosed by doctor: High blood pressure \|\| id:ukb-b-14177 | MR Egger | 21 | 6.03E-01 | 2.74E-03 | -7.42E-03 | 1.29E-02 | 1.27E-11 | -1.09E-03 | 0.2994 |
| Vascular/heart problems diagnosed by doctor: High blood pressure \|\| id:ukb-b-14177 | Inverse variance weighted | 21 | 5.19E-01 | -1.86E-03 | -7.52E-03 | 3.80E-03 | 3.02E-12 | | |
| Vascular/heart problems diagnosed by doctor: None of the above \|\| id:ukb-b-13352 | MR Egger | 21 | 6.56E-01 | -2.41E-03 | -1.29E-02 | 8.04E-03 | 1.19E-11 | 1.08E-03 | 0.3172 |
| Vascular/heart problems diagnosed by doctor: None of the above \|\| id:ukb-b-13352 | Inverse variance weighted | 21 | 4.69E-01 | 2.15E-03 | -3.66E-03 | 7.95E-03 | 3.31E-12 | | |
| Waist-to-hip ratio \|\| id:ieu-a-72 | MR Egger | 12 | 2.79E-01 | -3.78E-02 | -1.03E-01 | 2.69E-02 | 5.82E-01 | 3.88E-03 | 0.4359 |
| Waist-to-hip ratio \|\| id:ieu-a-72 | Inverse variance weighted | 12 | 2.18E-01 | -1.23E-02 | -3.18E-02 | 7.25E-03 | 6.09E-01 | | |
| Wheeze or whistling in the chest in last year \|\| id:ukb-b-18335 | MR Egger | 21 | 7.27E-01 | 2.65E-03 | -1.20E-02 | 1.73E-02 | 5.12E-36 | 2.12E-04 | 0.8869 |
| Wheeze or whistling in the chest in last year \|\| id:ukb-b-18335 | Inverse variance weighted | 21 | 3.80E-01 | 3.54E-03 | -4.37E-03 | 1.14E-02 | 1.58E-35 | | |
| Whole body fat mass \|\| id:ukb-b-19393 | MR Egger | 21 | 4.05E-01 | 6.03E-03 | -7.84E-03 | 1.99E-02 | 7.26E-03 | -1.60E-03 | 0.2647 |
| Whole body fat mass \|\| id:ukb-b-19393 | Inverse variance weighted | 21 | 8.53E-01 | -7.35E-04 | -8.49E-03 | 7.02E-03 | 5.14E-03 | | |
| Whole body fat-free mass \|\| id:ukb-b-13354 | MR Egger | 21 | 7.33E-01 | -2.07E-03 | -1.38E-02 | 9.67E-03 | 3.45E-07 | -1.05E-05 | 0.9930 |
| Whole body fat-free mass \|\| id:ukb-b-13354 | Inverse variance weighted | 21 | 5.14E-01 | -2.12E-03 | -8.47E-03 | 4.24E-03 | 6.66E-07 | | |
| Years of schooling \|\| id:ieu-a-1239 | MR Egger | 17 | 9.50E-01 | -9.45E-04 | -3.02E-02 | 2.83E-02 | 7.42E-02 | -1.49E-04 | 0.9460 |
| Years of schooling \|\| id:ieu-a-1239 | Inverse variance weighted | 17 | 6.68E-01 | -1.92E-03 | -1.07E-02 | 6.87E-03 | 1.01E-01 | | |

**Table S7. Full list of FinnGen authors and their affiliations.**

| Full Name | Affiliation | Role 1 | Role 2 |
|---|---|---|---|
| Aarno Palotie | Institute for Molecular Medicine Finland (FIMM), HiLIFE, University of Helsinki, Helsinki, Finland; Broad Institute of MIT and Harvard; Massachusetts General Hospital | Steering Committee | Steering Committee |
| Mark Daly | Institute for Molecular Medicine Finland (FIMM), HiLIFE, University of Helsinki, Helsinki, Finland; Broad Institute of MIT and Harvard; Massachusetts General Hospital | Steering Committee | Steering Committee |
| Bridget Riley-Gills | Abbvie, Chicago, IL, United States | Steering Committee | Pharmaceutical companies |
| Howard Jacob | Abbvie, Chicago, IL, United States | Steering Committee | Pharmaceutical companies |
| Dirk Paul | Astra Zeneca, Cambridge, United Kingdom | Steering Committee | Pharmaceutical companies |
| Slavé Petrovski | Astra Zeneca, Cambridge, United Kingdom | Steering Committee | Pharmaceutical companies |
| Heiko Runz | Biogen, Cambridge, MA, United States | Steering Committee | Pharmaceutical companies |
| Sally John | Biogen, Cambridge, MA, United States | Steering Committee | Pharmaceutical companies |
| George Okafo | Boehringer Ingelheim, Ingelheim am Rhein, Germany | Steering Committee | Pharmaceutical companies |
| Nathan Lawless | Boehringer Ingelheim, Ingelheim am Rhein, Germany | Steering Committee | Pharmaceutical companies |
| Heli Salminen-Mankonen | Boehringer Ingelheim, Ingelheim am Rhein, Germany | Steering Committee | Pharmaceutical companies |
| Robert Plenge | Bristol Myers Squibb, New York, NY, United States | Steering Committee | Pharmaceutical companies |
| Joseph Maranville | Bristol Myers Squibb, New York, NY, United States | Steering Committee | Pharmaceutical companies |
| Mark McCarthy | Genentech, San Francisco, CA, United States | Steering Committee | Pharmaceutical companies |
| Margaret G. Ehm | GlaxoSmithKline, Collegeville, PA, United States | Steering Committee | Pharmaceutical companies |
| Kirsi Auro | GlaxoSmithKline, Espoo, Finland | Steering Committee | Pharmaceutical companies |
| Simonne Longerich | Merck, Kenilworth, NJ, United States | Steering Committee | Pharmaceutical companies |
| Anders Mälarstig | Pfizer, New York, NY, United States | Steering Committee | Pharmaceutical companies |

| | | | |
|---|---|---|---|
| Katherine Klinger | Translational Sciences, Sanofi R&D, Framingham, MA, USA | Steering Committee | **Pharmaceutical companies** |
| Clement Chatelain | Translational Sciences, Sanofi R&D, Framingham, MA, USA | Steering Committee | **Pharmaceutical companies** |
| Matthias Gossel | Translational Sciences, Sanofi R&D, Framingham, MA, USA | Steering Committee | **Pharmaceutical companies** |
| Karol Estrada | Maze Therapeutics, San Francisco, CA, United States | Steering Committee | **Pharmaceutical companies** |
| Robert Graham | Maze Therapeutics, San Francisco, CA, United States | Steering Committee | **Pharmaceutical companies** |
| Robert Yang | Janssen Biotech, Beerse, Belgium | Steering Committee | **Pharmaceutical companies** |
| Chris O´Donnell | Novartis Institutes for BioMedical Research, Cambridge, MA, United States | Steering Committee | **Pharmaceutical companies** |
| Tomi P. Mäkelä | HiLIFE, University of Helsinki, Finland, Finland | Steering Committee | **University of Helsinki & Biobanks** |
| Jaakko Kaprio | Institute for Molecular Medicine Finland (FIMM), HiLIFE, University of Helsinki, Helsinki, Finland | Steering Committee | **University of Helsinki & Biobanks** |
| Petri Virolainen | Auria Biobank / University of Turku / Hospital District of Southwest Finland, Turku, Finland | Steering Committee | **University of Helsinki & Biobanks** |
| Antti Hakanen | Auria Biobank / University of Turku / Hospital District of Southwest Finland, Turku, Finland | Steering Committee | **University of Helsinki & Biobanks** |
| Terhi Kilpi | THL Biobank / Finnish Institute for Health and Welfare (THL), Helsinki, Finland | Steering Committee | **University of Helsinki & Biobanks** |
| Markus Perola | THL Biobank / Finnish Institute for Health and Welfare (THL), Helsinki, Finland | Steering Committee | **University of Helsinki & Biobanks** |
| Jukka Partanen | Finnish Red Cross Blood Service / Finnish Hematology Registry and Clinical Biobank, Helsinki, Finland | Steering Committee | **University of Helsinki & Biobanks** |
| Anne Pitkäranta | Helsinki Biobank / Helsinki University and Hospital District of Helsinki and Uusimaa, Helsinki | Steering Committee | **University of Helsinki & Biobanks** |
| Taneli Raivio | Helsinki Biobank / Helsinki University and Hospital District of Helsinki and Uusimaa, Helsinki | Steering Committee | **University of Helsinki & Biobanks** |
| Jani Tikkanen | Northern Finland Biobank Borealis / University of Oulu / Northern Ostrobothnia Hospital District, Oulu, Finland | Steering Committee | **University of Helsinki & Biobanks** |
| Raisa Serpi | Northern Finland Biobank Borealis / University of Oulu / Northern Ostrobothnia Hospital District, Oulu, Finland | Steering Committee | **University of Helsinki & Biobanks** |

| | | | |
|---|---|---|---|
| Tarja Laitinen | Finnish Clinical Biobank Tampere / University of Tampere / Pirkanmaa Hospital District, Tampere, Finland | **Steering Committee** | **University of Helsinki & Biobanks** |
| Veli-Matti Kosma | Biobank of Eastern Finland / University of Eastern Finland / Northern Savo Hospital District, Kuopio, Finland | **Steering Committee** | **University of Helsinki & Biobanks** |
| Jari Laukkanen | Central Finland Biobank / University of Jyväskylä / Central Finland Health Care District, Jyväskylä, Finland | **Steering Committee** | **University of Helsinki & Biobanks** |
| Marco Hautalahti | FINBB - Finnish biobank cooperative | **Steering Committee** | **University of Helsinki & Biobanks** |
| Outi Tuovila | Business Finland, Helsinki, Finland | **Steering Committee** | **Other Experts/ Non-Voting Members** |
| Raimo Pakkanen | Business Finland, Helsinki, Finland | **Steering Committee** | **Other Experts/ Non-Voting Members** |
| Jeffrey Waring | Abbvie, Chicago, IL, United States | **Scientific Committee** | **Pharmaceutical companies** |
| Bridget Riley-Gillis | Abbvie, Chicago, IL, United States | **Scientific Committee** | **Pharmaceutical companies** |
| Fedik Rahimov | Abbvie, Chicago, IL, United States | **Scientific Committee** | **Pharmaceutical companies** |
| Ioanna Tachmazidou | Astra Zeneca, Cambridge, United Kingdom | **Scientific Committee** | **Pharmaceutical companies** |
| Chia-Yen Chen | Biogen, Cambridge, MA, United States | **Scientific Committee** | **Pharmaceutical companies** |
| Heiko Runz | Biogen, Cambridge, MA, United States | **Scientific Committee** | **Pharmaceutical companies** |
| Zhihao Ding | Boehringer Ingelheim, Ingelheim am Rhein, Germany | **Scientific Committee** | **Pharmaceutical companies** |
| Marc Jung | Boehringer Ingelheim, Ingelheim am Rhein, Germany | **Scientific Committee** | **Pharmaceutical companies** |
| Shameek Biswas | Bristol Myers Squibb, New York, NY, United States | **Scientific Committee** | **Pharmaceutical companies** |
| Rion Pendergrass | Genentech, San Francisco, CA, United States | **Scientific Committee** | **Pharmaceutical companies** |
| Margaret G. Ehm | GlaxoSmithKline, Collegeville, PA, United States | **Scientific Committee** | **Pharmaceutical companies** |
| David Pulford | GlaxoSmithKline, Stevenage, United Kingdom | **Scientific Committee** | **Pharmaceutical companies** |
| Neha Raghavan | Merck, Kenilworth, NJ, United States | **Scientific Committee** | **Pharmaceutical companies** |
| Adriana Huertas-Vazquez | Merck, Kenilworth, NJ, United States | **Scientific Committee** | **Pharmaceutical companies** |

| | | | |
|---|---|---|---|
| Jae-Hoon Sul | Merck, Kenilworth, NJ, United States | Scientific Committee | Pharmaceutical companies |
| Anders Mälarstig | Pfizer, New York, NY, United States | Scientific Committee | Pharmaceutical companies |
| Xinli Hu | Pfizer, New York, NY, United States | Scientific Committee | Pharmaceutical companies |
| Åsa Hedman | Pfizer, New York, NY, United States | Scientific Committee | Pharmaceutical companies |
| Katherine Klinger | Translational Sciences, Sanofi R&D, Framingham, MA, USA | Scientific Committee | Pharmaceutical companies |
| Robert Graham | Maze Therapeutics, San Francisco, CA, United States | Scientific Committee | Pharmaceutical companies |
| Manuel Rivas | Maze Therapeutics, San Francisco, CA, United States | Scientific Committee | Pharmaceutical companies |
| Dawn Waterworth | Janssen Research & Development, LLC, Spring House, PA, United States | Scientific Committee | Pharmaceutical companies |
| Nicole Renaud | Novartis Institutes for BioMedical Research, Cambridge, MA, United States | Scientific Committee | Pharmaceutical companies |
| Ma´en Obeidat | Novartis Institutes for BioMedical Research, Cambridge, MA, United States | Scientific Committee | Pharmaceutical companies |
| Samuli Ripatti | Institute for Molecular Medicine Finland (FIMM), HiLIFE, University of Helsinki, Helsinki, Finland | Scientific Committee | University of Helsinki & Biobanks |
| Johanna Schleutker | Auria Biobank / Univ. of Turku / Hospital District of Southwest Finland, Turku, Finland | Scientific Committee | University of Helsinki & Biobanks |
| Markus Perola | THL Biobank / Finnish Institute for Health and Welfare (THL), Helsinki, Finland | Scientific Committee | University of Helsinki & Biobanks |
| Mikko Arvas | Finnish Red Cross Blood Service / Finnish Hematology Registry and Clinical Biobank, Helsinki, Finland | Scientific Committee | University of Helsinki & Biobanks |
| Olli Carpén | Helsinki Biobank / Helsinki University and Hospital District of Helsinki and Uusimaa, Helsinki | Scientific Committee | University of Helsinki & Biobanks |
| Reetta Hinttala | Northern Finland Biobank Borealis / University of Oulu / Northern Ostrobothnia Hospital District, Oulu, Finland | Scientific Committee | University of Helsinki & Biobanks |
| Johannes Kettunen | Northern Finland Biobank Borealis / University of Oulu / Northern Ostrobothnia Hospital District, Oulu, Finland | Scientific Committee | University of Helsinki & Biobanks |
| Arto Mannermaa | Biobank of Eastern Finland / University of Eastern Finland / Northern Savo Hospital District, Kuopio, Finland | Scientific Committee | University of Helsinki & Biobanks |
| Katriina Aalto-Setälä | Faculty of Medicine and Health Technology, Tampere University, Tampere, Finland | Scientific Committee | University of Helsinki & Biobanks |

| | | | |
|---|---|---|---|
| Mika Kähönen | Finnish Clinical Biobank Tampere / University of Tampere / Pirkanmaa Hospital District, Tampere, Finland | Scientific Committee | **University of Helsinki & Biobanks** |
| Jari Laukkanen | Central Finland Biobank / University of Jyväskylä / Central Finland Health Care District, Jyväskylä, Finland | Scientific Committee | **University of Helsinki & Biobanks** |
| Johanna Mäkelä | FINBB - Finnish biobank cooperative | Scientific Committee | **University of Helsinki & Biobanks** |
| Reetta Kälviäinen | Northern Savo Hospital District, Kuopio, Finland | Clinical Groups | **Neurology Group** |
| Valtteri Julkunen | Northern Savo Hospital District, Kuopio, Finland | Clinical Groups | **Neurology Group** |
| Hilkka Soininen | Northern Savo Hospital District, Kuopio, Finland | Clinical Groups | **Neurology Group** |
| Anne Remes | Northern Ostrobothnia Hospital District, Oulu, Finland | Clinical Groups | **Neurology Group** |
| Mikko Hiltunen | University of Eastern Finland, Kuopio, Finland | Clinical Groups | **Neurology Group** |
| Jukka Peltola | Pirkanmaa Hospital District, Tampere, Finland | Clinical Groups | **Neurology Group** |
| Minna Raivio | Hospital District of Helsinki and Uusimaa, Helsinki, Finland | Clinical Groups | **Neurology Group** |
| Pentti Tienari | Hospital District of Helsinki and Uusimaa, Helsinki, Finland | Clinical Groups | **Neurology Group** |
| Juha Rinne | Hospital District of Southwest Finland, Turku, Finland | Clinical Groups | **Neurology Group** |
| Roosa Kallionpää | Hospital District of Southwest Finland, Turku, Finland | Clinical Groups | **Neurology Group** |
| Juulia Partanen | Institute for Molecular Medicine Finland, HiLIFE, University of Helsinki, Finland | Clinical Groups | **Neurology Group** |
| Ali Abbasi | Abbvie, Chicago, IL, United States | Clinical Groups | **Neurology Group** |
| Adam Ziemann | Abbvie, Chicago, IL, United States | Clinical Groups | **Neurology Group** |
| Nizar Smaoui | Abbvie, Chicago, IL, United States | Clinical Groups | **Neurology Group** |
| Anne Lehtonen | Abbvie, Chicago, IL, United States | Clinical Groups | **Neurology Group** |
| Susan Eaton | Biogen, Cambridge, MA, United States | Clinical Groups | **Neurology Group** |
| Heiko Runz | Biogen, Cambridge, MA, United States | Clinical Groups | **Neurology Group** |

| | | | |
|---|---|---|---|
| Sanni Lahdenperä | Biogen, Cambridge, MA, United States | Clinical Groups | **Neurology Group** |
| Shameek Biswas | Bristol Myers Squibb, New York, NY, United States | Clinical Groups | **Neurology Group** |
| Natalie Bowers | Genentech, San Francisco, CA, United States | Clinical Groups | **Neurology Group** |
| Edmond Teng | Genentech, San Francisco, CA, United States | Clinical Groups | **Neurology Group** |
| Rion Pendergrass | Genentech, San Francisco, CA, United States | Clinical Groups | **Neurology Group** |
| Fanli Xu | GlaxoSmithKline, Brentford, United Kingdom | Clinical Groups | **Neurology Group** |
| David Pulford | GlaxoSmithKline, Stevenage, United Kingdom | Clinical Groups | **Neurology Group** |
| Kirsi Auro | GlaxoSmithKline, Espoo, Finland | Clinical Groups | **Neurology Group** |
| Laura Addis | GlaxoSmithKline, Brentford, United Kingdom | Clinical Groups | **Neurology Group** |
| John Eicher | GlaxoSmithKline, Brentford, United Kingdom | Clinical Groups | **Neurology Group** |
| Qingqin S Li | Janssen Research & Development, LLC, Titusville, NJ 08560, United States | Clinical Groups | **Neurology Group** |
| Karen He | Janssen Research & Development, LLC, Spring House, PA, United States | Clinical Groups | **Neurology Group** |
| Ekaterina Khramtsova | Janssen Research & Development, LLC, Spring House, PA, United States | Clinical Groups | **Neurology Group** |
| Neha Raghavan | Merck, Kenilworth, NJ, United States | Clinical Groups | **Neurology Group** |
| Martti Färkkilä | Hospital District of Helsinki and Uusimaa, Helsinki, Finland | Clinical Groups | **Gastroenterology Group** |
| Jukka Koskela | Hospital District of Helsinki and Uusimaa, Helsinki, Finland | Clinical Groups | **Gastroenterology Group** |
| Sampsa Pikkarainen | Hospital District of Helsinki and Uusimaa, Helsinki, Finland | Clinical Groups | **Gastroenterology Group** |
| Airi Jussila | Pirkanmaa Hospital District, Tampere, Finland | Clinical Groups | **Gastroenterology Group** |
| Katri Kaukinen | Pirkanmaa Hospital District, Tampere, Finland | Clinical Groups | **Gastroenterology Group** |
| Timo Blomster | Northern Ostrobothnia Hospital District, Oulu, Finland | Clinical Groups | **Gastroenterology Group** |

| | | | |
|---|---|---|---|
| Mikko Kiviniemi | Northern Savo Hospital District, Kuopio, Finland | Clinical Groups | **Gastroenterology Group** |
| Markku Voutilainen | Hospital District of Southwest Finland, Turku, Finland | Clinical Groups | **Gastroenterology Group** |
| Mark Daly | Institute for Molecular Medicine, Finland (FIMM), HiLIFE, University of Helsinki, Helsinki, Finland; Broad Institute of MIT and Harvard; Massachusetts General Hospital | Clinical Groups | **Gastroenterology Group** |
| Ali Abbasi | Abbvie, Chicago, IL, United States | Clinical Groups | **Gastroenterology Group** |
| Jeffrey Waring | Abbvie, Chicago, IL, United States | Clinical Groups | **Gastroenterology Group** |
| Nizar Smaoui | Abbvie, Chicago, IL, United States | Clinical Groups | **Gastroenterology Group** |
| Fedik Rahimov | Abbvie, Chicago, IL, United States | Clinical Groups | **Gastroenterology Group** |
| Anne Lehtonen | Abbvie, Chicago, IL, United States | Clinical Groups | **Gastroenterology Group** |
| Tim Lu | Genentech, San Francisco, CA, United States | Clinical Groups | **Gastroenterology Group** |
| Natalie Bowers | Genentech, San Francisco, CA, United States | Clinical Groups | **Gastroenterology Group** |
| Rion Pendergrass | Genentech, San Francisco, CA, United States | Clinical Groups | **Gastroenterology Group** |
| Linda McCarthy | GlaxoSmithKline, Brentford, United Kingdom | Clinical Groups | **Gastroenterology Group** |
| Amy Hart | Janssen Research & Development, LLC, Spring House, PA, United States | Clinical Groups | **Gastroenterology Group** |
| Meijian Guan | Janssen Research & Development, LLC, Spring House, PA, United States | Clinical Groups | **Gastroenterology Group** |
| Jason Miller | Merck, Kenilworth, NJ, United States | Clinical Groups | **Gastroenterology Group** |
| Kirsi Kalpala | Pfizer, New York, NY, United States | Clinical Groups | **Gastroenterology Group** |
| Melissa Miller | Pfizer, New York, NY, United States | Clinical Groups | **Gastroenterology Group** |
| Xinli Hu | Pfizer, New York, NY, United States | Clinical Groups | **Gastroenterology Group** |
| Kari Eklund | Hospital District of Helsinki and Uusimaa, Helsinki, Finland | Clinical Groups | **Rheumatology Group** |
| Antti Palomäki | Hospital District of Southwest Finland, Turku, Finland | Clinical Groups | **Rheumatology Group** |

| | | | |
|---|---|---|---|
| Pia Isomäki | Pirkanmaa Hospital District, Tampere, Finland | Clinical Groups | **Rheumatology Group** |
| Laura Pirilä | Hospital District of Southwest Finland, Turku, Finland | Clinical Groups | **Rheumatology Group** |
| Oili Kaipiainen-Seppänen | Northern Savo Hospital District, Kuopio, Finland | Clinical Groups | **Rheumatology Group** |
| Johanna Huhtakangas | Northern Ostrobothnia Hospital District, Oulu, Finland | Clinical Groups | **Rheumatology Group** |
| Nina Mars | Institute for Molecular Medicine Finland (FIMM), HiLIFE, University of Helsinki, Helsinki, Finland | Clinical Groups | **Rheumatology Group** |
| Ali Abbasi | Abbvie, Chicago, IL, United States | Clinical Groups | **Rheumatology Group** |
| Jeffrey Waring | Abbvie, Chicago, IL, United States | Clinical Groups | **Rheumatology Group** |
| Fedik Rahimov | Abbvie, Chicago, IL, United States | Clinical Groups | **Rheumatology Group** |
| Apinya Lertratanakul | Abbvie, Chicago, IL, United States | Clinical Groups | **Rheumatology Group** |
| Nizar Smaoui | Abbvie, Chicago, IL, United States | Clinical Groups | **Rheumatology Group** |
| Anne Lehtonen | Abbvie, Chicago, IL, United States | Clinical Groups | **Rheumatology Group** |
| Coralie Viollet | AstraZeneca, Cambridge, United Kingdom | Clinical Groups | **Rheumatology Group** |
| Marla Hochfeld | Bristol Myers Squibb, New York, NY, United States | Clinical Groups | **Rheumatology Group** |
| Natalie Bowers | Genentech, San Francisco, CA, United States | Clinical Groups | **Rheumatology Group** |
| Rion Pendergrass | Genentech, San Francisco, CA, United States | Clinical Groups | **Rheumatology Group** |
| Jorge Esparza Gordillo | GlaxoSmithKline, Brentford, United Kingdom | Clinical Groups | **Rheumatology Group** |
| Kirsi Auro | GlaxoSmithKline, Espoo, Finland | Clinical Groups | **Rheumatology Group** |
| Dawn Waterworth | Janssen Research & Development, LLC, Spring House, PA, United States | Clinical Groups | **Rheumatology Group** |
| Fabiana Farias | Merck, Kenilworth, NJ, United States | Clinical Groups | **Rheumatology Group** |
| Kirsi Kalpala | Pfizer, New York, NY, United States | Clinical Groups | **Rheumatology Group** |

| | | | |
|---|---|---|---|
| Nan Bing | Pfizer, New York, NY, United States | Clinical Groups | **Rheumatology Group** |
| Xinli Hu | Pfizer, New York, NY, United States | Clinical Groups | **Rheumatology Group** |
| Tarja Laitinen | Pirkanmaa Hospital District, Tampere, Finland | Clinical Groups | **Pulmonology Group** |
| Margit Pelkonen | Northern Savo Hospital District, Kuopio, Finland | Clinical Groups | **Pulmonology Group** |
| Paula Kauppi | Hospital District of Helsinki and Uusimaa, Helsinki, Finland | Clinical Groups | **Pulmonology Group** |
| Hannu Kankaanranta | University of Gothenburg, Gothenburg, Sweden/ Seinäjoki Central Hospital, Seinäjoki, Finland/ Tampere University, Tampere, Finland | Clinical Groups | **Pulmonology Group** |
| Terttu Harju | Northern Ostrobothnia Hospital District, Oulu, Finland | Clinical Groups | **Pulmonology Group** |
| Riitta Lahesmaa | Hospital District of Southwest Finland, Turku, Finland | Clinical Groups | **Pulmonology Group** |
| Nizar Smaoui | Abbvie, Chicago, IL, United States | Clinical Groups | **Pulmonology Group** |
| Coralie Viollet | AstraZeneca, Cambridge, United Kingdom | Clinical Groups | **Pulmonology Group** |
| Susan Eaton | Biogen, Cambridge, MA, United States | Clinical Groups | **Pulmonology Group** |
| Hubert Chen | Genentech, San Francisco, CA, United States | Clinical Groups | **Pulmonology Group** |
| Rion Pendergrass | Genentech, San Francisco, CA, United States | Clinical Groups | **Pulmonology Group** |
| Natalie Bowers | Genentech, San Francisco, CA, United States | Clinical Groups | **Pulmonology Group** |
| Joanna Betts | GlaxoSmithKline, Brentford, United Kingdom | Clinical Groups | **Pulmonology Group** |
| Kirsi Auro | GlaxoSmithKline, Espoo, Finland | Clinical Groups | **Pulmonology Group** |
| Rajashree Mishra | GlaxoSmithKline, Brentford, United Kingdom | Clinical Groups | **Pulmonology Group** |
| Majd Mouded | Novartis, Basel, Switzerland | Clinical Groups | **Pulmonology Group** |
| Debby Ngo | Novartis, Basel, Switzerland | Clinical Groups | **Pulmonology Group** |
| Teemu Niiranen | Finnish Institute for Health and Welfare (THL), Helsinki, Finland | Clinical Groups | **Cardiometabolic Diseases Group** |

| | | | |
|---|---|---|---|
| Felix Vaura | Finnish Institute for Health and Welfare (THL), Helsinki, Finland | Clinical Groups | **Cardiometabolic Diseases Group** |
| Veikko Salomaa | Finnish Institute for Health and Welfare (THL), Helsinki, Finland | Clinical Groups | **Cardiometabolic Diseases Group** |
| Kaj Metsärinne | Hospital District of Southwest Finland, Turku, Finland | Clinical Groups | **Cardiometabolic Diseases Group** |
| Jenni Aittokallio | Hospital District of Southwest Finland, Turku, Finland | Clinical Groups | **Cardiometabolic Diseases Group** |
| Mika Kähönen | Pirkanmaa Hospital District, Tampere, Finland | Clinical Groups | **Cardiometabolic Diseases Group** |
| Jussi Hernesniemi | Pirkanmaa Hospital District, Tampere, Finland | Clinical Groups | **Cardiometabolic Diseases Group** |
| Daniel Gordin | Hospital District of Helsinki and Uusimaa, Helsinki, Finland | Clinical Groups | **Cardiometabolic Diseases Group** |
| Juha Sinisalo | Hospital District of Helsinki and Uusimaa, Helsinki, Finland | Clinical Groups | **Cardiometabolic Diseases Group** |
| Marja-Riitta Taskinen | Hospital District of Helsinki and Uusimaa, Helsinki, Finland | Clinical Groups | **Cardiometabolic Diseases Group** |
| Tiinamaija Tuomi | Hospital District of Helsinki and Uusimaa, Helsinki, Finland | Clinical Groups | **Cardiometabolic Diseases Group** |
| Timo Hiltunen | Hospital District of Helsinki and Uusimaa, Helsinki, Finland | Clinical Groups | **Cardiometabolic Diseases Group** |
| Jari Laukkanen | Central Finland Health Care District, Jyväskylä, Finland | Clinical Groups | **Cardiometabolic Diseases Group** |
| Amanda Elliott | Institute for Molecular Medicine Finland (FIMM), HiLIFE, University of Helsinki, Helsinki, Finland; Broad Institute, Cambridge, MA, USA and Massachusetts General Hospital, Boston, MA, USA | Clinical Groups | **Cardiometabolic Diseases Group** |
| Mary Pat Reeve | Institute for Molecular Medicine Finland (FIMM), HiLIFE, University of Helsinki, Helsinki, Finland | Clinical Groups | **Cardiometabolic Diseases Group** |
| Sanni Ruotsalainen | Institute for Molecular Medicine Finland (FIMM), HiLIFE, University of Helsinki, Helsinki, Finland | Clinical Groups | **Cardiometabolic Diseases Group** |
| Dirk Paul | Astra Zeneca, Cambridge, United Kingdom | Clinical Groups | **Cardiometabolic Diseases Group** |
| Natalie Bowers | Genentech, San Francisco, CA, United States | Clinical Groups | **Cardiometabolic Diseases Group** |
| Rion Pendergrass | Genentech, San Francisco, CA, United States | Clinical Groups | **Cardiometabolic Diseases Group** |
| Audrey Chu | GlaxoSmithKline, Brentford, United Kingdom | Clinical Groups | **Cardiometabolic Diseases Group** |
| Kirsi Auro | GlaxoSmithKline, Espoo, Finland | Clinical Groups | **Cardiometabolic Diseases Group** |

| | | | |
|---|---|---|---|
| Dermot Reilly | Janssen Research & Development, LLC, Boston, MA, United States | Clinical Groups | **Cardiometabolic Diseases Group** |
| Mike Mendelson | Novartis, Boston, MA, United States | Clinical Groups | **Cardiometabolic Diseases Group** |
| Jaakko Parkkinen | Pfizer, New York, NY, United States | Clinical Groups | **Cardiometabolic Diseases Group** |
| Melissa Miller | Pfizer, New York, NY, United States | Clinical Groups | **Cardiometabolic Diseases Group** |
| Tuomo Meretoja | Hospital District of Helsinki and Uusimaa, Helsinki, Finland | Clinical Groups | **Oncology Group** |
| Heikki Joensuu | Hospital District of Helsinki and Uusimaa, Helsinki, Finland | Clinical Groups | **Oncology Group** |
| Olli Carpén | Hospital District of Helsinki and Uusimaa, Helsinki, Finland | Clinical Groups | **Oncology Group** |
| Johanna Mattson | Hospital District of Helsinki and Uusimaa, Helsinki, Finland | Clinical Groups | **Oncology Group** |
| Eveliina Salminen | Hospital District of Helsinki and Uusimaa, Helsinki, Finland | Clinical Groups | **Oncology Group** |
| Annika Auranen | Pirkanmaa Hospital District , Tampere, Finland | Clinical Groups | **Oncology Group** |
| Peeter Karihtala | Northern Ostrobothnia Hospital District, Oulu, Finland | Clinical Groups | **Oncology Group** |
| Päivi Auvinen | Northern Savo Hospital District, Kuopio, Finland | Clinical Groups | **Oncology Group** |
| Klaus Elenius | Hospital District of Southwest Finland, Turku, Finland | Clinical Groups | **Oncology Group** |
| Johanna Schleutker | Hospital District of Southwest Finland, Turku, Finland | Clinical Groups | **Oncology Group** |
| Esa Pitkänen | Institute for Molecular Medicine Finland (FIMM), HiLIFE, University of Helsinki, Helsinki, Finland | Clinical Groups | **Oncology Group** |
| Nina Mars | Institute for Molecular Medicine Finland (FIMM), HiLIFE, University of Helsinki, Helsinki, Finland | Clinical Groups | **Oncology Group** |
| Mark Daly | Institute for Molecular Medicine Finland (FIMM), HiLIFE, University of Helsinki, Helsinki, Finland; Broad Institute of MIT and Harvard; Massachusetts General Hospital | Clinical Groups | **Oncology Group** |
| Relja Popovic | Abbvie, Chicago, IL, United States | Clinical Groups | **Oncology Group** |
| Jeffrey Waring | Abbvie, Chicago, IL, United States | Clinical Groups | **Oncology Group** |
| Bridget Riley-Gillis | Abbvie, Chicago, IL, United States | Clinical Groups | **Oncology Group** |

| | | | |
|---|---|---|---|
| Anne Lehtonen | Abbvie, Chicago, IL, United States | Clinical Groups | **Oncology Group** |
| Margarete Fabre | AstraZeneca, Cambridge, United Kingdom | Clinical Groups | **Oncology Group** |
| Jennifer Schutzman | Genentech, San Francisco, CA, United States | Clinical Groups | **Oncology Group** |
| Natalie Bowers | Genentech, San Francisco, CA, United States | Clinical Groups | **Oncology Group** |
| Rion Pendergrass | Genentech, San Francisco, CA, United States | Clinical Groups | **Oncology Group** |
| Diptee Kulkarni | GlaxoSmithKline, Brentford, United Kingdom | Clinical Groups | **Oncology Group** |
| Kirsi Auro | GlaxoSmithKline, Espoo, Finland | Clinical Groups | **Oncology Group** |
| Alessandro Porello | Janssen Research & Development, LLC, Spring House, PA, United States | Clinical Groups | **Oncology Group** |
| Andrey Loboda | Merck, Kenilworth, NJ, United States | Clinical Groups | **Oncology Group** |
| Heli Lehtonen | Pfizer, New York, NY, United States | Clinical Groups | **Oncology Group** |
| Stefan McDonough | Pfizer, New York, NY, United States | Clinical Groups | **Oncology Group** |
| Sauli Vuoti | Janssen-Cilag Oy, Espoo, Finland | Clinical Groups | **Oncology Group** |
| Kai Kaarniranta | Northern Savo Hospital District, Kuopio, Finland; Department of Molecular Genetics, University of Lodz, Lodz, Poland | Clinical Groups | **Opthalmology Group** |
| Joni A Turunen | Helsinki University Hospital and University of Helsinki, Helsinki, Finland; Eye Genetics Group, Folkhälsan Research Center, Helsinki, Finland | Clinical Groups | **Opthalmology Group** |
| Terhi Ollila | Hospital District of Helsinki and Uusimaa, Helsinki, Finland | Clinical Groups | **Opthalmology Group** |
| Hannu Uusitalo | Pirkanmaa Hospital District, Tampere, Finland | Clinical Groups | **Opthalmology Group** |
| Juha Karjalainen | Institute for Molecular Medicine Finland (FIMM), HiLIFE, University of Helsinki, Helsinki, Finland | Clinical Groups | **Opthalmology Group** |
| Esa Pitkänen | Institute for Molecular Medicine Finland (FIMM), HiLIFE, University of Helsinki, Helsinki, Finland | Clinical Groups | **Opthalmology Group** |
| Mengzhen Liu | Abbvie, Chicago, IL, United States | Clinical Groups | **Opthalmology Group** |
| Heiko Runz | Biogen, Cambridge, MA, United States | Clinical Groups | **Opthalmology Group** |

| | | | |
|---|---|---|---|
| Stephanie Loomis | Biogen, Cambridge, MA, United States | Clinical Groups | **Opthalmology Group** |
| Erich Strauss | Genentech, San Francisco, CA, United States | Clinical Groups | **Opthalmology Group** |
| Natalie Bowers | Genentech, San Francisco, CA, United States | Clinical Groups | **Opthalmology Group** |
| Hao Chen | Genentech, San Francisco, CA, United States | Clinical Groups | **Opthalmology Group** |
| Rion Pendergrass | Genentech, San Francisco, CA, United States | Clinical Groups | **Opthalmology Group** |
| Kaisa Tasanen | Northern Ostrobothnia Hospital District, Oulu, Finland | Clinical Groups | **Dermatology Group** |
| Laura Huilaja | Northern Ostrobothnia Hospital District, Oulu, Finland | Clinical Groups | **Dermatology Group** |
| Katariina Hannula-Jouppi | Hospital District of Helsinki and Uusimaa, Helsinki, Finland | Clinical Groups | **Dermatology Group** |
| Teea Salmi | Pirkanmaa Hospital District, Tampere, Finland | Clinical Groups | **Dermatology Group** |
| Sirkku Peltonen | Hospital District of Southwest Finland, Turku, Finland | Clinical Groups | **Dermatology Group** |
| Leena Koulu | Hospital District of Southwest Finland, Turku, Finland | Clinical Groups | **Dermatology Group** |
| Nizar Smaoui | Abbvie, Chicago, IL, United States | Clinical Groups | **Dermatology Group** |
| Fedik Rahimov | Abbvie, Chicago, IL, United States | Clinical Groups | **Dermatology Group** |
| Anne Lehtonen | Abbvie, Chicago, IL, United States | Clinical Groups | **Dermatology Group** |
| David Choy | Genentech, San Francisco, CA, United States | Clinical Groups | **Dermatology Group** |
| Rion Pendergrass | Genentech, San Francisco, CA, United States | Clinical Groups | **Dermatology Group** |
| Dawn Waterworth | Janssen Research & Development, LLC, Spring House, PA, United States | Clinical Groups | **Dermatology Group** |
| Kirsi Kalpala | Pfizer, New York, NY, United States | Clinical Groups | **Dermatology Group** |
| Ying Wu | Pfizer, New York, NY, United States | Clinical Groups | **Dermatology Group** |
| Pirkko Pussinen | Hospital District of Helsinki and Uusimaa, Helsinki, Finland | Clinical Groups | **Odontology Group** |

| | | | |
|---|---|---|---|
| Aino Salminen | Hospital District of Helsinki and Uusimaa, Helsinki, Finland | Clinical Groups | **Odontology Group** |
| Tuula Salo | Hospital District of Helsinki and Uusimaa, Helsinki, Finland | Clinical Groups | **Odontology Group** |
| David Rice | Hospital District of Helsinki and Uusimaa, Helsinki, Finland | Clinical Groups | **Odontology Group** |
| Pekka Nieminen | Hospital District of Helsinki and Uusimaa, Helsinki, Finland | Clinical Groups | **Odontology Group** |
| Ulla Palotie | Hospital District of Helsinki and Uusimaa, Helsinki, Finland | Clinical Groups | **Odontology Group** |
| Maria Siponen | Northern Savo Hospital District, Kuopio, Finland | Clinical Groups | **Odontology Group** |
| Liisa Suominen | Northern Savo Hospital District, Kuopio, Finland | Clinical Groups | **Odontology Group** |
| Päivi Mäntylä | Northern Savo Hospital District, Kuopio, Finland | Clinical Groups | **Odontology Group** |
| Ulvi Gursoy | Hospital District of Southwest Finland, Turku, Finland | Clinical Groups | **Odontology Group** |
| Vuokko Anttonen | Northern Ostrobothnia Hospital District, Oulu, Finland | Clinical Groups | **Odontology Group** |
| Kirsi Sipilä | Research Unit of Oral Health Sciences Faculty of Medicine, University of Oulu, Oulu, Finland; Medical Research Center, Oulu, Oulu University Hospital and University of Oulu, Oulu, Finland | Clinical Groups | **Odontology Group** |
| Rion Pendergrass | Genentech, San Francisco, CA, United States | Clinical Groups | **Odontology Group** |
| Hannele Laivuori | Institute for Molecular Medicine Finland (FIMM), HiLIFE, University of Helsinki, Helsinki, Finland | Clinical Groups | **Women’s Health and Reproduction Group** |
| Venla Kurra | Pirkanmaa Hospital District, Tampere, Finland | Clinical Groups | **Women’s Health and Reproduction Group** |
| Laura Kotaniemi-Talonen | Pirkanmaa Hospital District, Tampere, Finland | Clinical Groups | **Women’s Health and Reproduction Group** |
| Oskari Heikinheimo | Hospital District of Helsinki and Uusimaa, Helsinki, Finland | Clinical Groups | **Women’s Health and Reproduction Group** |
| Ilkka Kalliala | Hospital District of Helsinki and Uusimaa, Helsinki, Finland | Clinical Groups | **Women’s Health and Reproduction Group** |
| Lauri Aaltonen | Hospital District of Helsinki and Uusimaa, Helsinki, Finland | Clinical Groups | **Women’s Health and Reproduction Group** |
| Varpu Jokimaa | Hospital District of Southwest Finland, Turku, Finland | Clinical Groups | **Women’s Health and Reproduction Group** |
| Johannes Kettunen | Northern Ostrobothnia Hospital District, Oulu, Finland | Clinical Groups | **Women’s Health and Reproduction Group** |

| | | | |
|---|---|---|---|
| Marja Vääräsmäki | Northern Ostrobothnia Hospital District, Oulu, Finland | Clinical Groups | **Women's Health and Reproduction Group** |
| Outi Uimari | Northern Ostrobothnia Hospital District, Oulu, Finland | Clinical Groups | **Women's Health and Reproduction Group** |
| Laure Morin-Papunen | Northern Ostrobothnia Hospital District, Oulu, Finland | Clinical Groups | **Women's Health and Reproduction Group** |
| Maarit Niinimäki | Northern Ostrobothnia Hospital District, Oulu, Finland | Clinical Groups | **Women's Health and Reproduction Group** |
| Terhi Piltonen | Northern Ostrobothnia Hospital District, Oulu, Finland | Clinical Groups | **Women's Health and Reproduction Group** |
| Katja Kivinen | Institute for Molecular Medicine Finland (FIMM), HiLIFE, University of Helsinki, Helsinki, Finland | Clinical Groups | **Women's Health and Reproduction Group** |
| Elisabeth Widen | Institute for Molecular Medicine Finland (FIMM), HiLIFE, University of Helsinki, Helsinki, Finland | Clinical Groups | **Women's Health and Reproduction Group** |
| Taru Tukiainen | Institute for Molecular Medicine Finland (FIMM), HiLIFE, University of Helsinki, Helsinki, Finland | Clinical Groups | **Women's Health and Reproduction Group** |
| Mary Pat Reeve | Institute for Molecular Medicine Finland (FIMM), HiLIFE, University of Helsinki, Helsinki, Finland | Clinical Groups | **Women's Health and Reproduction Group** |
| Mark Daly | Institute for Molecular Medicine Finland (FIMM), HiLIFE, University of Helsinki, Helsinki, Finland; Broad Institute of MIT and Harvard; Massachusetts General Hospital | Clinical Groups | **Women's Health and Reproduction Group** |
| Niko Välimäki | University of Helsinki, Helsinki, Finland | Clinical Groups | **Women's Health and Reproduction Group** |
| Eija Laakkonen | University of Jyväskylä, Jyväskylä, Finland | Clinical Groups | **Women's Health and Reproduction Group** |
| Jaakko Tyrmi | University of Oulu, Oulu, Finland / University of Tampere, Tampere, Finland | Clinical Groups | **Women's Health and Reproduction Group** |
| Heidi Silven | University of Oulu, Oulu, Finland | Clinical Groups | **Women's Health and Reproduction Group** |
| Eeva Sliz | University of Oulu, Oulu, Finland | Clinical Groups | **Women's Health and Reproduction Group** |
| Riikka Arffman | University of Oulu, Oulu, Finland | Clinical Groups | **Women's Health and Reproduction Group** |
| Susanna Savukoski | University of Oulu, Oulu, Finland | Clinical Groups | **Women's Health and Reproduction Group** |
| Triin Laisk | Estonian biobank, Tartu, Estonia | Clinical Groups | **Women's Health and Reproduction Group** |
| Natalia Pujol | Estonian biobank, Tartu, Estonia | Clinical Groups | **Women's Health and Reproduction Group** |
| Mengzhen Liu | Abbvie, Chicago, IL, United States | Clinical Groups | **Women's Health and Reproduction Group** |

| | | | |
|---|---|---|---|
| Bridget Riley-Gillis | Abbvie, Chicago, IL, United States | Clinical Groups | **Women's Health and Reproduction Group** |
| Rion Pendergrass | Genentech, San Francisco, CA, United States | Clinical Groups | **Women's Health and Reproduction Group** |
| Janet Kumar | GlaxoSmithKline, Collegeville, PA, United States | Clinical Groups | **Women's Health and Reproduction Group** |
| Kirsi Auro | GlaxoSmithKline, Espoo, Finland | Clinical Groups | **Women's Health and Reproduction Group** |
| Iiris Hovatta | University of Helsinki, Finland | Clinical Groups | **Depression group** |
| Chia-Yen Chen | Biogen, Cambridge, MA, United States | Clinical Groups | **Depression group** |
| Erkki Isometsä | Hospital District of Helsinki and Uusimaa, Helsinki, Finland | Clinical Groups | **Depression group** |
| Hanna Ollila | Institute for Molecular Medicine Finland (FIMM), HiLIFE, University of Helsinki, Helsinki, Finland | Clinical Groups | **Depression group** |
| Jaana Suvisaari | Finnish Institute for Health and Welfare (THL), Helsinki, Finland | Clinical Groups | **Depression group** |
| Thomas Damm Als | Aarhus University, Denmark | Clinical Groups | **Depression group** |
| Antti Mäkitie | Department of Otorhinolaryngology - Head and Neck Surgery, University of Helsinki and Helsinki University Hospital, Helsinki, Finland | Clinical Groups | **ENT (ear, nose and throath) Group** |
| Argyro Bizaki-Vallaskangas | Pirkanmaa Hospital District, Tampere, Finland | Clinical Groups | **ENT (ear, nose and throath) Group** |
| Sanna Toppila-Salmi | University of Helsinki, Finland | Clinical Groups | **ENT (ear, nose and throath) Group** |
| Tytti Willberg | Hospital District of Southwest Finland, Turku, Finland | Clinical Groups | **ENT (ear, nose and throath) Group** |
| Elmo Saarentaus | Institute for Molecular Medicine Finland (FIMM), HiLIFE, University of Helsinki, Helsinki, Finland | Clinical Groups | **ENT (ear, nose and throath) Group** |
| Antti Aarnisalo | Hospital District of Helsinki and Uusimaa, Helsinki, Finland | Clinical Groups | **ENT (ear, nose and throath) Group** |
| Eveliina Salminen | Hospital District of Helsinki and Uusimaa, Helsinki, Finland | Clinical Groups | **ENT (ear, nose and throath) Group** |
| Elisa Rahikkala | Northern Ostrobothnia Hospital District, Oulu, Finland | Clinical Groups | **ENT (ear, nose and throath) Group** |
| Johannes Kettunen | Northern Ostrobothnia Hospital District, Oulu, Finland | Clinical Groups | **ENT (ear, nose and throath) Group** |
| Kristiina Aittomäki | Department of Medical Genetics, Helsinki University Central Hospital, Helsinki, Finland | Clinical Groups | **POI (premature ovarian failure) Group** |

| | | | |
|---|---|---|---|
| Fredrik Åberg | Transplantation and Liver Surgery Clinic, Helsinki University Hospital, Helsinki University, Helsinki, Finland | Clinical Groups | **LiverScore Group** |
| Mitja Kurki | Institute for Molecular Medicine Finland (FIMM), HiLIFE, University of Helsinki, Helsinki, Finland; Broad Institute, Cambridge, MA, United States | **FinnGen Analysis working group** | **FinnGen Analysis working group** |
| Samuli Ripatti | Institute for Molecular Medicine Finland (FIMM), HiLIFE, University of Helsinki, Helsinki, Finland | **FinnGen Analysis working group** | **FinnGen Analysis working group** |
| Mark Daly | Institute for Molecular Medicine, Finland (FIMM), HiLIFE, University of Helsinki, Helsinki, Finland; Broad Institute of MIT and Harvard; Massachusetts General Hospital | **FinnGen Analysis working group** | **FinnGen Analysis working group** |
| Juha Karjalainen | Institute for Molecular Medicine Finland (FIMM), HiLIFE, University of Helsinki, Helsinki, Finland | **FinnGen Analysis working group** | **FinnGen Analysis working group** |
| Aki Havulinna | Institute for Molecular Medicine Finland (FIMM), HiLIFE, University of Helsinki, Helsinki, Finland; Finnish Institute for Health and Welfare (THL), Helsinki, Finland | **FinnGen Analysis working group** | **FinnGen Analysis working group** |
| Juha Mehtonen | Institute for Molecular Medicine Finland (FIMM), HiLIFE, University of Helsinki, Helsinki, Finland | **FinnGen Analysis working group** | **FinnGen Analysis working group** |
| Priit Palta | Institute for Molecular Medicine Finland (FIMM), HiLIFE, University of Helsinki, Helsinki, Finland | **FinnGen Analysis working group** | **FinnGen Analysis working group** |
| Shabbeer Hassan | Institute for Molecular Medicine Finland (FIMM), HiLIFE, University of Helsinki, Helsinki, Finland | **FinnGen Analysis working group** | **FinnGen Analysis working group** |
| Pietro Della Briotta Parolo | Institute for Molecular Medicine Finland (FIMM), HiLIFE, University of Helsinki, Helsinki, Finland | **FinnGen Analysis working group** | **FinnGen Analysis working group** |
| Wei Zhou | Broad Institute, Cambridge, MA, United States | **FinnGen Analysis working group** | **FinnGen Analysis working group** |
| Mutaamba Maasha | Broad Institute, Cambridge, MA, United States | **FinnGen Analysis working group** | **FinnGen Analysis working group** |
| Shabbeer Hassan | Institute for Molecular Medicine Finland (FIMM), HiLIFE, University of Helsinki, Helsinki, Finland | **FinnGen Analysis working group** | **FinnGen Analysis working group** |
| Susanna Lemmelä | Institute for Molecular Medicine Finland (FIMM), HiLIFE, University of Helsinki, Helsinki, Finland | **FinnGen Analysis working group** | **FinnGen Analysis working group** |

| | | | |
|---|---|---|---|
| Manuel Rivas | University of Stanford, Stanford, CA, United States | FinnGen Analysis working group | FinnGen Analysis working group |
| Aarno Palotie | Institute for Molecular Medicine Finland (FIMM), HiLIFE, University of Helsinki, Helsinki, Finland | FinnGen Analysis working group | FinnGen Analysis working group |
| Aoxing Liu | Institute for Molecular Medicine Finland (FIMM), HiLIFE, University of Helsinki, Helsinki, Finland | FinnGen Analysis working group | FinnGen Analysis working group |
| Arto Lehisto | Institute for Molecular Medicine Finland (FIMM), HiLIFE, University of Helsinki, Helsinki, Finland | FinnGen Analysis working group | FinnGen Analysis working group |
| Andrea Ganna | Institute for Molecular Medicine Finland (FIMM), HiLIFE, University of Helsinki, Helsinki, Finland | FinnGen Analysis working group | FinnGen Analysis working group |
| Vincent Llorens | Institute for Molecular Medicine Finland (FIMM), HiLIFE, University of Helsinki, Helsinki, Finland | FinnGen Analysis working group | FinnGen Analysis working group |
| Hannele Laivuori | Institute for Molecular Medicine Finland (FIMM), HiLIFE, University of Helsinki, Helsinki, Finland | FinnGen Analysis working group | FinnGen Analysis working group |
| Taru Tukiainen | Institute for Molecular Medicine Finland (FIMM), HiLIFE, University of Helsinki, Helsinki, Finland | FinnGen Analysis working group | FinnGen Analysis working group |
| Mary Pat Reeve | Institute for Molecular Medicine Finland (FIMM), HiLIFE, University of Helsinki, Helsinki, Finland | FinnGen Analysis working group | FinnGen Analysis working group |
| Henrike Heyne | Institute for Molecular Medicine Finland (FIMM), HiLIFE, University of Helsinki, Helsinki, Finland | FinnGen Analysis working group | FinnGen Analysis working group |
| Nina Mars | Institute for Molecular Medicine Finland (FIMM), HiLIFE, University of Helsinki, Helsinki, Finland | FinnGen Analysis working group | FinnGen Analysis working group |
| Joel Rämö | Institute for Molecular Medicine Finland (FIMM), HiLIFE, University of Helsinki, Helsinki, Finland | FinnGen Analysis working group | FinnGen Analysis working group |
| Elmo Saarentaus | Institute for Molecular Medicine Finland (FIMM), HiLIFE, University of Helsinki, Helsinki, Finland | FinnGen Analysis working group | FinnGen Analysis working group |

| | | | |
|---|---|---|---|
| Hanna Ollila | Institute for Molecular Medicine Finland (FIMM), HiLIFE, University of Helsinki, Helsinki, Finland | FinnGen Analysis working group | FinnGen Analysis working group |
| Rodos Rodosthenous | Institute for Molecular Medicine Finland (FIMM), HiLIFE, University of Helsinki, Helsinki, Finland | FinnGen Analysis working group | FinnGen Analysis working group |
| Satu Strausz | Institute for Molecular Medicine Finland (FIMM), HiLIFE, University of Helsinki, Helsinki, Finland | FinnGen Analysis working group | FinnGen Analysis working group |
| Tuula Palotie | University of Helsinki and Hospital District of Helsinki and Uusimaa, Helsinki, Finland | FinnGen Analysis working group | FinnGen Analysis working group |
| Kimmo Palin | University of Helsinki, Helsinki, Finland | FinnGen Analysis working group | FinnGen Analysis working group |
| Javier Garcia-Tabuenca | University of Tampere, Tampere, Finland | FinnGen Analysis working group | FinnGen Analysis working group |
| Harri Siirtola | University of Tampere, Tampere, Finland | FinnGen Analysis working group | FinnGen Analysis working group |
| Tuomo Kiiskinen | Institute for Molecular Medicine Finland (FIMM), HiLIFE, University of Helsinki, Helsinki, Finland | FinnGen Analysis working group | FinnGen Analysis working group |
| Jiwoo Lee | Institute for Molecular Medicine Finland (FIMM), HiLIFE, University of Helsinki, Helsinki, Finland; Broad Institute, Cambridge, MA, United States | FinnGen Analysis working group | FinnGen Analysis working group |
| Kristin Tsuo | Institute for Molecular Medicine Finland (FIMM), HiLIFE, University of Helsinki, Helsinki, Finland; Broad Institute, Cambridge, MA, United States | FinnGen Analysis working group | FinnGen Analysis working group |
| Amanda Elliott | Institute for Molecular Medicine Finland (FIMM), HiLIFE, University of Helsinki, Helsinki, Finland; Broad Institute, Cambridge, MA, USA and Massachusetts General Hospital, Boston, MA, USA | FinnGen Analysis working group | FinnGen Analysis working group |
| Kati Kristiansson | THL Biobank / Finnish Institute for Health and Welfare (THL), Helsinki, Finland | FinnGen Analysis working group | FinnGen Analysis working group |
| Mikko Arvas | Finnish Red Cross Blood Service / Finnish Hematology Registry and Clinical Biobank, Helsinki, Finland | FinnGen Analysis working group | FinnGen Analysis working group |

| | | | |
|---|---|---|---|
| Kati Hyvärinen | Finnish Red Cross Blood Service, Helsinki, Finland | FinnGen Analysis working group | FinnGen Analysis working group |
| Jarmo Ritari | Finnish Red Cross Blood Service, Helsinki, Finland | FinnGen Analysis working group | FinnGen Analysis working group |
| Olli Carpén | Helsinki Biobank / Helsinki University and Hospital District of Helsinki and Uusimaa, Helsinki | FinnGen Analysis working group | FinnGen Analysis working group |
| Johannes Kettunen | Northern Finland Biobank Borealis / University of Oulu / Northern Ostrobothnia Hospital District, Oulu, Finland | FinnGen Analysis working group | FinnGen Analysis working group |
| Katri Pylkäs | University of Oulu, Oulu, Finland | FinnGen Analysis working group | FinnGen Analysis working group |
| Eeva Sliz | University of Oulu, Oulu, Finland | FinnGen Analysis working group | FinnGen Analysis working group |
| Minna Karjalainen | University of Oulu, Oulu, Finland | FinnGen Analysis working group | FinnGen Analysis working group |
| Tuomo Mantere | Northern Finland Biobank Borealis / University of Oulu / Northern Ostrobothnia Hospital District, Oulu, Finland | FinnGen Analysis working group | FinnGen Analysis working group |
| Eeva Kangasniemi | Finnish Clinical Biobank Tampere / University of Tampere / Pirkanmaa Hospital District, Tampere, Finland | FinnGen Analysis working group | FinnGen Analysis working group |
| Sami Heikkinen | University of Eastern Finland, Kuopio, Finland | FinnGen Analysis working group | FinnGen Analysis working group |
| Arto Mannermaa | Biobank of Eastern Finland / University of Eastern Finland / Northern Savo Hospital District, Kuopio, Finland | FinnGen Analysis working group | FinnGen Analysis working group |
| Eija Laakkonen | University of Jyväskylä, Jyväskylä, Finland | FinnGen Analysis working group | FinnGen Analysis working group |
| Nina Pitkänen | Auria Biobank / University of Turku / Hospital District of Southwest Finland, Turku, Finland | FinnGen Analysis working group | FinnGen Analysis working group |

| | | | |
|---|---|---|---|
| Samuel Lessard | Translational Sciences, Sanofi R&D, Framingham, MA, USA | FinnGen Analysis working group | FinnGen Analysis working group |
| Clément Chatelain | Translational Sciences, Sanofi R&D, Framingham, MA, USA | FinnGen Analysis working group | FinnGen Analysis working group |
| Perttu Terho | Auria Biobank / University of Turku / Hospital District of Southwest Finland, Turku, Finland | Biobank directors | Biobank directors |
| Tiina Wahlfors | THL Biobank / Finnish Institute for Health and Welfare (THL), Helsinki, Finland | Biobank directors | Biobank directors |
| Jukka Partanen | Finnish Red Cross Blood Service / Finnish Hematology Registry and Clinical Biobank, Helsinki, Finland | Biobank directors | Biobank directors |
| Eero Punkka | Helsinki Biobank / Helsinki University and Hospital District of Helsinki and Uusimaa, Helsinki | Biobank directors | Biobank directors |
| Raisa Serpi | Northern Finland Biobank Borealis / University of Oulu / Northern Ostrobothnia Hospital District, Oulu, Finland | Biobank directors | Biobank directors |
| Sanna Siltanen | Finnish Clinical Biobank Tampere / University of Tampere / Pirkanmaa Hospital District, Tampere, Finland | Biobank directors | Biobank directors |
| Veli-Matti Kosma | Biobank of Eastern Finland / University of Eastern Finland / Northern Savo Hospital District, Kuopio, Finland | Biobank directors | Biobank directors |
| Teijo Kuopio | Central Finland Biobank / University of Jyväskylä / Central Finland Health Care District, Jyväskylä, Finland | Biobank directors | Biobank directors |
| Anu Jalanko | Institute for Molecular Medicine Finland (FIMM), HiLIFE, University of Helsinki, Helsinki, Finland | FinnGen Teams | **Administration** |
| Huei-Yi Shen | Institute for Molecular Medicine Finland (FIMM), HiLIFE, University of Helsinki, Helsinki, Finland | FinnGen Teams | **Administration** |
| Risto Kajanne | Institute for Molecular Medicine Finland (FIMM), HiLIFE, University of Helsinki, Helsinki, Finland | FinnGen Teams | **Administration** |
| Mervi Aavikko | Institute for Molecular Medicine Finland (FIMM), HiLIFE, University of Helsinki, Helsinki, Finland | FinnGen Teams | **Administration** |
| Rasko Leinonen | Institute for Molecular Medicine Finland (FIMM), HiLIFE, University of Helsinki, Helsinki, Finland; European Molecular Biology Laboratory, European Bioinformatics Institute, Cambridge, UK | FinnGen Teams | **Administration** |

| | | | |
|---|---|---|---|
| Henna Palin | Finnish Clinical Biobank Tampere / University of Tampere / Pirkanmaa Hospital District, Tampere, Finland | FinnGen Teams | **Administration** |
| Malla-Maria Linna | Helsinki Biobank / Helsinki University and Hospital District of Helsinki and Uusimaa, Helsinki | FinnGen Teams | **Administration** |
| Mitja Kurki | Institute for Molecular Medicine Finland (FIMM), HiLIFE, University of Helsinki, Helsinki, Finland; Broad Institute, Cambridge, MA, United States | FinnGen Teams | **Analysis** |
| Juha Karjalainen | Institute for Molecular Medicine Finland (FIMM), HiLIFE, University of Helsinki, Helsinki, Finland | FinnGen Teams | **Analysis** |
| Pietro Della Briotta Parolo | Institute for Molecular Medicine Finland (FIMM), HiLIFE, University of Helsinki, Helsinki, Finland | FinnGen Teams | **Analysis** |
| Arto Lehisto | Institute for Molecular Medicine Finland (FIMM), HiLIFE, University of Helsinki, Helsinki, Finland | FinnGen Teams | **Analysis** |
| Juha Mehtonen | Institute for Molecular Medicine Finland (FIMM), HiLIFE, University of Helsinki, Helsinki, Finland | FinnGen Teams | **Analysis** |
| Wei Zhou | Broad Institute, Cambridge, MA, United States | FinnGen Teams | **Analysis** |
| Masahiro Kanai | Broad Institute, Cambridge, MA, United States | FinnGen Teams | **Analysis** |
| Mutaamba Maasha | Broad Institute, Cambridge, MA, United States | FinnGen Teams | **Analysis** |
| Zhili Zheng | Broad Institute, Cambridge, MA, United States | FinnGen Teams | **Analysis** |
| Hannele Laivuori | Institute for Molecular Medicine Finland (FIMM), HiLIFE, University of Helsinki, Helsinki, Finland | FinnGen Teams | **Clinical Endpoint Development** |
| Aki Havulinna | Institute for Molecular Medicine Finland (FIMM), HiLIFE, University of Helsinki, Helsinki, Finland; Finnish Institute for Health and Welfare (THL), Helsinki, Finland | FinnGen Teams | **Clinical Endpoint Development** |
| Susanna Lemmelä | Institute for Molecular Medicine Finland (FIMM), HiLIFE, University of Helsinki, Helsinki, Finland | FinnGen Teams | **Clinical Endpoint Development** |
| Tuomo Kiiskinen | Institute for Molecular Medicine Finland (FIMM), HiLIFE, University of Helsinki, Helsinki, Finland | FinnGen Teams | **Clinical Endpoint Development** |
| L. Elisa Lahtela | Institute for Molecular Medicine Finland (FIMM), HiLIFE, University of Helsinki, Helsinki, Finland | FinnGen Teams | **Clinical Endpoint Development** |
| Mari Kaunisto | Institute for Molecular Medicine Finland (FIMM), HiLIFE, University of Helsinki, Helsinki, Finland | FinnGen Teams | **Communication** |
| Elina Kilpeläinen | Institute for Molecular Medicine Finland (FIMM), HiLIFE, University of Helsinki, Helsinki, Finland | FinnGen Teams | **E-Science** |

| | | | |
|---|---|---|---|
| Timo P. Sipilä | Institute for Molecular Medicine Finland (FIMM), HiLIFE, University of Helsinki, Helsinki, Finland | FinnGen Teams | **E-Science** |
| Oluwaseun Alexander Dada | Institute for Molecular Medicine Finland (FIMM), HiLIFE, University of Helsinki, Helsinki, Finland | FinnGen Teams | **E-Science** |
| Awaisa Ghazal | Institute for Molecular Medicine Finland (FIMM), HiLIFE, University of Helsinki, Helsinki, Finland | FinnGen Teams | **E-Science** |
| Anastasia Kytölä | Institute for Molecular Medicine Finland (FIMM), HiLIFE, University of Helsinki, Helsinki, Finland | FinnGen Teams | **E-Science** |
| Rigbe Weldatsadik | Institute for Molecular Medicine Finland (FIMM), HiLIFE, University of Helsinki, Helsinki, Finland | FinnGen Teams | **E-Science** |
| Sanni Ruotsalainen | Institute for Molecular Medicine Finland (FIMM), HiLIFE, University of Helsinki, Helsinki, Finland | FinnGen Teams | **E-Science** |
| Kati Donner | Institute for Molecular Medicine Finland (FIMM), HiLIFE, University of Helsinki, Helsinki, Finland | FinnGen Teams | **Genotyping** |
| Timo P. Sipilä | Institute for Molecular Medicine Finland (FIMM), HiLIFE, University of Helsinki, Helsinki, Finland | FinnGen Teams | **Genotyping** |
| Anu Loukola | Helsinki Biobank / Helsinki University and Hospital District of Helsinki and Uusimaa, Helsinki | FinnGen Teams | **Sample Collection Coordination** |
| Päivi Laiho | THL Biobank / Finnish Institute for Health and Welfare (THL), Helsinki, Finland | FinnGen Teams | **Sample Logistics** |
| Tuuli Sistonen | THL Biobank / Finnish Institute for Health and Welfare (THL), Helsinki, Finland | FinnGen Teams | **Sample Logistics** |
| Essi Kaiharju | THL Biobank / Finnish Institute for Health and Welfare (THL), Helsinki, Finland | FinnGen Teams | **Sample Logistics** |
| Markku Laukkanen | THL Biobank / Finnish Institute for Health and Welfare (THL), Helsinki, Finland | FinnGen Teams | **Sample Logistics** |
| Elina Järvensivu | THL Biobank / Finnish Institute for Health and Welfare (THL), Helsinki, Finland | FinnGen Teams | **Sample Logistics** |
| Sini Lähteenmäki | THL Biobank / Finnish Institute for Health and Welfare (THL), Helsinki, Finland | FinnGen Teams | **Sample Logistics** |
| Lotta Männikkö | THL Biobank / Finnish Institute for Health and Welfare (THL), Helsinki, Finland | FinnGen Teams | **Sample Logistics** |
| Regis Wong | THL Biobank / Finnish Institute for Health and Welfare (THL), Helsinki, Finland | FinnGen Teams | **Sample Logistics** |

| | | | |
|---|---|---|---|
| Auli Toivola | THL Biobank / Finnish Institute for Health and Welfare (THL), Helsinki, Finland | FinnGen Teams | **Sample Logistics** |
| Minna Brunfeldt | THL Biobank / Finnish Institute for Health and Welfare (THL), Helsinki, Finland | FinnGen Teams | **Registry Data Operations** |
| Hannele Mattsson | THL Biobank / Finnish Institute for Health and Welfare (THL), Helsinki, Finland | FinnGen Teams | **Registry Data Operations** |
| Kati Kristiansson | THL Biobank / Finnish Institute for Health and Welfare (THL), Helsinki, Finland | FinnGen Teams | **Registry Data Operations** |
| Susanna Lemmelä | Institute for Molecular Medicine Finland (FIMM), HiLIFE, University of Helsinki, Helsinki, Finland | FinnGen Teams | **Registry Data Operations** |
| Sami Koskelainen | THL Biobank / Finnish Institute for Health and Welfare (THL), Helsinki, Finland | FinnGen Teams | **Registry Data Operations** |
| Tero Hiekkalinna | THL Biobank / Finnish Institute for Health and Welfare (THL), Helsinki, Finland | FinnGen Teams | **Registry Data Operations** |
| Teemu Paajanen | THL Biobank / Finnish Institute for Health and Welfare (THL), Helsinki, Finland | FinnGen Teams | **Registry Data Operations** |
| Priit Palta | Institute for Molecular Medicine Finland (FIMM), HiLIFE, University of Helsinki, Helsinki, Finland | FinnGen Teams | **Sequencing Informatics** |
| Kalle Pärn | Institute for Molecular Medicine Finland (FIMM), HiLIFE, University of Helsinki, Helsinki, Finland | FinnGen Teams | **Sequencing Informatics** |
| Mart Kals | Institute for Molecular Medicine Finland (FIMM), HiLIFE, University of Helsinki, Helsinki, Finland | FinnGen Teams | **Sequencing Informatics** |
| Shuang Luo | Institute for Molecular Medicine Finland (FIMM), HiLIFE, University of Helsinki, Helsinki, Finland | FinnGen Teams | **Sequencing Informatics** |
| Tarja Laitinen | Pirkanmaa Hospital District, Tampere, Finland | FinnGen Teams | **Traject ory** |
| Mary Pat Reeve | Institute for Molecular Medicine Finland (FIMM), HiLIFE, University of Helsinki, Helsinki, Finland | FinnGen Teams | **Traject ory** |
| Shanmukha Sampath Padmanabhuni | Institute for Molecular Medicine Finland (FIMM), HiLIFE, University of Helsinki, Helsinki, Finland | FinnGen Teams | **Traject ory** |
| Marianna Niemi | University of Tampere, Tampere, Finland | FinnGen Teams | **Traject ory** |
| Harri Siirtola | University of Tampere, Tampere, Finland | FinnGen Teams | **Traject ory** |
| Javier Gracia-Tabuenca | University of Tampere, Tampere, Finland | FinnGen Teams | **Traject ory** |
| Mika Helminen | University of Tampere, Tampere, Finland | FinnGen Teams | **Traject ory** |
| Tiina Luukkaala | University of Tampere, Tampere, Finland | FinnGen Teams | **Traject ory** |

| | | | |
|---|---|---|---|
| Iida Vähätalo | University of Tampere, Tampere, Finland | FinnGen Teams | **Trajectory** |
| Jyrki Tammerluoto | Institute for Molecular Medicine Finland (FIMM), HiLIFE, University of Helsinki, Helsinki, Finland | FinnGen Teams | **Data protection officer** |
| Marco Hautalahti | Finnish Biobank Cooperative - FINBB | FinnGen Teams | **FINBB - Finnish biobank cooperative** |
| Johanna Mäkelä | Finnish Biobank Cooperative - FINBB | FinnGen Teams | **FINBB - Finnish biobank cooperative** |
| Sarah Smith | Finnish Biobank Cooperative - FINBB | FinnGen Teams | **FINBB - Finnish biobank cooperative** |
| Tom Southerington | Finnish Biobank Cooperative - FINBB | FinnGen Teams | **FINBB - Finnish biobank cooperative** |
| Petri Lehto | Finnish Biobank Cooperative - FINBB | FinnGen Teams | **FINBB - Finnish biobank cooperative** |
